# TrialCode Agent: LLM-Assisted Clinical Code-Set Construction for Trial Emulation

**DOI:** 10.64898/2026.08.20.26360962

**Authors:** Amir Habibdoust, Aria Sajjad, Diana Hernandez, Kashyap Patel, Xing Song

## Abstract

**Objective:** Translating free-text clinical trial criteria into computable code sets is a valuable standardization practice that is necessary for producing reproducible real-world evidence studies but requires standardized interpretation across multiple clinical vocabularies.

**Methods:** We developed TrialCode Agent, a hybrid-large language model (LLM)-terminology verification agent that generates, formats, verifies, and expands candidate codes from free-text clinical criteria. The system supports ICD-9-CM diagnoses and procedures, ICD-10-CM, ICD-10-PCS, LOINC, and RxNorm medication concepts. We compared Baseline, Hybrid biomedical retrieval-augmented generation (RAG), and terminology-guided Family expansion pipelines using Claude, GPT Qwen, and MedGemma on 40 criteria from 11 trial groups. Performance was evaluated against expert-built reference code sets using exact-code precision, recall, and F1.

**Results:** The optimal pipeline varied by model. Claude with Baseline achieved the highest performance (precision 0.755, recall 0.619, F1 0.680), followed by GPT-5.5 with Baseline (precision 0.569, recall 0.658, F1 0.610), Qwen with Hybrid biomedical RAG (precision 0.656, recall 0.470, F1 0.548), and MedGemma with Family expansion (precision 0.487, recall 0.316, F1 0.383). Hybrid biomedical RAG improved aggregate F1 only for Qwen but increased GPT-5.5 RxNorm F1 from 0.320 to 0.909. Macro-averaged results showed criterion-level gains despite lower micro-averaged aggregate performance. Family expansion increased recall across models but generally reduced precision. In staged verifier ablation, micro-F1 increased from 0.254 before verification to 0.505 after final verification and expansion. Existence/vocabulary checking removed 2,594 false-positive codes, and acceptance filtering removed 952 additional false-positive codes before controlled expansion.

**Conclusions:** Combining LLM-based clinical interpretation with deterministic terminology verification produces auditable, database-ready code sets, but retrieval and broad family expansion do not consistently improve exact-code performance. Retrieval was particularly useful for RxNorm mapping, whereas overly broad or incomplete candidate generation remained the main source of error. Deterministic verification improves code validity and query readiness but cannot replace accurate clinical interpretation.

## 1 Introduction

Real-world evidence studies increasingly rely on electronic health records (EHRs), claims, registries, and laboratory databases to define cohorts, exposures, outcomes, eligibility criteria, and baseline covariates. In target trial emulation (TTE)^1–3^, this translation step is especially important because trial protocol language must be converted into computable definitions that can be applied consistently to real-world data. Recent work on automating causal inference, especially TTE, has highlighted the need for scalable systems that can translate trial protocols into reproducible, real-world data representations while preserving clinical and causal validity^4–6^. A major bottleneck in this process is clinical code mapping: the identification and curation of diagnosis, procedure, laboratory, and medication codes that accurately operationalize each protocol-defined clinical concept.

This mapping process is difficult because clinical vocabularies are hierarchical, versioned, and vocabulary-specific ^7,8^. A clinically correct concept can still produce an unusable database definition if the mapping returns a parent category instead of final child codes, confuses diagnosis and procedure vocabularies, or treats laboratory thresholds as if they were part of the laboratory identifier. Other issues include confusing medication ingredients, strengths, branded products, and fixed-dose combination products in RxNorm^9,10^. Research code-set construction requires more than identifying a single representative code; it requires complete, vocabulary-aware code sets that are directly executable against clinical databases. For example, an ICD-10-CM parent category may represent a clinically meaningful diabetes complication, whereas database queries often require the full set of valid billable child codes to accurately identify eligible patients. Likewise, LOINC specifies the laboratory test itself, but eligibility criteria such as an eGFR between 25 and 75 mL/min/1.73 m² must be applied separately to the corresponding laboratory result values. Together, these examples illustrate that translating clinical concepts into computable phenotypes requires both comprehensive terminology coverage and appropriate application of clinical logic.

LLMs offer a promising way to assist with clinical code mapping because they can interpret clinical language, recognize synonyms, and reason across incomplete or informal descriptions. However, LLM-only code generation remains risky. Prior studies have shown poor performance when general-purpose LLMs are used as standalone medical code query systems, although domain-specific fine-tuning and agentic coding workflows can improve ICD coding from clinical documentation^8,11,12^. Separately, systems for eligibility criteria processing and patient-trial matching have focused on normalizing trial criteria, matching patients to trials, or generating executable database queries rather than producing reusable research code sets^10,13,14^. These studies address important adjacent tasks, but they do not directly evaluate the intermediate task of research code-set construction which is translating a free-text clinical criterion into a complete, terminology-verified, multi-vocabulary code set artifacts.

In this work, we present a hybrid LLM–terminology verification agent for clinical code mapping. The system uses a generate-then-verify design. A first LLM call interprets the free-text clinical concept and proposes candidate mappings, a second LLM call formats the response into a structured code list, and a deterministic terminology-verification layer checks code existence, expands supported parent or prefix codes into valid child codes, and separates coded concepts from non-code logic such as laboratory thresholds. The objective of this study is to describe and evaluate this agent as infrastructure for reproducible real-world data research and TTE. We benchmarked four contemporary LLM configurations, including two proprietary models (Claude and GPT-5.5) and two open-weight models (Qwen and MedGemma), together with two enhancement strategies: hybrid biomedical terminology RAG and terminology-guided family expansion. Performance was evaluated against manually curated reference code sets reviewed by subject-matter experts.

## 2 Related Work

Prior work on LLMs for medical coding has mainly focused on direct code retrieval or encounter-level coding. Soroush et al. (2024)^11^ evaluated LLMs as direct medical code query systems, where the model was given an official code description and asked to return a matching ICD-9-CM, ICD-10-CM, or CPT code. Their findings showed that general-purpose LLMs remain unreliable as standalone medical coders. Similarly, de Almeida et al. (2026)^15^ evaluated LLMs as code selectors using ICPC-2 and showed the value of constraining LLM outputs with retrieved candidate concepts. Hou et al. (2025)^12^ addressed the problem through ICD-10-specific fine-tuning, showing improved robustness to lexical variation, abbreviations, and clinical-note inputs. These studies are important, but they primarily address single-code querying, code selection, or ICD-focused prediction rather than research code-set construction across multiple vocabularies.

A second related literature focuses on automated ICD coding from clinical notes. Transformer-based systems such as PLM-ICD frame coding as a multi-label classification task, where the input is a discharge summary or clinical note and the output is an encounter-level set of diagnosis codes ^16^. More recently, Motzfeldt et al. (2025)^8^ proposed Code Like Humans, a multi-agent ICD coding framework that follows aspects of the human coding workflow, including evidence extraction, index navigation, tabular validation, and code reconciliation. This work is conceptually related to our use of terminology resources and verification, but its goal is clinical-note coding. In contrast, our system starts from free-text research criteria and clinical concepts and produces reusable, terminology-verified code sets for real-world data studies.

Electronic phenotyping and cohort discovery provide a closer conceptual foundation for our work. Phenotype repositories such as PheKB emphasize the creation, validation, sharing, and reuse of computable phenotype algorithms ^17^. In real-world data research, code sets are not only billing outputs. They define cohorts, exposures, outcomes, eligibility criteria, and covariates. However, phenotype repositories and mapping systems often begin from existing curated definitions, whereas our system begins from free-text criteria and uses LLM interpretation followed by deterministic terminology verification.

Several systems have also translated trial eligibility criteria into computable representations. Lee et al. (2025)^9^ developed an LLM-based pipeline for converting ClinicalTrials.gov eligibility criteria into OMOP CDM-compatible SQL queries, including preprocessing, concept mapping, SQL generation, and hallucination analysis. Lee et al. (2024)^10^ developed CriteriaMapper to normalize trial eligibility criteria and EHR patient characteristics using standard terminologies and in-house reference tables. Dobbins et al. (2023)^13^ introduced LeafAI for generating cohort discovery queries from free-text eligibility criteria using named entity recognition, relation extraction, logical forms, UMLS normalization, and knowledge-base reasoning. Yang et al. (2026)^13^ proposed EC2Seq2Sql, which combines semantic parsing and an LLM SQL agent to convert eligibility criteria into executable SQL. These systems are closely related because they transform eligibility criteria into computable forms, but their final targets are cohort queries, OMOP SQL, or patient matching rather than reusable ICD-9-CM, ICD-9 procedure, ICD-10-CM, ICD-10-PCS, and LOINC code artifacts sets.

LLM-based patient–trial matching represents another adjacent direction. Callies et al. (2025)^18^ developed a multimodal LLM pipeline that uses unprocessed EHR documents, retrieval, and reasoning models to assess trial eligibility. Chen et al. (2025)^19^ found in a scoping review that LLM-based patient–trial matching systems commonly include eligibility-criteria processing, patient-data processing, and matching modules. These systems evaluate whether patients match trials, whereas our work targets an earlier infrastructure step of translating trial-style criteria into terminology-verified code sets that can be used in broader context.

Across these literatures, prior systems generally address direct code querying, ICD coding from notes, electronic phenotyping, cohort query generation, or patient–trial matching. Our contribution is a generate-then-verify LLM architecture for research code-set construction. The LLM is used for clinical interpretation and structured formatting, while deterministic terminology verification checks code existence, preserves vocabulary boundaries, expands supported parent categories, prefixes, wildcard patterns, and ranges under safety limits, and separates coded concepts from non-code logic such as laboratory thresholds. Thus, the novelty is not simply using multiple vocabularies, but combining LLM-based interpretation with database-grounded verification to produce code sets suitable for real-world data research and target trial emulation.

## 3 Methods

### 3.1 Study design

We developed a hybrid LLM–terminology verification agent for clinical code-set construction from free-text clinical criteria. The task was defined as translating a natural-language criterion into a database-ready code set across multiple vocabularies used in real-world data research. The supported vocabularies included ICD-9-CM diagnosis codes^20^, ICD-9 procedure codes^21^ (labeled ICD-9-PCS in the system), ICD-10-CM diagnosis codes17, ICD-10-PCS procedure codes^22^, Logical Observation Identifiers Names and Codes (LOINC)^23^, and RxNorm medication concepts^24^. The system was evaluated under three conditions: a baseline non-retrieval pipeline (Baseline), a hybrid biomedical terminology retrieval-augmented pipeline (Hybrid biomedical RAG), and a terminology-guided family-expansion pipeline (Family expansion). In all conditions, the formatter and deterministic terminology-verification layer were held constant.

### 3.2 System architecture

The agent used a generate–format–verify architecture. First, an LLM-based generation step interpreted the input clinical criterion and produced a free-text expert coding answer. This answer could include single codes, parent categories, prefixes, ranges, wildcard expressions, or explanatory grouping by clinical sub-concept. Second, a separate LLM-based formatting step converted the free-text answer into structured code entries grouped by vocabulary. The formatter was instructed not to add, remove, correct, or clinically reinterpret codes, but only to restructure the first answer into machine-readable format. Third, a deterministic terminology-verification layer checked the formatted code entries against local terminology tables and produced the final accepted code set. In the Hybrid biomedical RAG condition, retrieval and refinement were inserted between generation and formatting, yielding a generate-retrieve-refine-format-verify sequence. The Baseline condition used generate-format-verify. The Family expansion condition also used generate-format-verify but replaced the first-call generation prompt with a family-selection prompt. The overall system workflow is summarized in Figure 1 and entailed in the following sections.

**Figure 1.**
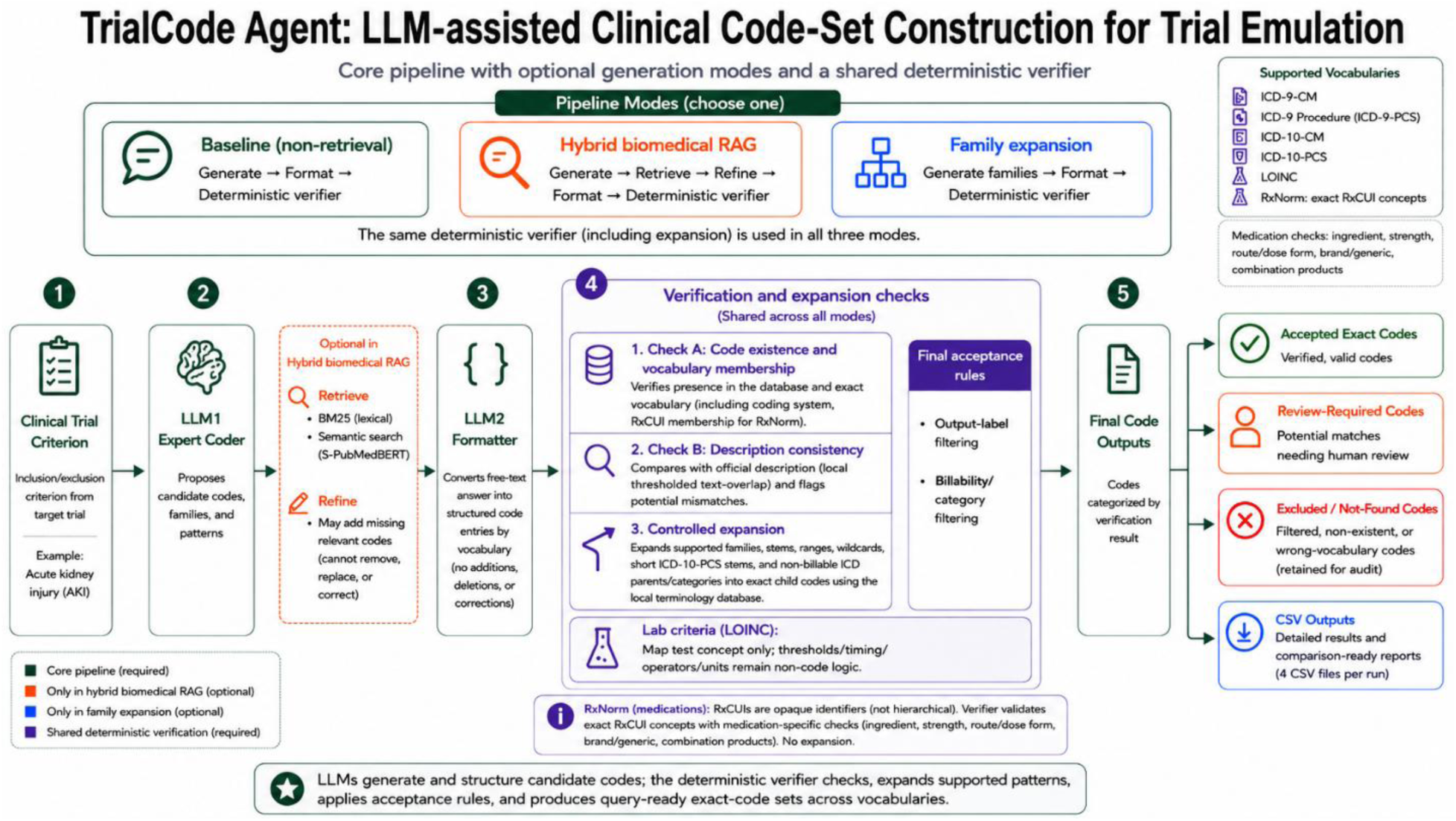
TrialCode Agent workflow and terminology-verification process. Draft created with OpenAI ChatGPT and subsequently edited and verified by the authors.

### 3.3 Baseline non-retrieval pipeline

In the Baseline condition, the input criterion was passed directly through the generate–format– verify sequence without retrieval augmentation.

### 3.4 Hybrid biomedical terminology retrieval-augmented pipeline

In the Hybrid biomedical RAG condition, the initial criterion and LLM1 answer were used as the query for hybrid retrieval from the local ICD/LOINC/RxNorm terminology database. Retrieval combined BM25 lexical retrieval, augmented by a manually defined clinical synonym dictionary, with semantic retrieval using the S-PubMedBert-MS-MARCO sentence-embedding model^25–28^. The lexical and semantic rankings were combined using reciprocal-rank fusion^29^. Hybrid-retrieval returned up to 80 candidates after reciprocal-rank fusion, and the top 60 candidates were supplied to the refinement LLM. Retrieved candidates were supplied to the LLM in a refinement step that was instructed to add only missing, directly relevant codes and not to remove, replace, or correct codes from the initial answer. The refined answer then passed through the same formatter and deterministic verifier used in the Baseline condition.

### 3.5 Terminology-guided family-expansion pipeline

The Family expansion condition used the same generate–format–verify sequence as the Baseline pipeline and did not use retrieval. The only change was to the first LLM call. Instead of a prompt that produced an expert coding answer in any form, the model was given a family-selection prompt that asked it to identify the clinical concept and select database-expandable code families, stems, ranges, or wildcard patterns, using a broad pattern only when every valid child code under it would satisfy the criterion. The model was instructed to prefer narrower stems, prefixes, or specific codes, and, for ICD-10-PCS, body-part-specific prefixes, when a broad family would also include non-qualifying children. The intent was to shift completeness from LLM enumeration to deterministic database expansion. The resulting answer was passed to the same formatter and verifier used in the other two conditions.

### 3.6 Terminology verification and expansion

The deterministic terminology-grounded verification consisted of three rule-based checks:

Check A - *Code existence and vocabulary membership check:* It confirmed whether each proposed code was present in the local terminology database and whether it belonged to the specified coding system, including RxCUI membership for RxNorm medication concepts. For RxNorm entries, verification additionally applied medication-specific matching checks for ingredient, strength, route or dose form, brand or generic equivalence, and fixed-dose combination products so that similar drug names or incorrect strengths were not accepted solely on text overlap. Codes absent from the relevant terminology table were marked as “not_found”.

Check B - *Description consistency check:* It compare existing codes with their official terminology descriptions using a local thresholded text-overlap score (0.34 for direct single codes and 0.25 for expanded child codes). An existing direct code whose description scored below the threshold was marked “review_required” rather than automatically rejected, because a weak description match alone does not prove that the code is clinically incorrect.

Check C - *Controlled terminology expansion:* it approved parent categories, prefixes, ranges, and wildcard patterns into valid child or member codes using the local terminology tables. Because expanded child codes are often more specific than the broad clinical description supplied by the LLM, valid descendants of a verified family, range, or wildcard pattern were allowed to pass with warnings.

Final acceptance rules were then applied to decide which verified items entered the exact-code output. Codes placed by the generator under non-primary headings such as ‘related’ or ‘reminder’ were excluded from the accepted set, as were non-billable ICD parent or category codes when more specific exact codes were required. These excluded items were retained in the detailed verification output for auditability but were not counted as accepted query-ready codes. The complete verifier rules, thresholds, expansion limits, and final acceptance logic are detailed in the Supplementary Materials.

The verifier also expanded supported parent categories, prefixes, ranges, and wildcard patterns into valid child codes using the local terminology tables. Expansion was performed deterministically from the database rather than by the LLM, reducing the risk of invented or miscounted child codes. Safety limits were used to prevent overly broad automatic expansion.

Plain parent stems expanding to large child sets were held for review, whereas explicit wildcard patterns were expanded up to a higher cap. The Family expansion condition therefore relied on explicit patterns for broad families. The verifier also preserved vocabulary distinctions, routing each code to the table for its own system. For laboratory criteria, the system mapped the codable laboratory test concept to LOINC while treating numeric thresholds, units, timing windows, and comparison operators as non-code logic. For example, a criterion involving eGFR by the CKD-EPI formula would require a LOINC code for the laboratory measurement, while the numeric threshold would be applied separately to observed laboratory results.

### 3.7 Implementation

The system was implemented as a lightweight Python application using local terminology tables for verification, expansion, and retrieval, including RxNorm files for medication concept lookup. The application supported multiple LLM providers for the generation and formatting steps.

Output files included detailed verification results and comparison-ready accepted code sets. Output logic was shared across all conditions. Generation and refinement were run with provider reasoning enabled, with high reasoning effort. Because reasoning mode does not permit a fixed zero temperature, model outputs were not fully deterministic across runs.

## 4 Evaluation

### 4.1 Reference standard

Evaluating code-set construction requires a criterion-level reference standard, consisting of codes that correctly operationalize each criterion, rather than encounter-level assignments such as MIMICs billed codes. We used the expert-built code sets published by the RCT-DUPLICATE trial-replication project. No reference set was LLM-generated, avoiding grading the models against their own outputs. After excluding two underspecified criteria for which reviewer agreement was not reached, the evaluation comprised 40 clinical criteria across 11 trial groups. The automated verifier-impact dataset for these criteria contained 51,898 unique verifier rows across the evaluated pipeline-model combinations, including 11,183 single-code rows and 40,715 family, member, wildcard, or range-derived rows, reflecting the code-level verification workload required to operationalize trial criteria. This exceeds the 45,095 post-expansion code pairs in Table 3 because those counts reflect pair-level results after expansion, filtering, and deduplication.

The reference standard was derived from the RCT Duplicate target trial emulation project^30^, which emulated 32 randomized trials using real-world data. For each selected clinical criterion, we used the corresponding code set generated and used in that project as the ground-truth reference. These reference code sets represented the codes used to operationalize trial eligibility criteria, outcomes, procedures, laboratory concepts, and clinical conditions in structured real-world data. The complete clinical criteria, assigned complexity tiers, and reference code sets are provided in Supplementary Table S1.

Reference code sets were organized by vocabulary, including ICD-9-CM diagnosis codes, ICD-9 procedure codes (labeled ICD-9-PCS in the system), ICD-10-CM diagnosis codes, ICD-10-PCS procedure codes, and LOINC when applicable. CPT-4 codes present in some reference sets were excluded from all metric calculations, as the agent’s supported vocabularies do not include CPT. When reference concepts were represented by parent categories, stems, or grouped expressions, they were converted into explicit code lists before scoring. A human domain expert manually reviewed and curated the reference code sets used for evaluation. In addition, because some reference sets required expansion from parent categories, stems, or grouped expressions into explicit exact-code lists, the expanded code lists generated by the Python preprocessing code were subject to manual quality control. The expert manually annotated 10 clinical criteria to verify the reference-standard construction process and randomly reviewed 20% of the final exact-code reference sets to confirm that expansion and deduplication produced clinically appropriate codes. Any identified issues were corrected before metric calculation. Comparison was performed on final valid codes and non-billable parent/category ICD codes were excluded where applicable.

To further evaluate reference-standard suitability, three human reviewers (S.A., D.H, and K.P.) independently reviewed a targeted subset of code sets selected for ambiguity, high error counts, or potential mismatch between the trial phrase and the operational code list. The review produced 31 reviewer-code-set assessments across 22 unique criteria and code sets. Five code sets had overlapping review by at least two reviewers, including four reviewed by all three reviewers. Reviewers assessed whether the existing reference set was fully supported by the criterion, whether codes were missing, and whether the criterion was sufficiently specific for exact-code evaluation. They also provided notes and proposed replacement code lists when appropriate. Disagreements were adjudicated before final scoring.

### 4.2 Criterion complexity tiers

To evaluate performance across different levels of input complexity, each criterion was assigned to one of three tiers. Tier 1 included simple criteria without explicit logical connectors and without a requirement to combine diagnosis and procedure information. These criteria generally described a single clinical concept, such as a disease, prior condition, procedure, or laboratory concept.

Tier 2 included criteria with one source of added complexity. This included criteria containing logical connectors such as “and” or “or,” or criteria requiring codes from more than one clinical domain, such as both diagnosis and procedure vocabularies. These criteria required the agent to preserve relationships among concepts or recognize that the same eligibility criterion could be represented through multiple code types.

Tier 3 included the most complex criteria, defined as criteria that contained both logical structure and multiple clinical code domains. For example, these criteria could require the combination of diagnosis and procedure codes while also involving logical relationships among sub-criteria. This tier was intended to test whether the agent could handle both semantic logic and multi-vocabulary code-set construction within the same input.

### 4.3 Experimental comparison

The primary comparison was among the non-retrieval, retrieval-augmented, and Family expansion pipelines. All three conditions used the same input criteria, formatting step, terminology-verification layer, and reference standard. Differences in performance were therefore interpreted as reflecting the generation step, retrieval augmentation or family selection, rather than downstream verification or output.

Each condition differed only in the generation step (Sections 3.3–3.5) such that inputs, formatting, verification, and scoring were identical. In the Family expansion condition, the first call used the family-selection prompt (Section 3.5) to produce expandable patterns rather than an enumerated answer, and the result was passed unchanged to the same formatter and verifier.

### 4.4 Metrics

The accepted output from each agent run was compared with the corresponding reference code set using exact-code matching. For medication concepts, the exact identifier was the RxNorm RxCUI. A generated code was counted as a true positive if it exactly matched a reference-standard code in the same vocabulary. A generated code absent from the reference standard was counted as a false positive. A reference-standard code not generated by the agent was counted as a false negative. Complete criterion-level precision, recall, and F1 results for every pipeline-model combination are reported in Supplementary Table S2.

Precision was calculated as true positives divided by all verified accepted codes (the post-verification output). Recall was calculated as true positives divided by all reference-standard codes. F1 score was calculated as the harmonic mean of precision and recall. Metrics were calculated overall across all criteria and separately within each criterion-complexity tier. To quantify the independent contribution of terminology verification, we also conducted a staged verifier ablation in which exact-code performance was calculated after formatting but before verification, after code-existence and vocabulary checks, after acceptance filtering without expansion, and after final deterministic expansion. This allowed us to assess both aggregate performance and whether performance changed as criteria became more logically complex or required multiple coding domains.

As an exploratory secondary analysis, mixed-effects binomial models assessed differences by LLM, pipeline, and criterion-complexity tier. Recall modeled TP as successes and FN as failures; precision modeled TP as successes and FP as failures, with fixed effects for LLM, pipeline, tier, and their interactions and random intercepts for criteria nested within trial. Because Tier 3 included only four criteria, analyses were considered exploratory and are reported in Supplementary Table S3.

## 5 Results

The evaluation included 40 clinical criteria across three pipeline conditions and four LLM configurations. Performance differed substantially across pipelines and models, with the strongest overall performance observed for the Baseline pipeline when paired with Claude. Across all criteria, Claude with Baseline achieved precision of 0.755, recall of 0.619, and F1 of 0.680. The Baseline GPT-5.5 configuration produced the next strongest overall result, with precision of 0.569, recall of 0.658, and F1 of 0.610. Qwen performed best under Hybrid biomedical RAG, reaching precision of 0.656, recall of 0.470, and F1 of 0.548. MedGemma produced lower recall than the hosted model configurations across most conditions, with its highest F1 under Family expansion (precision 0.487, recall 0.316, F1 0.383). Table 1 and Figure 2 summarize the overall model-by-pipeline comparison.

**Figure 2.**
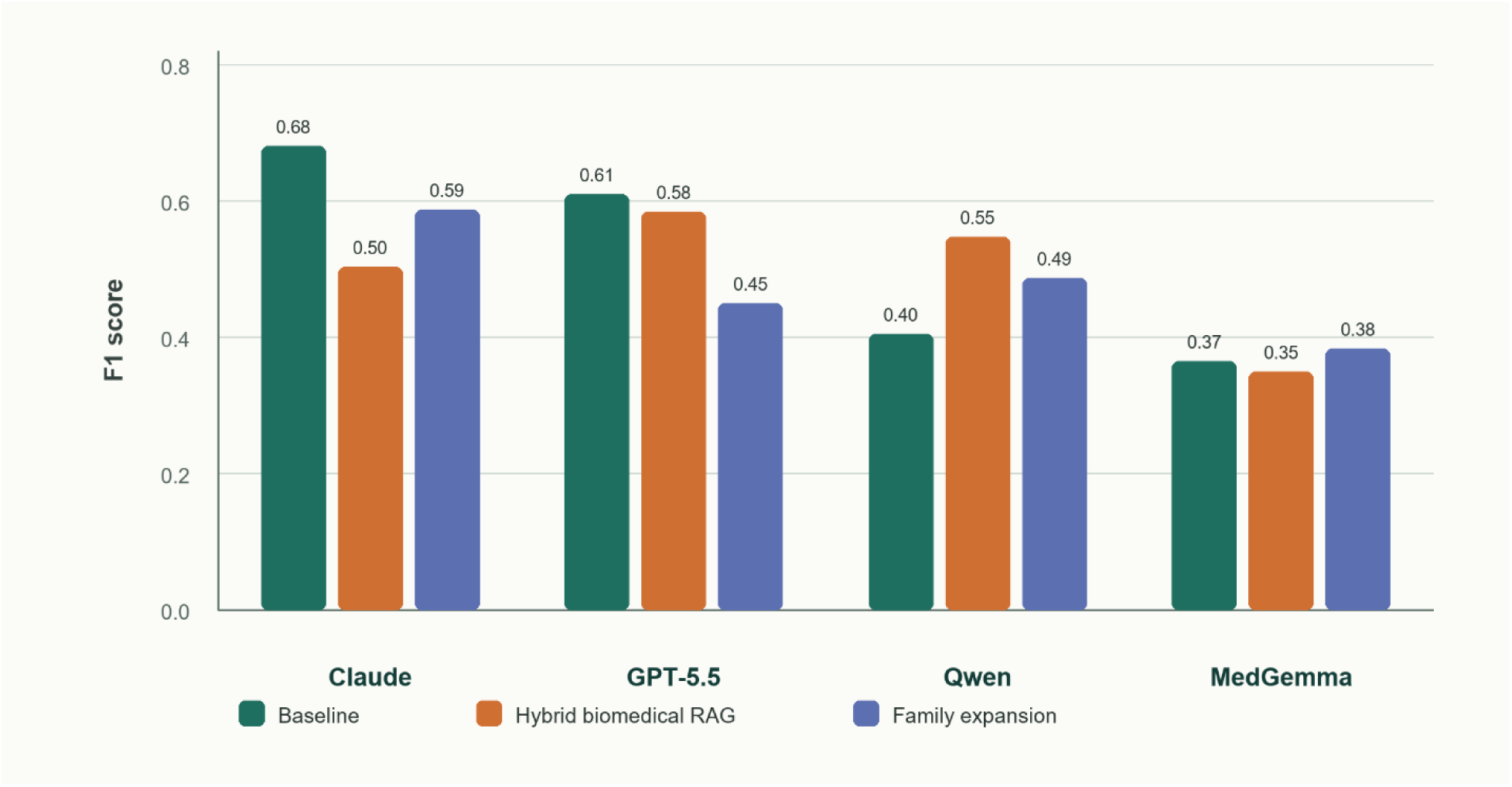
Overall F1 score by pipeline and model.

**Table 1.** Overall exact-code performance by pipeline and model with F1 confidence intervals.

| Pipeline | Model | Rows with output | Correct | Wrong | Missed | Precision | Recall | Micro F1, 95% CI | Macro F1, 95% CI |
| --- | --- | --- | --- | --- | --- | --- | --- | --- | --- |
| Baseline | Claude | 38 | 1421 | 460 | 875 | 0.755 | 0.619 | 0.680<br>[0.479-0.831] | 0.649<br>[0.542-0.749] |
| Baseline | GPT-5.5 | 40 | 1510 | 1145 | 786 | 0.569 | 0.658 | 0.610<br>[0.429-0.781] | 0.672<br>[0.581-0.759] |
| Baseline | Qwen | 36 | 717 | 529 | 1579 | 0.575 | 0.312 | 0.405<br>[0.288-0.546] | 0.483<br>[0.374-0.588] |
| Baseline | MedGemma | 35 | 573 | 268 | 1723 | 0.681 | 0.250 | 0.365<br>[0.226-0.479] | 0.293<br>[0.200-0.392] |
| Hybrid biomedical RAG | Claude | 39 | 907 | 405 | 1389 | 0.691 | 0.395 | 0.503<br>[0.360-0.668] | 0.652<br>[0.541-0.755] |
| Hybrid biomedical RAG | GPT-5.5 | 40 | 1465 | 1256 | 831 | 0.538 | 0.638 | 0.584<br>[0.434-0.760] | 0.764<br>[0.683-0.841] |
| Hybrid biomedical RAG | Qwen | 40 | 1080 | 567 | 1216 | 0.656 | 0.470 | 0.548<br>[0.413-0.682] | 0.625<br>[0.546-0.705] |
| Hybrid biomedical RAG | MedGemma | 39 | 568 | 383 | 1728 | 0.597 | 0.247 | 0.350<br>[0.190-0.529] | 0.366<br>[0.272-0.464] |
| **Family expansion** | Claude | 39 | 1557 | 1453 | 739 | 0.517 | 0.678 | 0.587<br>[0.463-0.715] | 0.618<br>[0.527-0.709] |
| **Family expansion** | GPT-5.5 | 40 | 1634 | 3328 | 662 | 0.329 | 0.712 | 0.450<br>[0.313-0.603] | 0.458<br>[0.379-0.538] |
| **Family expansion** | Qwen | 36 | 1274 | 1661 | 1022 | 0.434 | 0.555 | 0.487<br>[0.298-0.693] | 0.485<br>[0.376-0.589] |
| **Family expansion** | MedGemma | 32 | 726 | 766 | 1570 | 0.487 | 0.316 | 0.383<br>[0.216-0.516] | 0.273<br>[0.192-0.358] |

Note. Micro metrics aggregate exact code pairs across criteria. Macro F1 averages criterion-level F1. Confidence intervals are 95% percentile bootstrap intervals from criterion-level resampling.

Targeted human review identified ambiguity in several trial-derived phrases. Across 31 assessments of 22 unique criteria and code sets, two underspecified inputs were excluded and two reference sets were revised after adjudication before metrics were recalculated.

The Hybrid biomedical RAG pipeline did not uniformly improve performance. For Qwen, retrieval improved overall F1 compared with Baseline (0.548 versus 0.405). For GPT-5.5, retrieval produced a modest overall F1 decrease compared with Baseline (0.584 versus 0.610), although it improved performance in the most complex tier. For Claude, Hybrid biomedical RAG reduced recall and overall F1 compared with Baseline. For MedGemma, retrieval produced slightly lower F1 than Baseline (0.350 versus 0.365), mainly because recall remained low despite moderate precision.

Vocabulary-stratified results showed that the clearest retrieval benefit occurred for RxNorm medication mapping: GPT-5.5 Hybrid biomedical RAG achieved precision 0.972, recall 0.854, and F1 0.909 for RxNorm, compared with F1 0.320 for GPT-5.5 Baseline (Supplementary Table S8).

Macro-averaged results partly changed this interpretation: GPT-5.5 Hybrid biomedical RAG achieved higher macro-F1 than GPT-5.5 Baseline despite lower micro-F1, suggesting that retrieval helped some individual criteria even though it did not improve aggregate code-pair performance.

The family-expansion pipeline showed the expected recall-oriented behavior. It increased recall for all four model configurations relative to their Baseline settings, but the broader generated families also introduced more false-positive codes, lowering precision. For GPT-5.5, Family expansion achieved the highest overall recall among GPT-5.5 conditions (0.712) but also the lowest precision (0.329). For Claude, Family expansion achieved recall of 0.678 but lower precision than Baseline. For Qwen, Family expansion improved F1 compared with Qwen Baseline (0.487 versus 0.405) but remained below Qwen Hybrid biomedical RAG. For MedGemma, Family expansion produced its highest F1 (0.383) by improving recall relative to Baseline. These findings indicate that shifting the generation task toward family selection can recover additional reference codes, but it requires stricter control of overly broad expansions.

### 5.1 Performance by Criterion Complexity

Tier-stratified results showed that pipeline behavior depended on criterion complexity. For Claude, Baseline was strongest across all tiers, with F1 values of 0.638, 0.815, and 0.577 for Tiers 1, 2, and 3, respectively. For GPT-5.5, Baseline was strongest in Tier 2, while Hybrid biomedical RAG was strongest in Tier 3, where F1 increased to 0.607. For Qwen, Hybrid biomedical RAG was strongest in Tiers 1 and 3, while Family expansion was strongest in Tier 2. This suggests that retrieval evidence and family-selection logic may help selected model-criterion combinations, but their benefit is not uniform across complexity tiers.

MedGemma showed lower performance across all tiers. Baseline had the best MedGemma F1 in Tier 2 (0.482), and Family expansion was strongest for MedGemma in Tier 3 (0.374). Hybrid biomedical RAG improved MedGemma Tier 1 performance but remained weak in Tiers 2 and 3. This pattern suggests that local model performance was limited primarily by generation completeness rather than by downstream terminology verification alone.

Exploratory mixed-effects interaction analysis supported tier-dependent pipeline behavior for recall, with a significant LLM-by-pipeline-by-tier interaction (F = 2.37, p = 0.006). The corresponding interaction for precision was not statistically significant at the 0.05 level (F = 1.65, p = 0.077), consistent with the descriptive finding that pipeline effects differed more clearly for recall than for precision across tiers (Supplementary Table S3).

**Table 2.** F1 score by criterion-complexity tier.

| Pipeline | Model | Tier 1 F1 | Tier 2 F1 | Tier 3 F1 |
| --- | --- | --- | --- | --- |
| Baseline | Claude | 0.638 | 0.815 | 0.577 |
| Baseline | GPT-5.5 | 0.571 | 0.785 | 0.489 |
| Baseline | Qwen | 0.440 | 0.352 | 0.409 |
| Baseline | MedGemma | 0.280 | 0.482 | 0.334 |
| Hybrid biomedical RAG | Claude | 0.564 | 0.392 | 0.524 |
| Hybrid biomedical RAG | GPT-5.5 | 0.575 | 0.575 | 0.607 |
| Hybrid biomedical RAG | Qwen | 0.542 | 0.549 | 0.554 |
| Hybrid biomedical RAG | MedGemma | 0.530 | 0.235 | 0.134 |
| Family expansion | Claude | 0.563 | 0.665 | 0.541 |
| Family expansion | GPT-5.5 | 0.367 | 0.692 | 0.383 |
| Family expansion | Qwen | 0.374 | 0.622 | 0.576 |
| Family expansion | MedGemma | 0.305 | 0.490 | 0.374 |

### 5.2 Verifier Operational Impact

Automated auditing quantified three verifier functions that did not require human adjudication: database existence and vocabulary validation, filtering of LLM-labeled non-primary suggestions, and deterministic expansion of code families and patterns. The deterministic verifier checks and acceptance rules are summarized in Supplementary Table S5. Across the 12 pipeline-model combinations in the active 40-criterion evaluation set, the verifier classified 45,095 distinct post-expansion code pairs. It prevented 2,747 absent or wrong-vocabulary code pairs from entering accepted outputs including 897 with the Baseline pipeline, 1,124 with Hybrid biomedical RAG, and 726 with Family expansion.

The magnitude of nonexistent-code control varied substantially by model. MedGemma with Hybrid biomedical RAG had the largest number of absent or wrong-vocabulary codes blocked (938; 26.8%), followed by MedGemma with the Baseline pipeline (742; 27.6%). Claude produced the fewest such blocked codes in every pipeline (15 with Baseline, eight with Hybrid biomedical RAG, and 11 with Family expansion). Output-label filtering excluded 2,203 non-primary code pairs that the LLM had placed under headings such as related or reminder. The largest such effects occurred for GPT-5.5 with Hybrid biomedical RAG (680; 17.2%) and Claude with Family expansion (521; 10.3%). Table 3 reports these effects for each pipeline-model combination, with detailed verifier-impact counts provided in Supplementary Table S6.

Controlled expansion evaluated 2,499 family, prefix, range, or wildcard expressions. Baseline evaluated 423 expressions, Hybrid biomedical RAG evaluated 630, and Family expansion evaluated 1,446. After pair-level deduplication and final acceptance rules, the corresponding numbers of retained expanded code pairs, summed across model-specific runs, were 6,309, 6,337, and 20,817, respectively. Controlled-expansion volumes are provided in Supplementary Table S7.

**Table 3.** Automated verifier impact by pipeline and model.

| Pipeline | Model | Absent/wrong-vocabulary blocked, n (%) | Non-primary filtered, n (%) | Expressions evaluated | Expanded pairs retained | Post-expansion pairs |
| --- | --- | --- | --- | --- | --- | --- |
| **Baseline** | Claude | 15 (0.6%) | 134 (5.3%) | 118 | 1,669 | 2,519 |
| **Baseline** | GPT-5.5 | 21 (0.6%) | 6 (0.2%) | 100 | 1,999 | 3,681 |
| **Baseline** | Qwen | 119 (6.9%) | 53 (3.1%) | 95 | 912 | 1,731 |
| **Baseline** | MedGemma | 742 (27.6%) | 22 (0.8%) | 110 | 1,729 | 2,688 |
| **Hybrid biomedical RAG** | Claude | 8 (0.4%) | 163 (9.0%) | 140 | 1,371 | 1,810 |
| **Hybrid biomedical RAG** | GPT-5.5 | 37 (0.9%) | 680 (17.2%) | 172 | 1,939 | 3,958 |
| **Hybrid biomedical RAG** | Qwen | 141 (6.6%) | 15 (0.7%) | 136 | 1,330 | 2,151 |
| **Hybrid biomedical RAG** | MedGemma | 938 (26.8%) | 278 (7.9%) | 182 | 1,697 | 3,498 |
| **Family expansion** | Claude | 11 (0.2%) | 521 (10.3%) | 272 | 4,340 | 5,070 |
| **Family expansion** | GPT-5.5 | 39 (0.4%) | 102 (1.1%) | 575 | 8,583 | 8,989 |
| **Family expansion** | Qwen | 45 (0.9%) | 229 (4.6%) | 351 | 4,534 | 4,944 |
| **Family expansion** | MedGemma | 631 (15.6%) | 0 (0.0%) | 248 | 3,360 | 4,056 |

A staged verifier ablation directly tested whether terminology verification changed exact-code performance. Micro-averaged F1 increased from 0.254 before verification to 0.271 after existence/vocabulary checking, 0.260 after acceptance filtering without expansion, and 0.505 after final verification with deterministic expansion (Table 4). Existence and vocabulary checks removed 2,594 false-positive codes while removing 15 true-reference codes. Acceptance filtering removed 952 additional false positives but also removed 383 true-reference codes. The final expansion step added 8,843 true-reference codes and 10,533 false-positive codes. It also removed 1,843 false-positive pre-expansion items and 49 true-reference items. Thus, verification improved exact-code performance primarily through deterministic expansion, while filtering increased precision but reduced recall. Detailed stage-specific ablation results and code-removal/addition transition counts are provided in Supplementary Tables S10-S11.

**Table 4.** Staged verifier ablation, micro-averaged across all pipeline-model-criterion runs.

| Output stage | Precision | Recall | F1 | Delta F1 vs before verifier |
| --- | --- | --- | --- | --- |
| **Before verifier** | 0.416 | 0.183 | 0.254 | NA |
| **Existence/vocabulary check** | 0.528 | 0.182 | 0.271 | 0.017 |
| **Acceptance filtering** | 0.568 | 0.168 | 0.260 | 0.006 |
| **Final verifier + expansion** | 0.524 | 0.488 | 0.505 | 0.251 |

Stage-specific results showed that verifier gains were largest for pattern-heavy Family expansion outputs, while GPT-5.5 Baseline changed minimally after verification, indicating that deterministic verification contributed most when upstream outputs contained broad families, prefixes, or wildcard expressions (Supplementary Table S10).

### 5.3 Precision-Recall Tradeoffs

The main tradeoff across experimental conditions was between precision and recall. Baseline generally provided the best balance, especially for Claude and GPT. Family expansion increased the number of generated codes and improved recall in broad criteria, but it also increased false positives when expandable families included clinically adjacent or overly broad child codes. Retrieval was more conservative in some settings, improving precision for selected complex criteria but often reducing recall when the retrieved candidate set narrowed the final output too aggressively. Figure 3 visualizes these precision-recall tradeoffs across all model and pipeline combinations. Vocabulary-stratified performance and verifier input-form counts are provided in Supplementary Tables S8-S9.

**Figure 3.**
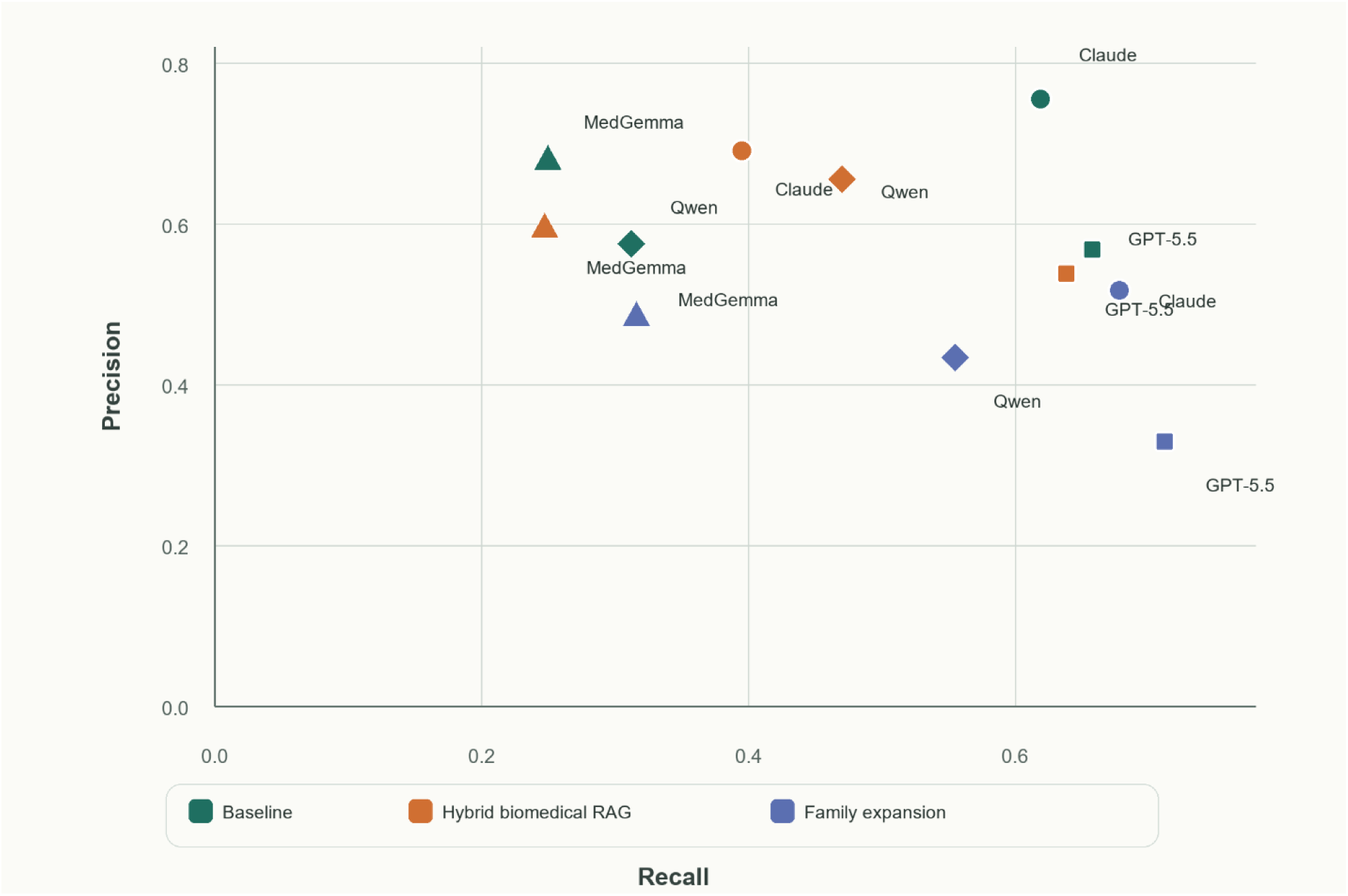
Precision-recall tradeoff by pipeline and model.

Taken together, the results suggest that terminology verification is useful for making outputs query-ready, but the largest remaining source of error is the upstream generation step. When the first LLM call omits relevant code families, verification cannot recover them unless they are introduced through retrieval or family-selection logic. Conversely, when the first LLM call proposes an overly broad family, deterministic expansion can increase recall while also producing false positives.

**Table 5.** Change in performance relative to the Baseline pipeline.

| Model | Comparison | Delta precision | Delta recall | Delta F1 |
| --- | --- | --- | --- | --- |
| Claude | Hybrid biomedical RAG vs Baseline | -0.064 | -0.224 | -0.178 |
| Claude | Family expansion vs Baseline | -0.238 | 0.059 | -0.094 |
| GPT-5.5 | Hybrid biomedical RAG vs Baseline | -0.030 | -0.020 | -0.026 |
| GPT-5.5 | Family expansion vs Baseline | -0.239 | 0.054 | -0.160 |
| Qwen | Hybrid biomedical RAG vs Baseline | 0.080 | 0.158 | 0.143 |
| Qwen | Family expansion vs Baseline | -0.141 | 0.243 | 0.082 |
| MedGemma | Hybrid biomedical RAG vs Baseline | -0.084 | -0.002 | -0.015 |
| MedGemma | Family expansion vs Baseline | -0.195 | 0.067 | 0.018 |

Relative to Baseline, Hybrid biomedical RAG reduced overall F1 for Claude, GPT-5.5, and MedGemma, but improved Qwen by 0.143 F1 points. Family expansion increased recall for all four model configurations, but reduced precision enough that overall F1 remained below Baseline for Claude and GPT-5.5. For Qwen, Family expansion improved F1 compared with Baseline (+0.082) but remained below Hybrid biomedical RAG, while for MedGemma it improved F1 compared with both Baseline and Hybrid biomedical RAG (0.383 versus 0.365 and 0.350).

## 6 Discussion

In this study, we developed TrialCode, an LLM-based agent for translating clinical trial eligibility criteria into computable code sets across multiple clinical vocabularies. Rather than relying on a single LLM, TrialCode employs a modular, generate, format–verify design that decomposes the workflow into sequential, coordinated subagents responsible for clinical interpretation, structured code-set generation and formatting, and deterministic terminology verification, separately. The results highlight both the importance of the underlying intelligence and reasoning capabilities of the base model and the potential for substantially enhancing open-source LLMs through targeted context-engineering strategies. In particular, approaches such as Hybrid biomedical RAG and terminology-guided family expansion can provide domain-specific knowledge and structured vocabulary context that may not be sufficiently represented in the base model itself.

The Hybrid biomedical RAG condition produced mixed results. Retrieval improved Qwen overall and helped GPT-5.5 on the most complex criteria, where additional terminology evidence appeared to support broader coverage of multi-concept inputs. However, retrieval did not improve overall performance for Claude, GPT-5.5, or MedGemma. This suggests that retrieval should not be treated as automatically beneficial. In this task, retrieval can help when it supplies missing candidate families, but it can also narrow the model’s answer or introduce distracting candidates when the retrieved evidence is incomplete or poorly aligned with the criterion.

The family-expansion condition highlighted a key design tradeoff. Broad trial criteria often require many child codes, and model-based enumeration of complete exact-code sets was unreliable in the tested configurations. Family expansion harnessed existing expert-curated knowledge systems (i.e., clinical terminologies and ontologies) that can improve recall for some general-purpose models. However, broad prefixes also introduced clinically adjacent or unintended child codes, reducing precision and sometimes F1. These findings suggest that family expansion is useful only when constrained by explicit family-selection rules, vocabulary-specific expansion logic, and clinical review for high-impact criteria.

The model comparison also has practical implications. Claude was the strongest and most stable model with Baseline, while GPT-5.5 performed competitively and showed the clearest Tier 3 benefit from Hybrid biomedical RAG. Qwen showed a different pattern in that its aggregate performance improved with Hybrid biomedical RAG, suggesting that retrieval evidence may be particularly useful for this hosted open-weight reasoning model. MedGemma had substantially lower recall overall. Family expansion modestly improved its F1, but these results should be interpreted as specific to the local MedGemma deployment evaluated here rather than as a general conclusion about all open-weight models. This configuration may require task-specific prompting, fine-tuning, or stronger candidate-generation support before matching the stronger hosted systems for this exact-code construction task.

The verifier should be interpreted as a terminology-control layer rather than a complete clinical adjudicator. It can validate code existence, check vocabulary membership, expand explicit patterns, remove non-final parent categories, and standardize accepted outputs. For RxNorm, this includes checking proposed RxCUIs against local concept names, while preserving strength, route, dose-form, brand or generic, and combination-product distinctions as medication-specific matching constraints. It cannot infer codes that were never proposed upstream, and it cannot fully decide whether a clinically valid code is appropriate for a particular trial criterion without additional clinical logic. This distinction is important because low recall often reflects incomplete generation, while low precision often reflects overly broad generation or expansion.

The verifier contributed differently across models and pipelines. Existence and vocabulary checks increased precision from 0.416 to 0.528 by removing invalid or wrong-vocabulary false positives, with the largest protection observed for MedGemma and the smallest for Claude. Acceptance filtering further increased average precision to 0.568 but reduced recall because some true-reference codes were also removed. The final verifier stage increased recall through deterministic expansion, raising micro-F1 from 0.254 before verification to 0.505 after full verification and expansion. This supports the benefits of dissecting the verification subagent into more granular modules, while also showing that expansion can introduce false positives especially when upstream families are already too broad.

Controlled expansion also translated broad family-level expressions into explicit, database-queryable codes. However, database validity is not equivalent to clinical relevance: every expanded child may be valid within its source family and ontology while still being too broad for the trial criterion. The verifier should therefore be viewed as a reproducible terminology safeguard that controls nonexistent codes and produces query-ready expansions, not as a substitute for clinical adjudication.

Another important source of lower exact-code metrics is the nature of the input itself. The agent was not given the entire referential terminology databases. It was given clinical phrases derived from trial-emulation criteria. These phrases often require interpretation of the trial context, intended data source, and operational definition. As a result, an output can be clinically plausible and data source adaptive while still differing from the reference set, because the reference set reflects one specific operationalization used in the RCT-DUPLICATE emulation rather than the only possible valid code set. This issue is especially relevant for broad criteria such as intracranial bleeding, cardiovascular disease, prior bypass surgery, or gastrointestinal bleeding, where choices about history codes, sequela codes, traumatic variants, procedure codes, status codes, or data-source proxies can materially change the final code set.

Targeted human review showed that some trial-derived criteria were too broad or underspecified to support a stable exact-code reference set without adjudication. Two inputs were removed because reviewers did not converge on a sufficiently specific operational definition, and two reference sets were revised after review. These manual adjudications showed that exact-code performance depended not only on model behavior, but also on whether the clinical criterion contained enough context to define a reproducible computable phenotype. For example, “intracranial bleeding” lacked sufficient context to determine whether traumatic, neonatal/perinatal, sequela, or history codes should be included, requiring adjudication of the final reference set.

Several limitations should be considered. The evaluation used a finite set of trial-derived criteria and reference code sets, so results may differ for other therapeutic areas, coding systems, medication vocabularies, or local data models. The reference standard reflects the operational choices used in the target trial emulation source, and alternative valid code-set definitions may exist for some criteria. The evaluation also measured exact-code agreement rather than downstream cohort retrieval, so future work should assess whether code-set differences materially change patient counts, phenotype validity, and treatment-effect estimates. Finally, LLM outputs can vary across runs and model versions; production use should include saved prompts, model identifiers, outputs, and deterministic post-processing logs.

## 7 Conclusion

Overall, we have shown that the TrialCode agent provides a useful, modular, prototypical architecture (i.e., generate-format-verify) for translating narrative selection criteria to computable phenotypes that can be directly processed by existing clinical data query systems. We have also shown that effectively incorporating relevant biomedical knowledge and terminology structure into the model’s reasoning context can enable open-source models to achieve performance comparable to that of proprietary models at substantially lower cost. Future improvements should focus on better upstream family selection, safer retrieval candidate ranking, and explicit handling of criteria that require data-model decisions beyond diagnosis, procedure, laboratory, and medication code mapping.

## Data Availability

The reference code sets used in this study are provided in the Supplementary Materials. Source code, prompts, verifier rules, and evaluation scripts for TrialCode Agent will be made publicly available in a GitHub repository upon publication.

## Declaration of generative AI and AI-assisted technologies in the manuscript preparation process

During the preparation of this work, the authors used OpenAI ChatGPT to assist in creating the workflow illustration in Figure 1. The authors subsequently reviewed and edited the figure and take full responsibility for its accuracy and content.

## References

1. Danaei G, Rodríguez LAG, Cantero OF, Logan RW, Hernán MA. Electronic medical records can be used to emulate target trials of sustained treatment strategies. J Clin Epidemiol. 2018;96:12–22. doi:10.1016/j.jclinepi.2017.11.021

2. Habibdoust A, Zuo H, Koopman RJ, Gupta A, Mazzotti DR, Song X. Target trial emulation in hypertension research: a scoping review of current applications and methodological practices. J Hypertens. 2026;44(1):37–48. doi:10.1097/HJH.0000000000004188

3. Hernán MA, Robins JM. Using Big Data to Emulate a Target Trial When a Randomized Trial Is Not Available. Am J Epidemiol. 2016;183(8):758–764. doi:10.1093/aje/kwv254

4. Habibdoust A, Song X. TrialCalibre: A Fully Automated Causal Engine for RCT Benchmarking and Observational Trial Calibration. In: ICML 2025 Workshop on Scaling Up Intervention Models; 2025. Accessed February 9, 2026. https://openreview.net/forum?id=r2DGtDFRrq

5. Kim H, Kim M, Kim S, You SC. From study design to executable code: automating target trial emulation with large language models. Jamia Open. 2026;9(4):ooag131. doi:10.1093/jamiaopen/ooag131

6. Li H, Pan W, Rajendran S, Zang C, Wang F. EmulatRx: Empowering Clinical Trial Design with Agentic Intelligence and Real World Data. medRxiv. Preprint posted online March 3, 2026:2025.04.17.25326033. doi:10.1101/2025.04.17.25326033

7. Zhang J, He H, Ma L, et al. ConceptPsy: A comprehensive benchmark suite for hierarchical psychological concept understanding in LLMs. Neurocomputing. 2025;637:130070. doi:10.1016/j.neucom.2025.130070

8. Motzfeldt AG, Edin J, Christensen CL, Hardmeier C, Maaløe L, Rogers A. Code Like Humans: A Multi-Agent Solution for Medical Coding. In: Christodoulopoulos C, Chakraborty T, Rose C, Peng V, eds. Findings of the Association for Computational Linguistics: EMNLP 2025. Association for Computational Linguistics; 2025:22612–22627. doi:10.18653/v1/2025.findings-emnlp.1231

9. Lee KH, Jang S, Kim GJ, et al. Large Language Models for Automating Clinical Trial Criteria Conversion to Observational Medical Outcomes Partnership Common Data Model Queries: Validation and Evaluation Study. JMIR Med Inform. 2025;13:e71252. doi:10.2196/71252

10. Lee K, Mai Y, Liu Z, et al. CriteriaMapper: establishing the automatic identification of clinical trial cohorts from electronic health records by matching normalized eligibility criteria and patient clinical characteristics. Sci Rep. 2024;14(1):25387. doi:10.1038/s41598-024-77447-x

11. Soroush A, Glicksberg BS, Zimlichman E, et al. Large Language Models Are Poor Medical Coders — Benchmarking of Medical Code Querying. NEJM AI. 2024;1(5):AIdbp2300040. doi:10.1056/AIdbp2300040

12. Hou Z, Liu H, Bian J, He X, Zhuang Y. Enhancing medical coding efficiency through domain-specific fine-tuned large language models. npj Health Syst. 2025;2(1):14. doi:10.1038/s44401-025-00018-3

13. Dobbins NJ, Han B, Zhou W, et al. LeafAI: query generator for clinical cohort discovery rivaling a human programmer. J Am Med Inform Assoc. 2023;30(12):1954–1964. doi:10.1093/jamia/ocad149

14. Yang L, Han Y, Liu L, et al. EC2Seq2Sql: Patient-trial matching with LLM agents. PLOS ONE. 2026;21(2):e0341827. doi:10.1371/journal.pone.0341827

15. Anjos De Almeida V, De Camargo V, Gómez-Bravo R, et al. Large language models as medical code selectors: a benchmark using the International Classification of Primary Care. JAMIA Open. 2026;9(1):ooag017. doi:10.1093/jamiaopen/ooag017

16. Huang CW, Tsai SC, Chen YN. PLM-ICD: Automatic ICD Coding with Pretrained Language Models. In: Proceedings of the 4th Clinical Natural Language Processing Workshop. Association for Computational Linguistics; 2022:10–20. doi:10.18653/v1/2022.clinicalnlp-1.2

17. Kirby JC, Speltz P, Rasmussen LV, et al. PheKB: a catalog and workflow for creating electronic phenotype algorithms for transportability. J Am Med Inform Assoc. 2016;23(6):1046–1052. doi:10.1093/jamia/ocv202

18. Callies A, Bodinier Q, Ravaud P, Davarpanah K. Real-world validation of a multimodal LLM-powered pipeline for high-accuracy clinical trial patient matching. Commun Med. 2025;5(1):536. doi:10.1038/s43856-025-01256-0

19. Chen H, Li X, He X, et al. Enhancing Patient-Trial Matching With Large Language Models: A Scoping Review of Emerging Applications and Approaches. JCO Clin Cancer Inform. 2025;9:e2500071. doi:10.1200/CCI-25-00071

20. National Center For Health Statistics. International Classification of Diseases, Ninth Revision, Clinical Modification (ICD-9-CM). Centers for Disease Control and Prevention. Accessed June 22, 2026. https://www.cdc.gov/nchs/icd/icd9cm.htm

21. Centers for Medicare & Medicaid Services. ICD-9-CM Volume 3 Procedures. Centers for Medicare & Medicaid Services. Accessed June 22, 2026. https://www.cms.gov/medicare/coding-billing/icd-9-codes

22. Centers for Medicare & Medicaid Services. 2026 ICD-10-PCS Code Files. Centers for Medicare & Medicaid Services. 2026. Accessed June 22, 2026. https://www.cms.gov/medicare/coding-billing/icd-10-codes

23. Regenstrief Institute, Inc. Logical Observation Identifiers Names and Codes (LOINC). Published online March 4, 2026. Accessed June 22, 2026. https://loinc.org/downloads/loinc/

24. National Library of Medicine. RxNorm. National Library of Medicine. Accessed June 22, 2026.. https://www.nlm.nih.gov/research/umls/rxnorm/

25. Robertson S, Zaragoza H. The Probabilistic Relevance Framework: BM25 and Beyond. Foundations and Trends® in Information Retrieval. 2009;4(1-2):1–174. doi:10.1561/1500000019

26. Gu Y, Tinn R, Cheng H, et al. Domain-Specific Language Model Pretraining for Biomedical Natural Language Processing. ACM Trans Comput Healthcare. 2022;3(1):1–23. doi:10.1145/3458754

27. Reimers N, Gurevych I. Sentence-BERT: Sentence Embeddings using Siamese BERT-Networks. In: Proceedings of the 2019 Conference on Empirical Methods in Natural Language Processing and the 9th International Joint Conference on Natural Language Processing (EMNLP-IJCNLP). Association for Computational Linguistics; 2019:3980–3990. doi:10.18653/v1/D19-1410

28. Deka P. S-PubMedBert-MS-MARCO. Hugging Face. Accessed June 22, 2026. https://huggingface.co/pritamdeka/S-PubMedBert-MS-MARCO

29. Cormack GV, Clarke CLA, Buettcher S. Reciprocal rank fusion outperforms condorcet and individual rank learning methods. In: Proceedings of the 32nd International ACM SIGIR Conference on Research and Development in Information Retrieval. ACM; 2009:758–759. doi:10.1145/1571941.1572114

30. Wang SV, Schneeweiss S, RCT-DUPLICATE Initiative, et al. Emulation of Randomized Clinical Trials With Nonrandomized Database Analyses: Results of 32 Clinical Trials. JAMA. 2023;329(16):1376. doi:10.1001/jama.2023.4221

