## Supplementary material for "TrialCode Agent: LLM-Assisted Clinical Code-Set Construction for Trial Emulation"

### Supplementary Materials

Hybrid LLM-Terminology Verification Pipeline for Clinical Code-Set Construction

This supplement reports the complete criterion-level reference standard, per-criterion performance, exploratory interaction analysis, deterministic verifier specification, and automated verifier-impact results. All values were generated from the final evaluation files after dropping two underspecified criteria and updating the adjudicated reference standards.

#### Supplementary Table S1. Clinical criteria, complexity tiers, and reference code sets

The evaluation included 40 distinct clinical criteria. CPT-4 codes were excluded from scoring because CPT was not a supported agent vocabulary; RxNorm RxCUIs were included for medication criteria after RxNorm support was added.

| ID | Trial | Tier | Clinical criterion | Reference code set |
|---|---|---|---|---|
| **C01** | DAPA-CKD(Kidney / cardio-renal) | 1 | CKD Stage II | ICD-9 diagnosis: 585.2<br>ICD-10 diagnosis: N18.2 |
| **C02** | DAPA-CKD(Kidney / cardio-renal) | 2 | CKD Stage II and CKD Stage IIIa&b and CKD Stage IV | ICD-9 diagnosis: 585.2<br>ICD-10 diagnosis: N18.2<br>ICD-9 diagnosis: 585.3<br>ICD-10 diagnosis: N18.30, N18.31, N18.32<br>ICD-9 diagnosis: 585.4<br>ICD-10 diagnosis: N18.4 |
| **C03** | DAPA-CKD(Kidney / cardio-renal) | 1 | Type I diabetes mellitus | Type I diabetes mellitus: ICD-9 diagnosis: 250.01, 250.03, 250.11, 250.13, 250.21, 250.23, 250.31, 250.33, 250.41, 250.43, 250.51, 250.53, 250.61, 250.63, 250.71, 250.73, 250.81, 250.83, 250.91, 250.93; ICD-10 diagnosis: E10.10, E10.11, E10.21, E10.22, E10.29, E10.311, E10.319, E10.3211, E10.3212, E10.3213, E10.3219, E10.3291, E10.3292, E10.3293, E10.3299, E10.3311, E10.3312, E10.3313, E10.3319, E10.3391, E10.3392, E10.3393, E10.3399, E10.3411, E10.3412, E10.3413, E10.3419, E10.3491, E10.3492, E10.3493, E10.3499, E10.3511, E10.3512, E10.3513, E10.3519, E10.3521, E10.3522, E10.3523, E10.3529, E10.3531, E10.3532, E10.3533, E10.3539, E10.3541, E10.3542, E10.3543, E10.3549, E10.3551, E10.3552, E10.3553, E10.3559, E10.3591, E10.3592, E10.3593, E10.3599, E10.36, E10.37X1, E10.37X2, E10.37X3, E10.37X9, E10.39, E10.40, E10.41, E10.42, E10.43, E10.44, E10.49, E10.51, E10.52, E10.59, E10.610, E10.618, E10.620, E10.621, E10.622, E10.628, E10.630, E10.638, E10.641, E10.649, E10.65, E10.69, E10.8, E10.9, E10.A0, E10.A1, E10.A2, E10.00, E10.01 |
| **C04** | DAPA-CKD(Kidney / cardio-renal) | 2 | Autosomal dominant or recessive PKD | ICD-9 diagnosis: 753.12, 753.13, 753.14, ICD-10 diagnosis: Q61.3, Q61.2, Q61.19, Q61.11 |

| | | | | |
|---|---|---|---|---|
| **C05** | DAPA-CKD(Kidney / cardio-renal) | 1 | Acute kidney injury (AKI) | ICD-9 diagnosis: 584.5, 584.6, 584.7, 584.8, 584.9, ICD-10 diagnosis: N17.0, N17.1, N17.2, N17.8, N17.9 |
| **C06** | DAPA-CKD(Kidney / cardio-renal) | 1 | Dapagliflozin 10 mg | RXNORM: 1488564,1488565,1488569,1534397,1486977 |
| **C07** | ONTARGET | 1 | essential hypertension | ICD 9:401.9, 401.0 ,401.1 , ICD 10 : I10 |
| **C08** | ONTARGET | 3 | Previous limb bypass surgery or angioplasty | Z95.820, Z95.828, Z98.62, V43.4, 39.25, 39.29, 041K09H, 041K09J, 041K09K, 041K09L, 041K09M, 041K09N, 041K09P, 041K09Q, 041K09S, 041K0AH, 041K0AJ, 041K0AK, 041K0AL, 041K0AM, 041K0AN, 041K0AP, 041K0AQ, 041K0AS, 041K0JH, 041K0JJ, 041K0JK, 041K0JL, 041K0JM, 041K0JN, 041K0JP, 041K0JQ, 041K0JS, 041K0KH, 041K0KJ, 041K0KK, 041K0KL, 041K0KM, 041K0KN, 041K0KP, 041K0KQ, 041K0KS, 041K0ZH, 041K0ZJ, 041K0ZK, 041K0ZL, 041K0ZM, 041K0ZN, 041K0ZP, 041K0ZQ, 041K0ZS, 041L09H, 041L09J, 041L09K, 041L09L, 041L09M, 041L09N, 041L09P, 041L09Q, 041L09S, 041L0AH, 041L0AJ, 041L0AK, 041L0AL, 041L0AM, 041L0AN, 041L0AP, 041L0AQ, 041L0AS, 041L0JH, 041L0JJ, 041L0JK, 041L0JL, 041L0JM, 041L0JN, 041L0JP, 041L0JQ, 041L0JS, 041L0KH, 041L0KJ, 041L0KK, 041L0KL, 041L0KM, 041L0KN, 041L0KP, 041L0KQ, 041L0KS, 041L0ZH, 041L0ZJ, 041L0ZK, 041L0ZL, 041L0ZM, 041L0ZN, 041L0ZP, 041L0ZQ, 041L0ZS, 041M09L, 041M09M, 041M09P, 041M09Q, 041M09S, 041M0AL, 041M0AM, 041M0AP, 041M0AQ, 041M0AS, 041M0JL, 041M0JM, 041M0JP, 041M0JQ, 041M0JS, 041M0KL, 041M0KM, 041M0KP, 041M0KQ, 041M0KS, 041M0ZL, 041M0ZM, 041M0ZP, 041M0ZQ, 041M0ZS, 041N09L, 041N09M, 041N09P, 041N09Q, 041N09S, 041N0AL, 041N0AM, 041N0AP, 041N0AQ, 041N0AS, 041N0JL, 041N0JM, 041N0JP, 041N0JQ, 041N0JS, 041N0KL, 041N0KM, 041N0KP, 041N0KQ, 041N0KS, 041N0ZL, 041N0ZM, 041N0ZP, 041N0ZQ, 041N0ZS, 39.50, 39.90, 00.55, 00.60, 047C3ZZ, 047C3DZ, 047C34Z, 047D3ZZ, 047D3DZ, 047D34Z, 047H3ZZ, 047H3DZ, 047H34Z, 047J3ZZ, 047J3DZ, 047J34Z, 047K3ZZ, 047K3DZ, 047K34Z, 047L3ZZ, 047L3DZ, 047L34Z, 047M3ZZ, 047M3DZ, 047M34Z, 047N3ZZ, 047N3DZ, 047N34Z, 047P3ZZ, 047P3DZ, 047P34Z, 047Q3ZZ, 047Q3DZ, 047Q34Z, 047R3ZZ, 047R3DZ, 047R34Z, 047S3ZZ, 047S3DZ, 047S34Z, 047T3ZZ, 047T3DZ, 047T34Z, 047U3ZZ, 047U3DZ, 047U34Z, 047V3ZZ, 047V3DZ, 047V34Z, 047W3ZZ, 047W3DZ, 047W34Z |
| **C09** | ONTARGET | 3 | Diabetes Mellitus (types I or II): with evidence of end-organ damage (retinopathy, Left ventricular hypertrophy, micro or macro albuminuria) | Diabetic retinopathy ICD-9 diagnosis: 362.01, 362.02, 362.03, 362.04, 362.05, 362.06, 362.07.<br>Diabetes with other ophthalmic manifestations ICD-9 diagnosis: 250.50, 250.51, 250.52, 250.53, 362.01, 362.02, 362.03, 362.04, 362.05, 362.06, 362.07<br>Diabetic nephropathy ICD-9 diagnosis:250.40, 250.41, 250.42, 250.43, 583.81<br>Diabetic neuropathy ICD-9 diagnosis:,, . ICD10: E10.311, E10.319, E10.3211, E10.3212, E10.3213, E10.3219, E10.3291, E10.3292, E10.3293, E10.3299, E10.3311, E10.3312, E10.3313, E10.3319, |

| | | | | |
|---|---|---|---|---|
| | | | | E10.3391, E10.3392, E10.3393, E10.3399, E10.3411, E10.3412, E10.3413, E10.3419, E10.3491, E10.3492, E10.3493, E10.3499, E10.3511, E10.3512, E10.3513, E10.3519, E10.3521, E10.3522, E10.3523, E10.3529, E10.3531, E10.3532, E10.3533, E10.3539, E10.3541, E10.3542, E10.3543, E10.3549, E10.3551, E10.3552, E10.3553, E10.3559, E10.3591, E10.3592, E10.3593, E10.3599, E10.37X1, E10.37X2, E10.37X3, E10.37X9, E11.311, E11.319, E11.3211, E11.3212, E11.3213, E11.3219, E11.3291, E11.3292, E11.3293, E11.3299, E11.3311, E11.3312, E11.3313, E11.3319, E11.3391, E11.3392, E11.3393, E11.3399, E11.3411, E11.3412, E11.3413, E11.3419, E11.3491, E11.3492, E11.3493, E11.3499, E11.3511, E11.3512, E11.3513, E11.3519, E11.3521, E11.3522, E11.3523, E11.3529, E11.3531, E11.3532, E11.3533, E11.3539, E11.3541, E11.3542, E11.3543, E11.3549, E11.3551, E11.3552, E11.3553, E11.3559, E11.3591, E11.3592, E11.3593, E11.3599, E11.37X1, E11.37X2, E11.37X3, E11.37X9, I51.7, R80.0, R80.1, R80.8, R80.9, E10.21, E10.22, E10.29, E11.21, E11.22, E11.29 |
| **C10** | ONTARGET | 1 | Lower extremity amputation | Lower extremity amputation<br>ICD-9 diagnosis: V49.70, V49.71, V49.72, V49.73, V49.74, V49.75, V49.76, V49.77<br>ICD-9 procedure: 84.10, 84.11, 84.12, 84.13, 84.14, 84.15, 84.16, 84.17, 84.18<br>ICD-10 diagnosis: Z89.511, Z89.512, Z89.519, Z89.611, Z89.612, Z89.619, Z89.431, Z89.432, Z89.411, Z89.412, Z89.611, Z89.612, Z89.419, Z89.421, Z89.422, Z89.429, Z89.439, Z89.441, Z89.442, Z89.449, Z89.521, Z89.522, Z89.529, Z89.621, Z89.622, Z89.629, ICD-10 procedure: 0Y6J0Z0, 0Y6K0Z0, 0Y6N0Z0, 0Y6M0Z0, 0Y6P0Z0, 0Y6L0Z0, 0Y6P0Z1, 0Y6L0Z1, 0Y6Q0Z0, 0Y6Q0Z1, 0Y6N0Z1, 0Y6R0Z0, 0Y6S0Z0, 0Y6Q0Z0, 0Y6T0Z0, 0Y620ZZ, 0Y630ZZ, 0Y640ZZ, 0Y670ZZ, 0Y680ZZ, 0Y6C0Z1, 0Y6C0Z2, 0Y6C0Z3, 0Y6D0Z1, 0Y6D0Z2, 0Y6D0Z3, 0Y6F0ZZ, 0Y6G0ZZ, 0Y6H0Z1, 0Y6H0Z2, 0Y6H0Z3, 0Y6J0Z1, 0Y6J0Z2, 0Y6J0Z3, 0Y6M0Z4, 0Y6M0Z5, 0Y6M0Z6, 0Y6M0Z7, 0Y6M0Z8, 0Y6M0Z9, 0Y6M0ZB, 0Y6M0ZC, 0Y6M0ZD, 0Y6M0ZF, 0Y6N0Z4, 0Y6N0Z5, 0Y6N0Z6, 0Y6N0Z7, 0Y6N0Z8, 0Y6N0Z9, 0Y6N0ZB, 0Y6N0ZC, 0Y6N0ZD, 0Y6N0ZF, 0Y6P0Z2, 0Y6P0Z3, 0Y6Q0Z2, 0Y6Q0Z3, 0Y6R0Z1, 0Y6R0Z2, 0Y6R0Z3, 0Y6S0Z1, 0Y6S0Z2, 0Y6S0Z3, 0Y6T0Z1, 0Y6T0Z2, 0Y6T0Z3, 0Y6U0Z0, 0Y6U0Z1, 0Y6U0Z2, 0Y6U0Z3, 0Y6V0Z0, 0Y6V0Z1, 0Y6V0Z2, 0Y6V0Z3, 0Y6W0Z0, 0Y6W0Z1, 0Y6W0Z2, 0Y6W0Z3, 0Y6X0Z0, 0Y6X0Z1, 0Y6X0Z2, 0Y6X0Z3, 0Y6Y0Z0, 0Y6Y0Z1, 0Y6Y0Z2, 0Y6Y0Z3 |

| | | | | |
|---|---|---|---|---|
| **C11** | ONTARGET | 1 | Subarachnoid hemorrhage | Subarachnoid hemorrhage (SAH) ICD-9 diagnosis: 430 . ICD10: I60.00, I60.01, I60.02, I60.10, I60.11, I60.12, I60.2, I60.30, I60.31, I60.32, I60.4, I60.50, I60.51, I60.52, I60.6, I60.7, I60.8, I60.9 |
| **C12** | ONTARGET | 1 | Telmisartan 80 mg | RXNORM: 213432,573321,205305,316765 |
| **C13** | ONTARGET | 1 | Ramipril 10 mg | RXNORM: 35296,316627,261962,401968,574127,260333 |
| **C14** | IMPACT (Pulmonary / COPD) | 1 | COPD | COPD: ICD-9 diagnosis: 491.0, 491.1, 491.20, 491.21, 491.22, 491.8, 491.9, 492.0, 492.8, 496; ICD-10 diagnosis: J41.0, J41.1, J41.8, J42, J43.0, J43.1, J43.2, J43.8, J43.9, J44.0, J44.1, J44.81, J44.89, J44.9. |
| **C15** | IMPACT (Pulmonary / COPD) | 1 | Alpha-1-antitrypsin deficiency | ICD-9: 273.4<br>ICD-10: E88.01 |
| **C16** | IMPACT (Pulmonary / COPD) | 2 | Unstable or life threatening cardiac disease: subjects with any of the following : Myocardial infarction or unstable angina | ACS/unstable angina: ICD-9 diagnosis: 411.0, 411.1, 411.8, 411.81, 411.89; Acute MI: ICD-9 diagnosis: 410.00, 410.01, 410.10, 410.11, 410.20, 410.21, 410.30, 410.31, 410.40, 410.41, 410.50, 410.51, 410.60, 410.61, 410.70, 410.71, 410.80, 410.81, 410.90, 410.91. ICD-10 diagnosis: I20.0, I24.0, I24.8, I24.9, I21.01, I21.02, I21.09, I21.11, I21.19, I21.21, I21.29, I21.3, I21.4, I21.9, I21.A1, I21.A9, I22.0, I22.1, I22.2, I22.8, I22.9 |
| **C17** | IMPACT (Pulmonary / COPD) | 1 | Acute upper or lower respiratory infections | ICD-10 diagnosis: J00, J01.00, J01.01, J01.10, J01.11, J01.20, J01.21, J01.30, J01.31, J01.40, J01.41, J01.80, J01.81, J01.90, J01.91, J02.0, J02.8, J02.9, J03.00, J03.01, J03.80, J03.81, J03.90, J03.91, J04.0, J04.10, J04.11, J04.2, J05.0, J05.10, J05.11, J06.0, J06.9, J20.0, J20.1, J20.2, J20.3, J20.4, J20.5, J20.6, J20.7, J20.8, J20.9, J21.0, J21.1, J21.8, J21.9, J22, J06.8; ICD-9 diagnosis: 460, 461.0, 461.1, 461.2, 461.3, 461.8, 461.9, 462, 463, 464.00, 464.01, 464.10, 464.11, 464.20, 464.21, 464.30, 464.31, 464.4, 464.50, 464.51, 465.0, 465.8, 465.9, 466.0, 466.11, 466.19 |
| **C18** | IMPACT (Pulmonary / COPD) | 2 | Fluticasone furoate / umeclidinium / vilanterol | RXNORM: 1945039,2395769,1945047,2395774,1945041,2395770,1945044,2395771,1945048,2395775,2648696,2656072,2663247 |
| **C19** | vero(Bone / osteoporosis) | 1 | Cirrhosis | ICD-9 Diagnosis: 571.2, 571.5, 571.6<br>ICD-10 Diagnosis: K70.30, K70.31, K74.00, K74.01, K74.02, K74.1, K74.2, K74.3, K74.4, K74.5, K74.60, K74.69 |
| **C20** | vero(Bone / osteoporosis) | 2 | nephrolithiasis or urolithiasis | ICD-9 diagnosis: 592.0, 592.1, 592.9, 274.11<br><br>ICD-10 diagnosis: N20.0, N20.1, N20.2, N20.9, N21.0, N21.1, N21.8, N21.9, N23<br><br>ICD-9 procedure: 57.0, 59.95, 56.0, 98.19, 98.51<br><br>ICD-10-PCS procedure: 0TCB7ZZ, 0TCB8ZZ, 0TFB0ZZ, 0TFB3ZZ, 0TFB4ZZ, 0TFB7ZZ, 0TFB8ZZ, 0TFC0ZZ, 0TFC3ZZ, 0TFC4ZZ, 0TFC7ZZ, 0TFC8ZZ, 0T9B7ZZ, 0T9B8ZZ, 0T9C7ZZ, 0T9C8ZZ, 0TCC7ZZ, 0TCC8ZZ, 0TC37ZZ, 0TC38ZZ, 0TC47ZZ, 0TC48ZZ, 0TC67ZZ, 0TC68ZZ, 0TC77ZZ, 0TC78ZZ, 0T768DZ, 0T778DZ, 0T788DZ, 0TF38ZZ, 0TF48ZZ, 0TF68ZZ, 0TF78ZZ |
| **C21** | vero(Bone / osteoporosis) | 1 | Renal osteodystrophy | ICD-9 diagnosis: 588.0<br>ICD-10 diagnosis: N25.0 |
| **C22** | Lead - 2 (Dibte) | 1 | HbA1c: 7.0-10.0% | Loinc codes: |

| | | | | |
|---|---|---|---|---|
| | | | | 17855-8, 17856-6, 41995-2, 43150-2, 4548-4, 4549-2, 55454-3, 71875-9, 74246-0 |
| **C23** | Lead - 2 (Dibte) | 2 | Clinically significant active cardiovascular disease including history of myocardial infarction and/or heart failure (New York Heart Association class III and IV) | Myocardial Infarction: ICD-9 diagnosis: 410.00, 410.01, 410.02, 410.10, 410.11, 410.12, 410.20, 410.21, 410.22, 410.30, 410.31, 410.32, 410.40, 410.41, 410.42, 410.50, 410.51, 410.52, 410.60, 410.61, 410.62, 410.70, 410.71, 410.72, 410.80, 410.81, 410.82, 410.90, 410.91, 410.92; ICD-10 diagnosis: I21.01, I21.02, I21.09, I21.11, I21.19, I21.21, I21.29, I21.3, I21.4, I21.9, I21.A1, I21.A9, I22.0, I22.1, I22.2, I22.8, I22.9.<br><br>Heart Failure: ICD-9 diagnosis:412, 428.0, 428.1, 428.20, 428.21, 428.22, 428.23, 428.30, 428.31, 428.32, 428.33, 428.40, 428.41, 428.42, 428.43, 428.9, 398.91, 402.01, 402.11, 402.91, 404.01, 404.11, 404.91, 404.03, 404.13, 404.93; ICD-10 diagnosis: I09.81, I11.0, I13.0, I13.2, I50.1, I50.20, I50.21, I50.22, I50.23, I50.30, I50.31, I50.32, I50.33, I50.40, I50.41, I50.42, I50.43, I50.810, I50.811, I50.812, I50.813, I50.814, I50.82, I50.83, I50.84, I50.89, I50.9, I97.130, I97.13, I25.2 |
| **C24** | Lead - 2 (Dibte) | 1 | Morbid obesity diagnosis | Morbid obesity diagnosis: ICD-9 diagnosis: 278.01, 278.03, V85.41, V85.42, V85.43, V85.44, V85.45; ICD-10 diagnosis: E66.01, E66.2, Z68.41, Z68.42, Z68.43, Z68.44, Z68.45. |
| **C25** | Lead - 2 (Dibte) | 2 | Uncontrolled treated/untreated hypertension | Hypertensive crisis: ICD-10 diagnosis: I16.0, I16.1, I16.9.<br><br>Uncontrolled hypertension: ICD-9 diagnosis: 401.9, 401.0, 402.00, 402.01, 403.00, 403.01, 404.00, 404.01, 404.02, 404.03, 405.01, 405.09, 403.90, 403.91, 796.2. |
| **C26** | Lead - 2 (Dibte) | 2 | Liraglutide s.c. 0.6 mg/day, 1.2 mg/day and 1.8 mg/day in combination with metformin | RXNORM: 475968,6809,1163230,1653594,897120,897124,897122,897126 |
| **C27** | Lead - 2 (Dibte) | 1 | Diabetic ketoacidosis (DKA) | ICD-9 diagnosis: 250.10, 250.11, 250.12, 250.13; ICD-10 diagnosis: E08.10, E08.11, E09.10, E09.11, E10.10, E10.11, E11.10, E11.11, E13.10, E13.11. |
| **C28** | AMPLIFY (Cardiovascular / VTE) | 1 | Deep vein thrombosis (DVT) | Deep vein thrombosis (DVT)<br><br>ICD-9-CM diagnosis:<br>451.11, 451.19, 451.81, 451.83,<br>453.40, 453.41, 453.42,<br>453.50, 453.51, 453.52,<br>453.71, 453.72, 453.73, 453.74, 453.75, 453.76, 453.77, 453.79,<br>453.81, 453.82, 453.83, 453.84, 453.85, 453.86, 453.87, 453.89<br><br>ICD-10-CM diagnosis:<br>I80.10, I80.11, I80.12, I80.13,<br>I80.201, I80.202, I80.203, I80.209,<br>I80.211, I80.212, I80.213, I80.219,<br>I80.221, I80.222, I80.223, I80.229,<br>I80.231, I80.232, I80.233, I80.239,<br>I80.291, I80.292, I80.293, I80.299,<br>I82.4Y1, I82.4Y2, I82.4Y3, I82.4Y9,<br>I82.4Z1, I82.4Z2, I82.4Z3, I82.4Z9, |

| | | | | |
|---|---|---|---|---|
| | | | | I82.401, I82.402, I82.403, I82.409, I82.411, I82.412, I82.413, I82.419, I82.421, I82.422, I82.423, I82.429, I82.431, I82.432, I82.433, I82.439, I82.441, I82.442, I82.443, I82.449, I82.451, I82.452, I82.453, I82.459, I82.461, I82.462, I82.463, I82.469, I82.491, I82.492, I82.493, I82.499, I82.5Y1, I82.5Y2, I82.5Y3, I82.5Y9, I82.5Z1, I82.5Z2, I82.5Z3, I82.5Z9, I82.501, I82.502, I82.503, I82.509, I82.511, I82.512, I82.513, I82.519, I82.521, I82.522, I82.523, I82.529, I82.531, I82.532, I82.533, I82.539, I82.541, I82.542, I82.543, I82.549, I82.551, I82.552, I82.553, I82.559, I82.561, I82.562, I82.563, I82.569, I82.591, I82.592, I82.593, I82.599, I82.601, I82.602, I82.603, I82.609, I82.621, I82.622, I82.623, I82.629, I82.701, I82.702, I82.703, I82.709, I82.721, I82.722, I82.723, I82.729 |
| **C29** | AMPLIFY (Cardiovascular / VTE) | 1 | Intracranial bleeding | 430, 431, 432.0, 432.1, 432.9, I60.00, I60.01, I60.02, I60.10, I60.11, I60.12, I60.2, I60.30, I60.31, I60.32, I60.4, I60.50, I60.51, I60.52, I60.6, I60.7, I60.8, I60.9, I61.0, I61.1, I61.2, I61.3, I61.4, I61.5, I61.6, I61.8, I61.9, I62.00, I62.01, I62.02, I62.03, I62.1, I62.9, I69.00, I69.010, I69.011, I69.012, I69.013, I69.014, I69.015, I69.018, I69.019, I69.020, I69.021, I69.022, I69.023, I69.028, I69.031, I69.032, I69.033, I69.034, I69.039, I69.041, I69.042, I69.043, I69.044, I69.049, I69.051, I69.052, I69.053, I69.054, I69.059, I69.061, I69.062, I69.063, I69.064, I69.065, I69.069, I69.090, I69.091, I69.092, I69.093, I69.098, I69.10, I69.110, I69.111, I69.112, I69.113, I69.114, I69.115, I69.118, I69.119, I69.120, I69.121, I69.122, I69.123, I69.128, I69.131, I69.132, I69.133, I69.134, I69.139, I69.141, I69.142, I69.143, I69.144, I69.149, I69.151, I69.152, I69.153, I69.154, I69.159, I69.161, I69.162, I69.163, I69.164, I69.165, I69.169, I69.190, I69.191, I69.192, I69.193, I69.198, I69.20, I69.210, I69.211, I69.212, I69.213, I69.214, I69.215, I69.218, I69.219, I69.220, I69.221, I69.222, I69.223, I69.228, I69.231, I69.232, I69.233, I69.234, I69.239, I69.241, I69.242, I69.243, I69.244, I69.249, I69.251, I69.252, I69.253, I69.254, I69.259, I69.261, I69.262, I69.263, I69.264, I69.265, I69.269, I69.290, I69.291, I69.292, I69.293, I69.298 |
| **C30** | AMPLIFY (Cardiovascular / VTE) | 3 | Gastrointestinal bleeding and/or endoscopically verified ulcer disease | ICD-9-CM diagnosis:<br>531.00, 531.01, 531.10, 531.11, 531.20, 531.21, 531.30, 531.31, 531.40, 531.41, 531.50, 531.51, 531.60, 531.61, 531.70, 531.71, 531.90, 531.91,<br>532.00, 532.01, 532.10, 532.11, 532.20, 532.21, 532.30, 532.31, 532.40, 532.41, 532.50, 532.51, 532.60, 532.61, 532.70, 532.71, 532.90, 532.91,<br>533.00, 533.01, 533.10, 533.11, 533.20, 533.21, 533.30, 533.31, 533.40, 533.41, 533.50, 533.51, 533.60, 533.61, 533.70, 533.71, |

| | | | | |
|---|---|---|---|---|
| | | | | 533.90, 533.91,<br>534.00, 534.01, 534.10, 534.11, 534.20, 534.21, 534.30, 534.31,<br>534.40, 534.41, 534.50, 534.51, 534.60, 534.61, 534.70, 534.71,<br>534.90, 534.91,<br>562.01, 562.03, 562.12, 562.13,<br>569.3, 569.84, 569.85, 569.86,<br>578.0, 578.1, 578.9<br><br>ICD-10-CM diagnosis:<br>K25.0, K25.1, K25.2, K25.3, K25.4, K25.5, K25.6, K25.7, K25.9,<br>K26.0, K26.1, K26.2, K26.3, K26.4, K26.5, K26.6, K26.7, K26.9,<br>K27.0, K27.1, K27.2, K27.3, K27.4, K27.5, K27.6, K27.7, K27.9,<br>K28.0, K28.1, K28.2, K28.3, K28.4, K28.5, K28.6, K28.7, K28.9,<br>K55.21,<br>K57.01, K57.11, K57.13, K57.21, K57.31, K57.33, K57.41, K57.51,<br>K57.53, K57.81, K57.91, K57.93,<br>K62.5, K63.81,<br>K92.0, K92.1, K92.2<br><br>ICD-9 procedure:<br>44.43<br><br>ICD-10-PCS procedure:<br>0W3P8ZZ |
| **C31** | AMPLIFY (Cardiovascular / VTE) | 2 | Mechanical valve | Heart valve replacement/repair: ICD-9 diagnosis: V43.3; ICD-10 diagnosis: Z95.2, Z95.3, Z95.4; ICD-9 procedure: 35.00, 35.01, 35.02, 35.03, 35.04, 35.20, 35.21, 35.22, 35.23, 35.24, 35.25, 35.26, 35.27, 35.28, 35.31, 35.32, 35.33, 35.34, 35.35, 35.39; ICD-10-PCS procedure prefix: 02RG, 02RF, 02RH, 02RJ, 02QG, 02QF, 02QH, 02QJ. |
| **C32** | AMPLIFY (Cardiovascular / VTE) | 1 | Apixaban 5 mg twice daily | RXNORM: 1364430,1364444,1364445,1364446,1364447 |
| **C33** | ARISTOTLE | 2 | alcohol or drug abuse | Alcohol or drug abuse<br><br>Alcohol abuse or dependence<br>ICD-9-CM diagnosis:<br>291.0, 291.1, 291.2, 291.3, 291.4, 291.5, 291.81, 291.82, 291.89,<br>291.9,<br>303.00, 303.01, 303.02, 303.03, 303.90, 303.91, 303.92, 303.93,<br>305.00, 305.01, 305.02, 305.03,<br>357.5, 425.5,<br>571.0, 571.1, 571.2, 571.3,<br>V11.3<br><br>ICD-10-CM diagnosis:<br>F10.10, F10.11, F10.120, F10.121, F10.129, F10.130, F10.131,<br>F10.132, F10.139, F10.14, F10.150, F10.151, F10.159, F10.180,<br>F10.181, F10.182, F10.188, F10.19,<br>F10.20, F10.21, F10.220, F10.221, F10.229, F10.230, F10.231, |

F10.232, F10.239, F10.24, F10.250, F10.251, F10.259, F10.26,
F10.27, F10.280, F10.281, F10.282, F10.288, F10.29,
G62.1, I42.6,
K70.0, K70.10, K70.11, K70.2, K70.30, K70.31, K70.40, K70.41, K70.9,
Z87.898

Drug abuse or dependence
ICD-9-CM diagnosis:
292.0, 292.11, 292.12, 292.2, 292.81, 292.82, 292.83, 292.84, 292.85,
292.89, 292.9,
304.00, 304.01, 304.02, 304.03,
304.10, 304.11, 304.12, 304.13,
304.20, 304.21, 304.22, 304.23,
304.30, 304.31, 304.32, 304.33,
304.40, 304.41, 304.42, 304.43,
304.50, 304.51, 304.52, 304.53,
304.60, 304.61, 304.62, 304.63,
304.70, 304.71, 304.72, 304.73,
304.80, 304.81, 304.82, 304.83,
304.90, 304.91, 304.92, 304.93,
305.20, 305.21, 305.22, 305.23,
305.30, 305.31, 305.32, 305.33,
305.40, 305.41, 305.42, 305.43,
305.50, 305.51, 305.52, 305.53,
305.60, 305.61, 305.62, 305.63,
305.70, 305.71, 305.72, 305.73,
305.80, 305.81, 305.82, 305.83,
305.90, 305.91, 305.92, 305.93,
648.30, 648.31, 648.32, 648.33, 648.34

ICD-10-CM diagnosis:
F11.10, F11.11, F11.120, F11.121, F11.122, F11.129, F11.14,
F11.150, F11.151, F11.159, F11.180, F11.181, F11.182, F11.188,
F11.19,
F11.20, F11.21, F11.220, F11.221, F11.222, F11.229, F11.23, F11.24,
F11.250, F11.251, F11.259, F11.280, F11.281, F11.282, F11.288,
F11.29,
F12.10, F12.11, F12.120, F12.121, F12.122, F12.129, F12.150,
F12.151, F12.159, F12.180, F12.188, F12.19,
F12.20, F12.21, F12.220, F12.221, F12.222, F12.229, F12.250,
F12.251, F12.259, F12.280, F12.288, F12.29,
F13.10, F13.11, F13.120, F13.121, F13.129, F13.130, F13.131,
F13.132, F13.139, F13.14, F13.150, F13.151, F13.159, F13.180,
F13.181, F13.182, F13.188, F13.19,
F13.20, F13.21, F13.220, F13.221, F13.229, F13.230, F13.231,
F13.232, F13.239, F13.24, F13.250, F13.251, F13.259, F13.26,
F13.27, F13.280, F13.281, F13.282, F13.288, F13.29,
F14.10, F14.11, F14.120, F14.121, F14.122, F14.129, F14.14,
F14.150, F14.151, F14.159, F14.180, F14.181, F14.182, F14.188,
F14.19,

| | | | | |
|---|---|---|---|---|
| | | | | F14.20, F14.21, F14.220, F14.221, F14.222, F14.229, F14.23, F14.24, F14.250, F14.251, F14.259, F14.280, F14.281, F14.282, F14.288, F14.29,<br>F15.10, F15.11, F15.120, F15.121, F15.122, F15.129, F15.14, F15.150, F15.151, F15.159, F15.180, F15.181, F15.182, F15.188, F15.19,<br>F15.20, F15.21, F15.220, F15.221, F15.222, F15.229, F15.23, F15.24, F15.250, F15.251, F15.259, F15.280, F15.281, F15.282, F15.288, F15.29,<br>F16.10, F16.11, F16.120, F16.121, F16.122, F16.129, F16.14, F16.150, F16.151, F16.159, F16.180, F16.183, F16.188, F16.19,<br>F16.20, F16.21, F16.220, F16.221, F16.229, F16.24, F16.250, F16.251, F16.259, F16.280, F16.283, F16.288, F16.29,<br>F18.10, F18.11, F18.120, F18.121, F18.129, F18.14, F18.150, F18.151, F18.159, F18.17, F18.180, F18.188, F18.19,<br>F18.20, F18.21, F18.220, F18.221, F18.229, F18.24, F18.250, F18.251, F18.259, F18.27, F18.280, F18.288, F18.29,<br>F19.10, F19.11, F19.120, F19.121, F19.122, F19.129, F19.130, F19.131, F19.132, F19.139, F19.14, F19.150, F19.151, F19.159, F19.16, F19.17, F19.180, F19.181, F19.182, F19.188, F19.19,<br>F19.20, F19.21, F19.220, F19.221, F19.222, F19.229, F19.230, F19.231, F19.232, F19.239, F19.24, F19.250, F19.251, F19.259, F19.26, F19.27, F19.280, F19.281, F19.282, F19.288, F19.29 |
| **C34** | INSPIRE | 2 | atopic dermatitis and/or allergic rhinitis | Atopic dermatitis: ICD-9 diagnosis: 691.0, 691.8, 691.9; ICD-10 diagnosis: L20.0, L20.81, L20.82, L20.83, L20.84, L20.89, L20.9.<br><br>Allergic rhinitis: ICD-9 diagnosis: 477.0, 477.1, 477.2, 477.8, 477.9; ICD-10 diagnosis: J30.0, J30.1, J30.2, J30.5, J30.81, J30.89, J30.9. |
| **C35** | leader | 1 | Lupus OR Crohn’s disease and other IBD | Lupus: ICD-9 diagnosis: 710.0; ICD-10 diagnosis: M32.10.<br><br>Crohn’s disease and other IBD: ICD-9 diagnosis: 555.0, 555.1, 555.2, 555.9, 556.0, 556.1, 556.2, 556.3, 556.4, 556.5, 556.6, 556.8, 556.9; ICD-10 diagnosis: K50.00, K50.011, K50.012, K50.013, K50.014, K50.018, K50.019, K50.10, K50.111, K50.112, K50.113, K50.114, K50.118, K50.119, K50.80, K50.811, K50.812, K50.813, K50.814, K50.818, K50.819, K50.90, K50.911, K50.912, K50.913, K50.914, K50.918, K50.919, K51.00, K51.011, K51.012, K51.013, K51.014, K51.018, K51.019, K51.20, K51.211, K51.212, K51.213, K51.214, K51.218, K51.219, K51.30, K51.311, K51.312, K51.313, K51.314, K51.318, K51.319, K51.40, K51.411, K51.412, K51.413, K51.414, K51.418, K51.419, K51.50, K51.511, K51.512, K51.513, K51.514, K51.518, K51.519, K51.80, K51.811, K51.812, K51.813, K51.814, K51.818, K51.819, K51.90, K51.911, K51.912, K51.913, K51.914, K51.918, K51.919. |
| **C36** | leader | 2 | Hyperparathyroidism and hyperthyroidism | Parathyroid disorder: ICD-9 diagnosis: 252.0, 252.00, 252.01, 252.02, 252.08, 252.1, 252.8, 252.9; ICD-10 diagnosis: E20.0, E20.1, E20.8, E20.9, E21.0, E21.1, E21.2, E21.3, E21.4, E21.5. |
| **C37** | leader | 2 | Cushing syndrome and disorders of adrenal glands | ICD-9 diagnosis: 255.0, 255.10, 255.11, 255.12, 255.13, 255.14, 255.2, 255.3, 255.41, 255.42, 255.5, 255.6, 255.8, 255.9; ICD-10 diagnosis: E24.0, E24.1, E24.2, E24.3, E24.4, E24.8, E24.9, E25.0, E25.8, E25.9, |

| | | | | |
|---|---|---|---|---|
| | | | | E26.01, E26.02, E26.09, E26.1, E26.81, E26.89, E26.9, E27.0, E27.1, E27.2, E27.3, E27.40, E27.49, E27.5, E27.8, E27.9. |
| **C38** | RECORD1 | 1 | Congestive heart failure | 428.0, 428.1, 428.20, 428.21, 428.22, 428.23, 428.30, 428.31, 428.32, 428.33, 428.40, 428.41, 428.42, 428.43, 428.9, 398.91, 402.01, 402.11, 402.91, 404.01, 404.11, 404.91, 404.03, 404.13, 404.93 I50.1, I50.20, I50.21, I50.22, I50.23, I50.30, I50.31, I50.32, I50.33, I50.40, I50.41, I50.42, I50.43, I50.810, I50.811, I50.812, I50.813, I50.814, I50.82, I50.83, I50.84, I50.89, I50.9, I09.81, I11.0, I13.0, I13.2 |
| **C39** | PRONOUNCE_ | 3 | nonfatal myocardial infarction, nonfatal stroke | Nonfatal MI ICD-9 : 410.00, 410.01, 410.10, 410.11, 410.20, 410.21, 410.30, 410.31, 410.40, 410.41, 410.50, 410.51, 410.60, 410.61, 410.70, 410.71, 410.80, 410.81, 410.90, 410.91<br>Nonfatal MI — ICD-10 : I21.01, I21.02, I21.09, I21.11, I21.19, I21.21, I21.29, I21.3, I21.4, I21.A1, I21.A9, I21.B, I22.0, I22.1, I22.2, I22.8, I22.9<br>Nonfatal Stroke — ICD-9 : 430, 431, 432.0, 432.1, 432.9, 433.01, 433.11, 433.21, 433.31, 433.81, 433.91, 434.01, 434.11, 434.91, 436<br>Nonfatal Stroke — ICD-10 : I60.00, I60.01, I60.02, I60.10, I60.11, I60.12, I60.2, I60.30, I60.31, I60.32, I60.4, I60.50, I60.51, I60.52, I60.6, I60.7, I60.8, I60.9, I61.0, I61.1, I61.2, I61.3, I61.4, I61.5, I61.6, I61.8, I61.9, I63.00, I63.011, I63.012, I63.019, I63.02, I63.031, I63.032, I63.039, I63.09, I63.10, I63.111, I63.112, I63.119, I63.12, I63.131, I63.132, I63.139, I63.19, I63.20, I63.211, I63.212, I63.219, I63.22, I63.231, I63.232, I63.239, I63.29, I63.30, I63.311, I63.312, I63.319, I63.321, I63.322, I63.329, I63.331, I63.332, I63.339, I63.341, I63.342, I63.349, I63.39, I63.40, I63.411, I63.412, I63.419, I63.421, I63.422, I63.429, I63.431, I63.432, I63.439, I63.441, I63.442, I63.449, I63.49, I63.50, I63.511, I63.512, I63.519, I63.521, I63.522, I63.529, I63.531, I63.532, I63.539, I63.541, I63.542, I63.549, I63.59, I63.6, I63.8, I63.9 |
| **C40** | PRONOUNCE_ | 2 | Hypokalemia and Hypomagnesemia | ICD-9 diagnosis: 276.8<br>, 275.2<br>ICD-10 diagnosis: E83.42, E87.6 |

#### Supplementary Table S2. Per-criterion performance by pipeline and model

Precision, recall, and F1 were calculated from exact vocabulary-matched code pairs. TP denotes true positives, FP false positives, and FN false negatives. The full accepted code strings are provided in the accompanying Excel workbook.

| ID | Pipeline | Model | Trial | Tier | TP | FP | FN | Precision | Recall | F1 |
|---|---|---|---|---|---|---|---|---|---|---|
| **C01** | Baseline | Claude | DAPA-CKD(Kidney / cardio-renal) | 1 | 2 | 0 | 0 | 1 | 1 | 1 |
| **C02** | Baseline | Claude | DAPA-CKD(Kidney / cardio-renal) | 2 | 7 | 5 | 1 | 0.583 | 0.875 | 0.700 |
| **C03** | Baseline | Claude | DAPA-CKD(Kidney / cardio-renal) | 1 | 107 | 0 | 2 | 1 | 0.982 | 0.991 |
| **C04** | Baseline | Claude | DAPA-CKD(Kidney / cardio-renal) | 2 | 5 | 0 | 2 | 1 | 0.714 | 0.833 |
| **C05** | Baseline | Claude | DAPA-CKD(Kidney / cardio-renal) | 1 | 10 | 0 | 0 | 1 | 1 | 1 |
| **C06** | Baseline | Claude | DAPA-CKD(Kidney / cardio-renal) | 1 | 1 | 0 | 4 | 1 | 0.200 | 0.333 |
| **C07** | Baseline | Claude | ONTARGET | 1 | 4 | 0 | 0 | 1 | 1 | 1 |
| **C08** | Baseline | Claude | ONTARGET | 3 | 17 | 204 | 181 | 0.077 | 0.086 | 0.081 |
| **C09** | Baseline | Claude | ONTARGET | 3 | 83 | 3 | 52 | 0.965 | 0.615 | 0.751 |
| **C10** | Baseline | Claude | ONTARGET | 1 | 44 | 1 | 83 | 0.978 | 0.346 | 0.512 |
| **C11** | Baseline | Claude | ONTARGET | 1 | 18 | 0 | 1 | 1 | 0.947 | 0.973 |
| **C12** | Baseline | Claude | ONTARGET | 1 | 0 | 1 | 4 | 0 | 0 | 0 |
| **C13** | Baseline | Claude | ONTARGET | 1 | 1 | 0 | 5 | 1 | 0.167 | 0.286 |
| **C14** | Baseline | Claude | IMPACT (Pulmonary / COPD) | 1 | 19 | 0 | 5 | 1 | 0.792 | 0.884 |
| **C15** | Baseline | Claude | IMPACT (Pulmonary / COPD) | 1 | 1 | 1 | 1 | 0.500 | 0.500 | 0.500 |
| **C16** | Baseline | Claude | IMPACT (Pulmonary / COPD) | 2 | 29 | 8 | 17 | 0.784 | 0.630 | 0.699 |
| **C17** | Baseline | Claude | IMPACT (Pulmonary / COPD) | 1 | 69 | 2 | 6 | 0.972 | 0.920 | 0.945 |
| **C18** | Baseline | Claude | IMPACT (Pulmonary / COPD) | 2 | 0 | 0 | 13 | 0 | 0 | 0 |
| **C19** | Baseline | Claude | vero(Bone / osteoporosis) | 1 | 12 | 1 | 3 | 0.923 | 0.800 | 0.857 |
| **C20** | Baseline | Claude | vero(Bone / | 2 | 12 | 7 | 39 | 0.632 | 0.235 | 0.343 |

| | | | | | | | | | | |
|---|---|---|---|---|---|---|---|---|---|---|
| | | | osteoporosis) | | | | | | | |
| **C21** | Baseline | Claude | vero(Bone / osteoporosis) | 1 | 2 | 0 | 0 | 1 | 1 | 1 |
| **C22** | Baseline | Claude | Lead - 2 (Dibte) | 1 | 4 | 2 | 5 | 0.667 | 0.444 | 0.533 |
| **C23** | Baseline | Claude | Lead - 2 (Dibte) | 2 | 72 | 0 | 31 | 1 | 0.699 | 0.823 |
| **C24** | Baseline | Claude | Lead - 2 (Dibte) | 1 | 13 | 0 | 1 | 1 | 0.929 | 0.963 |
| **C25** | Baseline | Claude | Lead - 2 (Dibte) | 2 | 3 | 3 | 15 | 0.500 | 0.167 | 0.250 |
| **C26** | Baseline | Claude | Lead - 2 (Dibte) | 2 | 2 | 2 | 6 | 0.500 | 0.250 | 0.333 |
| **C27** | Baseline | Claude | Lead - 2 (Dibte) | 1 | 14 | 0 | 0 | 1 | 1 | 1 |
| **C28** | Baseline | Claude | AMPLIFY (Cardiovascular / VTE) | 1 | 94 | 9 | 52 | 0.913 | 0.644 | 0.755 |
| **C29** | Baseline | Claude | AMPLIFY (Cardiovascular / VTE) | 1 | 38 | 119 | 120 | 0.242 | 0.241 | 0.241 |
| **C30** | Baseline | Claude | AMPLIFY (Cardiovascular / VTE) | 3 | 116 | 26 | 23 | 0.817 | 0.835 | 0.826 |
| **C31** | Baseline | Claude | AMPLIFY (Cardiovascular / VTE) | 2 | 8 | 4 | 16 | 0.667 | 0.333 | 0.444 |
| **C32** | Baseline | Claude | AMPLIFY (Cardiovascular / VTE) | 1 | 1 | 0 | 4 | 1 | 0.200 | 0.333 |
| **C33** | Baseline | Claude | ARISTOTLE | 2 | 370 | 5 | 47 | 0.987 | 0.887 | 0.934 |
| **C34** | Baseline | Claude | INSPIRE | 2 | 19 | 0 | 3 | 1 | 0.864 | 0.927 |
| **C35** | Baseline | Claude | leader | 1 | 0 | 0 | 92 | 0 | 0 | 0 |
| **C36** | Baseline | Claude | leader | 2 | 8 | 28 | 10 | 0.222 | 0.444 | 0.296 |
| **C37** | Baseline | Claude | leader | 2 | 40 | 0 | 0 | 1 | 1 | 1 |
| **C38** | Baseline | Claude | RECORD1 | 1 | 29 | 0 | 23 | 1 | 0.558 | 0.716 |
| **C39** | Baseline | Claude | PRONOUNCE_ | 3 | 143 | 29 | 8 | 0.831 | 0.947 | 0.885 |
| **C40** | Baseline | Claude | PRONOUNCE_ | 2 | 4 | 0 | 0 | 1 | 1 | 1 |
| **C01** | Baseline | GPT-5.5 | DAPA-CKD(Kidney / cardio-renal) | 1 | 2 | 0 | 0 | 1 | 1 | 1 |
| **C02** | Baseline | GPT-5.5 | DAPA-CKD(Kidney / cardio-renal) | 2 | 8 | 5 | 0 | 0.615 | 1 | 0.762 |
| **C03** | Baseline | GPT-5.5 | DAPA-CKD(Kidney / cardio-renal) | 1 | 84 | 6 | 25 | 0.933 | 0.771 | 0.844 |
| **C04** | Baseline | GPT-5.5 | DAPA- | 2 | 4 | 0 | 3 | 1 | 0.571 | 0.727 |

| | | | | | | | | | | |
|---|---|---|---|---|---|---|---|---|---|---|
| | | | CKD(Kidney / cardio-renal) | | | | | | | |
| **C05** | Baseline | GPT-5.5 | DAPA-CKD(Kidney / cardio-renal) | 1 | 10 | 0 | 0 | 1 | 1 | 1 |
| **C06** | Baseline | GPT-5.5 | DAPA-CKD(Kidney / cardio-renal) | 1 | 1 | 1 | 4 | 0.500 | 0.200 | 0.286 |
| **C07** | Baseline | GPT-5.5 | ONTARGET | 1 | 4 | 0 | 0 | 1 | 1 | 1 |
| **C08** | Baseline | GPT-5.5 | ONTARGET | 3 | 27 | 198 | 171 | 0.120 | 0.136 | 0.128 |
| **C09** | Baseline | GPT-5.5 | ONTARGET | 3 | 132 | 206 | 3 | 0.391 | 0.978 | 0.558 |
| **C10** | Baseline | GPT-5.5 | ONTARGET | 1 | 21 | 0 | 106 | 1 | 0.165 | 0.284 |
| **C11** | Baseline | GPT-5.5 | ONTARGET | 1 | 19 | 0 | 0 | 1 | 1 | 1 |
| **C12** | Baseline | GPT-5.5 | ONTARGET | 1 | 1 | 0 | 3 | 1 | 0.250 | 0.400 |
| **C13** | Baseline | GPT-5.5 | ONTARGET | 1 | 1 | 0 | 5 | 1 | 0.167 | 0.286 |
| **C14** | Baseline | GPT-5.5 | IMPACT (Pulmonary / COPD) | 1 | 23 | 3 | 1 | 0.885 | 0.958 | 0.920 |
| **C15** | Baseline | GPT-5.5 | IMPACT (Pulmonary / COPD) | 1 | 2 | 0 | 0 | 1 | 1 | 1 |
| **C16** | Baseline | GPT-5.5 | IMPACT (Pulmonary / COPD) | 2 | 39 | 19 | 7 | 0.672 | 0.848 | 0.750 |
| **C17** | Baseline | GPT-5.5 | IMPACT (Pulmonary / COPD) | 1 | 74 | 81 | 1 | 0.477 | 0.987 | 0.643 |
| **C18** | Baseline | GPT-5.5 | IMPACT (Pulmonary / COPD) | 2 | 2 | 0 | 11 | 1 | 0.154 | 0.267 |
| **C19** | Baseline | GPT-5.5 | vero(Bone / osteoporosis) | 1 | 10 | 2 | 5 | 0.833 | 0.667 | 0.741 |
| **C20** | Baseline | GPT-5.5 | vero(Bone / osteoporosis) | 2 | 19 | 17 | 32 | 0.528 | 0.373 | 0.437 |
| **C21** | Baseline | GPT-5.5 | vero(Bone / osteoporosis) | 1 | 2 | 0 | 0 | 1 | 1 | 1 |
| **C22** | Baseline | GPT-5.5 | Lead - 2 (Dibte) | 1 | 5 | 2 | 4 | 0.714 | 0.556 | 0.625 |
| **C23** | Baseline | GPT-5.5 | Lead - 2 (Dibte) | 2 | 20 | 1 | 83 | 0.952 | 0.194 | 0.323 |
| **C24** | Baseline | GPT-5.5 | Lead - 2 (Dibte) | 1 | 14 | 1 | 0 | 0.933 | 1 | 0.966 |
| **C25** | Baseline | GPT-5.5 | Lead - 2 (Dibte) | 2 | 14 | 6 | 4 | 0.700 | 0.778 | 0.737 |
| **C26** | Baseline | GPT-5.5 | Lead - 2 (Dibte) | 2 | 2 | 0 | 6 | 1 | 0.250 | 0.400 |
| **C27** | Baseline | GPT-5.5 | Lead - 2 (Dibte) | 1 | 14 | 2 | 0 | 0.875 | 1 | 0.933 |
| **C28** | Baseline | GPT-5.5 | AMPLIFY (Cardiovascula | 1 | 111 | 40 | 35 | 0.735 | 0.760 | 0.747 |

| | | | | | | | | | | |
|---|---|---|---|---|---|---|---|---|---|---|
| | | | r / VTE) | | | | | | | |
| **C29** | Baseline | GPT-5.5 | AMPLIFY (Cardiovascular / VTE) | 1 | 38 | 382 | 120 | 0.090 | 0.241 | 0.131 |
| **C30** | Baseline | GPT-5.5 | AMPLIFY (Cardiovascular / VTE) | 3 | 72 | 27 | 67 | 0.727 | 0.518 | 0.605 |
| **C31** | Baseline | GPT-5.5 | AMPLIFY (Cardiovascular / VTE) | 2 | 7 | 12 | 17 | 0.368 | 0.292 | 0.326 |
| **C32** | Baseline | GPT-5.5 | AMPLIFY (Cardiovascular / VTE) | 1 | 1 | 0 | 4 | 1 | 0.200 | 0.333 |
| **C33** | Baseline | GPT-5.5 | ARISTOTLE | 2 | 390 | 20 | 27 | 0.951 | 0.935 | 0.943 |
| **C34** | Baseline | GPT-5.5 | INSPIRE | 2 | 19 | 0 | 3 | 1 | 0.864 | 0.927 |
| **C35** | Baseline | GPT-5.5 | leader | 1 | 85 | 22 | 7 | 0.794 | 0.924 | 0.854 |
| **C36** | Baseline | GPT-5.5 | leader | 2 | 8 | 32 | 10 | 0.200 | 0.444 | 0.276 |
| **C37** | Baseline | GPT-5.5 | leader | 2 | 40 | 1 | 0 | 0.976 | 1 | 0.988 |
| **C38** | Baseline | GPT-5.5 | RECORD1 | 1 | 52 | 2 | 0 | 0.963 | 1 | 0.981 |
| **C39** | Baseline | GPT-5.5 | PRONOUNCE_ | 3 | 129 | 57 | 22 | 0.694 | 0.854 | 0.766 |
| **C40** | Baseline | GPT-5.5 | PRONOUNCE_ | 2 | 4 | 0 | 0 | 1 | 1 | 1 |
| **C01** | Baseline | Qwen | DAPA-CKD(Kidney / cardio-renal) | 1 | 2 | 0 | 0 | 1 | 1 | 1 |
| **C02** | Baseline | Qwen | DAPA-CKD(Kidney / cardio-renal) | 2 | 8 | 0 | 0 | 1 | 1 | 1 |
| **C03** | Baseline | Qwen | DAPA-CKD(Kidney / cardio-renal) | 1 | 3 | 0 | 106 | 1 | 0.028 | 0.054 |
| **C04** | Baseline | Qwen | DAPA-CKD(Kidney / cardio-renal) | 2 | 3 | 0 | 4 | 1 | 0.429 | 0.600 |
| **C05** | Baseline | Qwen | DAPA-CKD(Kidney / cardio-renal) | 1 | 10 | 0 | 0 | 1 | 1 | 1 |
| **C06** | Baseline | Qwen | DAPA-CKD(Kidney / cardio-renal) | 1 | 0 | 0 | 5 | 0 | 0 | 0 |
| **C07** | Baseline | Qwen | ONTARGET | 1 | 4 | 0 | 0 | 1 | 1 | 1 |
| **C08** | Baseline | Qwen | ONTARGET | 3 | 5 | 4 | 193 | 0.556 | 0.025 | 0.048 |
| **C09** | Baseline | Qwen | ONTARGET | 3 | 50 | 10 | 85 | 0.833 | 0.370 | 0.513 |
| **C10** | Baseline | Qwen | ONTARGET | 1 | 17 | 4 | 110 | 0.810 | 0.134 | 0.230 |
| **C11** | Baseline | Qwen | ONTARGET | 1 | 17 | 26 | 2 | 0.395 | 0.895 | 0.548 |
| **C12** | Baseline | Qwen | ONTARGET | 1 | 0 | 0 | 4 | 0 | 0 | 0 |
| **C13** | Baseline | Qwen | ONTARGET | 1 | 0 | 0 | 6 | 0 | 0 | 0 |
| **C14** | Baseline | Qwen | IMPACT (Pulmonary / | 1 | 5 | 0 | 19 | 1 | 0.208 | 0.345 |

| | | | | | | | | | | |
|---|---|---|---|---|---|---|---|---|---|---|
| | | | COPD) | | | | | | | |
| **C15** | Baseline | Qwen | IMPACT (Pulmonary / COPD) | 1 | 1 | 0 | 1 | 1 | 0.500 | 0.667 |
| **C16** | Baseline | Qwen | IMPACT (Pulmonary / COPD) | 2 | 28 | 10 | 18 | 0.737 | 0.609 | 0.667 |
| **C17** | Baseline | Qwen | IMPACT (Pulmonary / COPD) | 1 | 67 | 69 | 8 | 0.493 | 0.893 | 0.635 |
| **C18** | Baseline | Qwen | IMPACT (Pulmonary / COPD) | 2 | 0 | 0 | 13 | 0 | 0 | 0 |
| **C19** | Baseline | Qwen | vero(Bone / osteoporosis) | 1 | 12 | 0 | 3 | 1 | 0.800 | 0.889 |
| **C20** | Baseline | Qwen | vero(Bone / osteoporosis) | 2 | 11 | 13 | 40 | 0.458 | 0.216 | 0.293 |
| **C21** | Baseline | Qwen | vero(Bone / osteoporosis) | 1 | 2 | 0 | 0 | 1 | 1 | 1 |
| **C22** | Baseline | Qwen | Lead - 2 (Dibte) | 1 | 2 | 2 | 7 | 0.500 | 0.222 | 0.308 |
| **C23** | Baseline | Qwen | Lead - 2 (Dibte) | 2 | 22 | 7 | 81 | 0.759 | 0.214 | 0.333 |
| **C24** | Baseline | Qwen | Lead - 2 (Dibte) | 1 | 3 | 0 | 11 | 1 | 0.214 | 0.353 |
| **C25** | Baseline | Qwen | Lead - 2 (Dibte) | 2 | 5 | 9 | 13 | 0.357 | 0.278 | 0.313 |
| **C26** | Baseline | Qwen | Lead - 2 (Dibte) | 2 | 1 | 0 | 7 | 1 | 0.125 | 0.222 |
| **C27** | Baseline | Qwen | Lead - 2 (Dibte) | 1 | 10 | 0 | 4 | 1 | 0.714 | 0.833 |
| **C28** | Baseline | Qwen | AMPLIFY (Cardiovascula r / VTE) | 1 | 4 | 0 | 142 | 1 | 0.027 | 0.053 |
| **C29** | Baseline | Qwen | AMPLIFY (Cardiovascula r / VTE) | 1 | 35 | 95 | 123 | 0.269 | 0.222 | 0.243 |
| **C30** | Baseline | Qwen | AMPLIFY (Cardiovascula r / VTE) | 3 | 108 | 6 | 31 | 0.947 | 0.777 | 0.854 |
| **C31** | Baseline | Qwen | AMPLIFY (Cardiovascula r / VTE) | 2 | 0 | 1 | 24 | 0 | 0 | 0 |
| **C32** | Baseline | Qwen | AMPLIFY (Cardiovascula r / VTE) | 1 | 1 | 0 | 4 | 1 | 0.200 | 0.333 |
| **C33** | Baseline | Qwen | ARISTOTLE | 2 | 45 | 4 | 372 | 0.918 | 0.108 | 0.193 |
| **C34** | Baseline | Qwen | INSPIRE | 2 | 19 | 0 | 3 | 1 | 0.864 | 0.927 |
| **C35** | Baseline | Qwen | leader | 1 | 91 | 13 | 1 | 0.875 | 0.989 | 0.929 |
| **C36** | Baseline | Qwen | leader | 2 | 8 | 24 | 10 | 0.250 | 0.444 | 0.320 |

| | | | | | | | | | | |
|---|---|---|---|---|---|---|---|---|---|---|
| **C37** | Baseline | Qwen | leader | 2 | 28 | 0 | 12 | 1 | 0.700 | 0.824 |
| **C38** | Baseline | Qwen | RECORD1 | 1 | 25 | 0 | 27 | 1 | 0.481 | 0.649 |
| **C39** | Baseline | Qwen | PRONOUNCE_ | 3 | 62 | 232 | 89 | 0.211 | 0.411 | 0.279 |
| **C40** | Baseline | Qwen | PRONOUNCE_ | 2 | 3 | 0 | 1 | 1 | 0.750 | 0.857 |
| **C01** | Baseline | MedGemma | DAPA-CKD(Kidney / cardio-renal) | 1 | 2 | 0 | 0 | 1 | 1 | 1 |
| **C02** | Baseline | MedGemma | DAPA-CKD(Kidney / cardio-renal) | 2 | 4 | 1 | 4 | 0.800 | 0.500 | 0.615 |
| **C03** | Baseline | MedGemma | DAPA-CKD(Kidney / cardio-renal) | 1 | 2 | 0 | 107 | 1 | 0.018 | 0.036 |
| **C04** | Baseline | MedGemma | DAPA-CKD(Kidney / cardio-renal) | 2 | 0 | 1 | 7 | 0 | 0 | 0 |
| **C05** | Baseline | MedGemma | DAPA-CKD(Kidney / cardio-renal) | 1 | 5 | 0 | 5 | 1 | 0.500 | 0.667 |
| **C06** | Baseline | MedGemma | DAPA-CKD(Kidney / cardio-renal) | 1 | 0 | 0 | 5 | 0 | 0 | 0 |
| **C07** | Baseline | MedGemma | ONTARGET | 1 | 2 | 0 | 2 | 1 | 0.500 | 0.667 |
| **C08** | Baseline | MedGemma | ONTARGET | 3 | 0 | 14 | 198 | 0 | 0 | 0 |
| **C09** | Baseline | MedGemma | ONTARGET | 3 | 122 | 101 | 13 | 0.547 | 0.904 | 0.682 |
| **C10** | Baseline | MedGemma | ONTARGET | 1 | 4 | 1 | 123 | 0.800 | 0.031 | 0.061 |
| **C11** | Baseline | MedGemma | ONTARGET | 1 | 16 | 0 | 3 | 1 | 0.842 | 0.914 |
| **C12** | Baseline | MedGemma | ONTARGET | 1 | 0 | 0 | 4 | 0 | 0 | 0 |
| **C13** | Baseline | MedGemma | ONTARGET | 1 | 0 | 20 | 6 | 0 | 0 | 0 |
| **C14** | Baseline | MedGemma | IMPACT (Pulmonary / COPD) | 1 | 7 | 0 | 17 | 1 | 0.292 | 0.452 |
| **C15** | Baseline | MedGemma | IMPACT (Pulmonary / COPD) | 1 | 0 | 0 | 2 | 0 | 0 | 0 |
| **C16** | Baseline | MedGemma | IMPACT (Pulmonary / COPD) | 2 | 40 | 23 | 6 | 0.635 | 0.870 | 0.734 |
| **C17** | Baseline | MedGemma | IMPACT (Pulmonary / COPD) | 1 | 45 | 41 | 30 | 0.523 | 0.600 | 0.559 |
| **C18** | Baseline | MedGemma | IMPACT (Pulmonary / COPD) | 2 | 0 | 1 | 13 | 0 | 0 | 0 |
| **C19** | Baseline | MedGemma | vero(Bone / osteoporosis) | 1 | 2 | 3 | 13 | 0.400 | 0.133 | 0.200 |
| **C20** | Baseline | MedGemma | vero(Bone / osteoporosis) | 2 | 3 | 0 | 48 | 1 | 0.059 | 0.111 |

| | | | | | | | | | | |
|---|---|---|---|---|---|---|---|---|---|---|
| **C21** | Baseline | MedGemma | vero(Bone / osteoporosis) | 1 | 0 | 25 | 2 | 0 | 0 | 0 |
| **C22** | Baseline | MedGemma | Lead - 2 (Dibte) | 1 | 1 | 0 | 8 | 1 | 0.111 | 0.200 |
| **C23** | Baseline | MedGemma | Lead - 2 (Dibte) | 2 | 67 | 2 | 36 | 0.971 | 0.650 | 0.779 |
| **C24** | Baseline | MedGemma | Lead - 2 (Dibte) | 1 | 2 | 0 | 12 | 1 | 0.143 | 0.250 |
| **C25** | Baseline | MedGemma | Lead - 2 (Dibte) | 2 | 2 | 11 | 16 | 0.154 | 0.111 | 0.129 |
| **C26** | Baseline | MedGemma | Lead - 2 (Dibte) | 2 | 0 | 1 | 8 | 0 | 0 | 0 |
| **C27** | Baseline | MedGemma | Lead - 2 (Dibte) | 1 | 10 | 0 | 4 | 1 | 0.714 | 0.833 |
| **C28** | Baseline | MedGemma | AMPLIFY (Cardiovascula r / VTE) | 1 | 22 | 4 | 124 | 0.846 | 0.151 | 0.256 |
| **C29** | Baseline | MedGemma | AMPLIFY (Cardiovascula r / VTE) | 1 | 17 | 0 | 141 | 1 | 0.108 | 0.194 |
| **C30** | Baseline | MedGemma | AMPLIFY (Cardiovascula r / VTE) | 3 | 8 | 0 | 131 | 1 | 0.058 | 0.109 |
| **C31** | Baseline | MedGemma | AMPLIFY (Cardiovascula r / VTE) | 2 | 0 | 0 | 24 | 0 | 0 | 0 |
| **C32** | Baseline | MedGemma | AMPLIFY (Cardiovascula r / VTE) | 1 | 0 | 1 | 5 | 0 | 0 | 0 |
| **C33** | Baseline | MedGemma | ARISTOTLE | 2 | 129 | 4 | 288 | 0.970 | 0.309 | 0.469 |
| **C34** | Baseline | MedGemma | INSPIRE | 2 | 2 | 0 | 20 | 1 | 0.091 | 0.167 |
| **C35** | Baseline | MedGemma | leader | 1 | 3 | 1 | 89 | 0.750 | 0.033 | 0.062 |
| **C36** | Baseline | MedGemma | leader | 2 | 4 | 0 | 14 | 1 | 0.222 | 0.364 |
| **C37** | Baseline | MedGemma | leader | 2 | 11 | 3 | 29 | 0.786 | 0.275 | 0.407 |
| **C38** | Baseline | MedGemma | RECORD1 | 1 | 21 | 0 | 31 | 1 | 0.404 | 0.575 |
| **C39** | Baseline | MedGemma | PRONOUNCE _ | 3 | 20 | 10 | 131 | 0.667 | 0.132 | 0.221 |
| **C40** | Baseline | MedGemma | PRONOUNCE _ | 2 | 0 | 0 | 4 | 0 | 0 | 0 |
| **C01** | Hybrid biomedical RAG | Claude | DAPA-CKD(Kidney / cardio-renal) | 1 | 2 | 0 | 0 | 1 | 1 | 1 |
| **C02** | Hybrid biomedical RAG | Claude | DAPA-CKD(Kidney / cardio-renal) | 2 | 8 | 0 | 0 | 1 | 1 | 1 |
| **C03** | Hybrid biomedical RAG | Claude | DAPA-CKD(Kidney / cardio-renal) | 1 | 91 | 0 | 18 | 1 | 0.835 | 0.910 |
| **C04** | Hybrid biomedical RAG | Claude | DAPA-CKD(Kidney / | 2 | 5 | 0 | 2 | 1 | 0.714 | 0.833 |

| | | | | | | | | | | |
|---|---|---|---|---|---|---|---|---|---|---|
| | | | cardio-renal) | | | | | | | |
| **C05** | Hybrid biomedical RAG | Claude | DAPA-CKD(Kidney / cardio-renal) | 1 | 10 | 0 | 0 | 1 | 1 | 1 |
| **C06** | Hybrid biomedical RAG | Claude | DAPA-CKD(Kidney / cardio-renal) | 1 | 5 | 0 | 0 | 1 | 1 | 1 |
| **C07** | Hybrid biomedical RAG | Claude | ONTARGET | 1 | 4 | 0 | 0 | 1 | 1 | 1 |
| **C08** | Hybrid biomedical RAG | Claude | ONTARGET | 3 | 3 | 0 | 195 | 1 | 0.015 | 0.030 |
| **C09** | Hybrid biomedical RAG | Claude | ONTARGET | 3 | 0 | 2 | 135 | 0 | 0 | 0 |
| **C10** | Hybrid biomedical RAG | Claude | ONTARGET | 1 | 3 | 0 | 124 | 1 | 0.024 | 0.046 |
| **C11** | Hybrid biomedical RAG | Claude | ONTARGET | 1 | 18 | 29 | 1 | 0.383 | 0.947 | 0.545 |
| **C12** | Hybrid biomedical RAG | Claude | ONTARGET | 1 | 4 | 0 | 0 | 1 | 1 | 1 |
| **C13** | Hybrid biomedical RAG | Claude | ONTARGET | 1 | 6 | 0 | 0 | 1 | 1 | 1 |
| **C14** | Hybrid biomedical RAG | Claude | IMPACT (Pulmonary / COPD) | 1 | 20 | 0 | 4 | 1 | 0.833 | 0.909 |
| **C15** | Hybrid biomedical RAG | Claude | IMPACT (Pulmonary / COPD) | 1 | 2 | 0 | 0 | 1 | 1 | 1 |
| **C16** | Hybrid biomedical RAG | Claude | IMPACT (Pulmonary / COPD) | 2 | 33 | 12 | 13 | 0.733 | 0.717 | 0.725 |
| **C17** | Hybrid biomedical RAG | Claude | IMPACT (Pulmonary / COPD) | 1 | 70 | 0 | 5 | 1 | 0.933 | 0.966 |
| **C18** | Hybrid biomedical RAG | Claude | IMPACT (Pulmonary / COPD) | 2 | 3 | 4 | 10 | 0.429 | 0.231 | 0.300 |
| **C19** | Hybrid biomedical RAG | Claude | vero(Bone / osteoporosis) | 1 | 4 | 1 | 11 | 0.800 | 0.267 | 0.400 |
| **C20** | Hybrid biomedical RAG | Claude | vero(Bone / osteoporosis) | 2 | 8 | 2 | 43 | 0.800 | 0.157 | 0.262 |
| **C21** | Hybrid biomedical RAG | Claude | vero(Bone / osteoporosis) | 1 | 2 | 0 | 0 | 1 | 1 | 1 |
| **C22** | Hybrid biomedical RAG | Claude | Lead - 2 (Dibte) | 1 | 4 | 2 | 5 | 0.667 | 0.444 | 0.533 |
| **C23** | Hybrid biomedical RAG | Claude | Lead - 2 (Dibte) | 2 | 0 | 0 | 103 | 0 | 0 | 0 |
| **C24** | Hybrid biomedical RAG | Claude | Lead - 2 (Dibte) | 1 | 14 | 0 | 0 | 1 | 1 | 1 |
| **C25** | Hybrid biomedical RAG | Claude | Lead - 2 (Dibte) | 2 | 14 | 24 | 4 | 0.368 | 0.778 | 0.500 |

| | | | | | | | | | | |
|---|---|---|---|---|---|---|---|---|---|---|
| **C26** | Hybrid biomedical RAG | Claude | Lead - 2 (Dibte) | 2 | 3 | 2 | 5 | 0.600 | 0.375 | 0.462 |
| **C27** | Hybrid biomedical RAG | Claude | Lead - 2 (Dibte) | 1 | 14 | 2 | 0 | 0.875 | 1 | 0.933 |
| **C28** | Hybrid biomedical RAG | Claude | AMPLIFY (Cardiovascular / VTE) | 1 | 88 | 0 | 58 | 1 | 0.603 | 0.752 |
| **C29** | Hybrid biomedical RAG | Claude | AMPLIFY (Cardiovascular / VTE) | 1 | 38 | 243 | 120 | 0.135 | 0.241 | 0.173 |
| **C30** | Hybrid biomedical RAG | Claude | AMPLIFY (Cardiovascular / VTE) | 3 | 90 | 0 | 49 | 1 | 0.647 | 0.786 |
| **C31** | Hybrid biomedical RAG | Claude | AMPLIFY (Cardiovascular / VTE) | 2 | 2 | 12 | 22 | 0.143 | 0.083 | 0.105 |
| **C32** | Hybrid biomedical RAG | Claude | AMPLIFY (Cardiovascular / VTE) | 1 | 3 | 0 | 2 | 1 | 0.600 | 0.750 |
| **C33** | Hybrid biomedical RAG | Claude | ARISTOTLE | 2 | 90 | 0 | 327 | 1 | 0.216 | 0.355 |
| **C34** | Hybrid biomedical RAG | Claude | INSPIRE | 2 | 20 | 0 | 2 | 1 | 0.909 | 0.952 |
| **C35** | Hybrid biomedical RAG | Claude | leader | 1 | 29 | 9 | 63 | 0.763 | 0.315 | 0.446 |
| **C36** | Hybrid biomedical RAG | Claude | leader | 2 | 8 | 30 | 10 | 0.211 | 0.444 | 0.286 |
| **C37** | Hybrid biomedical RAG | Claude | leader | 2 | 13 | 0 | 27 | 1 | 0.325 | 0.491 |
| **C38** | Hybrid biomedical RAG | Claude | RECORD1 | 1 | 32 | 0 | 20 | 1 | 0.615 | 0.762 |
| **C39** | Hybrid biomedical RAG | Claude | PRONOUNCE_ | 3 | 140 | 31 | 11 | 0.819 | 0.927 | 0.870 |
| **C40** | Hybrid biomedical RAG | Claude | PRONOUNCE_ | 2 | 4 | 0 | 0 | 1 | 1 | 1 |
| **C01** | Hybrid biomedical RAG | GPT-5.5 | DAPA-CKD(Kidney / cardio-renal) | 1 | 2 | 0 | 0 | 1 | 1 | 1 |
| **C02** | Hybrid biomedical RAG | GPT-5.5 | DAPA-CKD(Kidney / cardio-renal) | 2 | 8 | 5 | 0 | 0.615 | 1 | 0.762 |
| **C03** | Hybrid biomedical RAG | GPT-5.5 | DAPA-CKD(Kidney / cardio-renal) | 1 | 76 | 0 | 33 | 1 | 0.697 | 0.822 |
| **C04** | Hybrid biomedical RAG | GPT-5.5 | DAPA-CKD(Kidney / cardio-renal) | 2 | 4 | 0 | 3 | 1 | 0.571 | 0.727 |
| **C05** | Hybrid biomedical RAG | GPT-5.5 | DAPA-CKD(Kidney / cardio-renal) | 1 | 10 | 0 | 0 | 1 | 1 | 1 |

| | | | | | | | | | | |
|---|---|---|---|---|---|---|---|---|---|---|
| **C06** | Hybrid biomedical RAG | GPT-5.5 | DAPA-CKD(Kidney / cardio-renal) | 1 | 4 | 1 | 1 | 0.800 | 0.800 | 0.800 |
| **C07** | Hybrid biomedical RAG | GPT-5.5 | ONTARGET | 1 | 4 | 0 | 0 | 1 | 1 | 1 |
| **C08** | Hybrid biomedical RAG | GPT-5.5 | ONTARGET | 3 | 8 | 161 | 190 | 0.047 | 0.040 | 0.044 |
| **C09** | Hybrid biomedical RAG | GPT-5.5 | ONTARGET | 3 | 132 | 72 | 3 | 0.647 | 0.978 | 0.779 |
| **C10** | Hybrid biomedical RAG | GPT-5.5 | ONTARGET | 1 | 113 | 232 | 14 | 0.328 | 0.890 | 0.479 |
| **C11** | Hybrid biomedical RAG | GPT-5.5 | ONTARGET | 1 | 19 | 0 | 0 | 1 | 1 | 1 |
| **C12** | Hybrid biomedical RAG | GPT-5.5 | ONTARGET | 1 | 4 | 0 | 0 | 1 | 1 | 1 |
| **C13** | Hybrid biomedical RAG | GPT-5.5 | ONTARGET | 1 | 5 | 0 | 1 | 1 | 0.833 | 0.909 |
| **C14** | Hybrid biomedical RAG | GPT-5.5 | IMPACT (Pulmonary / COPD) | 1 | 13 | 3 | 11 | 0.812 | 0.542 | 0.650 |
| **C15** | Hybrid biomedical RAG | GPT-5.5 | IMPACT (Pulmonary / COPD) | 1 | 2 | 2 | 0 | 0.500 | 1 | 0.667 |
| **C16** | Hybrid biomedical RAG | GPT-5.5 | IMPACT (Pulmonary / COPD) | 2 | 39 | 19 | 7 | 0.672 | 0.848 | 0.750 |
| **C17** | Hybrid biomedical RAG | GPT-5.5 | IMPACT (Pulmonary / COPD) | 1 | 73 | 76 | 2 | 0.490 | 0.973 | 0.652 |
| **C18** | Hybrid biomedical RAG | GPT-5.5 | IMPACT (Pulmonary / COPD) | 2 | 11 | 0 | 2 | 1 | 0.846 | 0.917 |
| **C19** | Hybrid biomedical RAG | GPT-5.5 | vero(Bone / osteoporosis) | 1 | 10 | 2 | 5 | 0.833 | 0.667 | 0.741 |
| **C20** | Hybrid biomedical RAG | GPT-5.5 | vero(Bone / osteoporosis) | 2 | 20 | 18 | 31 | 0.526 | 0.392 | 0.449 |
| **C21** | Hybrid biomedical RAG | GPT-5.5 | vero(Bone / osteoporosis) | 1 | 2 | 0 | 0 | 1 | 1 | 1 |
| **C22** | Hybrid biomedical RAG | GPT-5.5 | Lead - 2 (Dibte) | 1 | 4 | 0 | 5 | 1 | 0.444 | 0.615 |
| **C23** | Hybrid biomedical RAG | GPT-5.5 | Lead - 2 (Dibte) | 2 | 72 | 2 | 31 | 0.973 | 0.699 | 0.814 |
| **C24** | Hybrid biomedical RAG | GPT-5.5 | Lead - 2 (Dibte) | 1 | 14 | 1 | 0 | 0.933 | 1 | 0.966 |
| **C25** | Hybrid biomedical RAG | GPT-5.5 | Lead - 2 (Dibte) | 2 | 15 | 7 | 3 | 0.682 | 0.833 | 0.750 |
| **C26** | Hybrid biomedical RAG | GPT-5.5 | Lead - 2 (Dibte) | 2 | 6 | 0 | 2 | 1 | 0.750 | 0.857 |
| **C27** | Hybrid biomedical RAG | GPT-5.5 | Lead - 2 (Dibte) | 1 | 14 | 0 | 0 | 1 | 1 | 1 |

| | | | | | | | | | | |
|---|---|---|---|---|---|---|---|---|---|---|
| **C28** | Hybrid biomedical RAG | GPT-5.5 | AMPLIFY (Cardiovascular / VTE) | 1 | 130 | 43 | 16 | 0.751 | 0.890 | 0.815 |
| **C29** | Hybrid biomedical RAG | GPT-5.5 | AMPLIFY (Cardiovascular / VTE) | 1 | 38 | 424 | 120 | 0.082 | 0.241 | 0.123 |
| **C30** | Hybrid biomedical RAG | GPT-5.5 | AMPLIFY (Cardiovascular / VTE) | 3 | 136 | 76 | 3 | 0.642 | 0.978 | 0.775 |
| **C31** | Hybrid biomedical RAG | GPT-5.5 | AMPLIFY (Cardiovascular / VTE) | 2 | 6 | 4 | 18 | 0.600 | 0.250 | 0.353 |
| **C32** | Hybrid biomedical RAG | GPT-5.5 | AMPLIFY (Cardiovascular / VTE) | 1 | 5 | 0 | 0 | 1 | 1 | 1 |
| **C33** | Hybrid biomedical RAG | GPT-5.5 | ARISTOTLE | 2 | 105 | 17 | 312 | 0.861 | 0.252 | 0.390 |
| **C34** | Hybrid biomedical RAG | GPT-5.5 | INSPIRE | 2 | 19 | 0 | 3 | 1 | 0.864 | 0.927 |
| **C35** | Hybrid biomedical RAG | GPT-5.5 | leader | 1 | 92 | 21 | 0 | 0.814 | 1 | 0.898 |
| **C36** | Hybrid biomedical RAG | GPT-5.5 | leader | 2 | 8 | 32 | 10 | 0.200 | 0.444 | 0.276 |
| **C37** | Hybrid biomedical RAG | GPT-5.5 | leader | 2 | 40 | 1 | 0 | 0.976 | 1 | 0.988 |
| **C38** | Hybrid biomedical RAG | GPT-5.5 | RECORD1 | 1 | 52 | 0 | 0 | 1 | 1 | 1 |
| **C39** | Hybrid biomedical RAG | GPT-5.5 | PRONOUNCE_ | 3 | 146 | 37 | 5 | 0.798 | 0.967 | 0.874 |
| **C40** | Hybrid biomedical RAG | GPT-5.5 | PRONOUNCE_ | 2 | 4 | 0 | 0 | 1 | 1 | 1 |
| **C01** | Hybrid biomedical RAG | Qwen | DAPA-CKD(Kidney / cardio-renal) | 1 | 2 | 0 | 0 | 1 | 1 | 1 |
| **C02** | Hybrid biomedical RAG | Qwen | DAPA-CKD(Kidney / cardio-renal) | 2 | 7 | 0 | 1 | 1 | 0.875 | 0.933 |
| **C03** | Hybrid biomedical RAG | Qwen | DAPA-CKD(Kidney / cardio-renal) | 1 | 48 | 6 | 61 | 0.889 | 0.440 | 0.589 |
| **C04** | Hybrid biomedical RAG | Qwen | DAPA-CKD(Kidney / cardio-renal) | 2 | 5 | 0 | 2 | 1 | 0.714 | 0.833 |
| **C05** | Hybrid biomedical RAG | Qwen | DAPA-CKD(Kidney / cardio-renal) | 1 | 10 | 4 | 0 | 0.714 | 1 | 0.833 |
| **C06** | Hybrid biomedical RAG | Qwen | DAPA-CKD(Kidney / cardio-renal) | 1 | 2 | 0 | 3 | 1 | 0.400 | 0.571 |
| **C07** | Hybrid | Qwen | ONTARGET | 1 | 4 | 0 | 0 | 1 | 1 | 1 |

| | | | | | | | | | | |
|---|---|---|---|---|---|---|---|---|---|---|
| | biomedical RAG | | | | | | | | | |
| **C08** | Hybrid biomedical RAG | Qwen | ONTARGET | 3 | 4 | 166 | 194 | 0.024 | 0.020 | 0.022 |
| **C09** | Hybrid biomedical RAG | Qwen | ONTARGET | 3 | 121 | 4 | 14 | 0.968 | 0.896 | 0.931 |
| **C10** | Hybrid biomedical RAG | Qwen | ONTARGET | 1 | 27 | 2 | 100 | 0.931 | 0.213 | 0.346 |
| **C11** | Hybrid biomedical RAG | Qwen | ONTARGET | 1 | 19 | 0 | 0 | 1 | 1 | 1 |
| **C12** | Hybrid biomedical RAG | Qwen | ONTARGET | 1 | 1 | 0 | 3 | 1 | 0.250 | 0.400 |
| **C13** | Hybrid biomedical RAG | Qwen | ONTARGET | 1 | 2 | 0 | 4 | 1 | 0.333 | 0.500 |
| **C14** | Hybrid biomedical RAG | Qwen | IMPACT (Pulmonary / COPD) | 1 | 8 | 0 | 16 | 1 | 0.333 | 0.500 |
| **C15** | Hybrid biomedical RAG | Qwen | IMPACT (Pulmonary / COPD) | 1 | 2 | 0 | 0 | 1 | 1 | 1 |
| **C16** | Hybrid biomedical RAG | Qwen | IMPACT (Pulmonary / COPD) | 2 | 32 | 12 | 14 | 0.727 | 0.696 | 0.711 |
| **C17** | Hybrid biomedical RAG | Qwen | IMPACT (Pulmonary / COPD) | 1 | 51 | 54 | 24 | 0.486 | 0.680 | 0.567 |
| **C18** | Hybrid biomedical RAG | Qwen | IMPACT (Pulmonary / COPD) | 2 | 8 | 0 | 5 | 1 | 0.615 | 0.762 |
| **C19** | Hybrid biomedical RAG | Qwen | vero(Bone / osteoporosis) | 1 | 9 | 2 | 6 | 0.818 | 0.600 | 0.692 |
| **C20** | Hybrid biomedical RAG | Qwen | vero(Bone / osteoporosis) | 2 | 11 | 12 | 40 | 0.478 | 0.216 | 0.297 |
| **C21** | Hybrid biomedical RAG | Qwen | vero(Bone / osteoporosis) | 1 | 2 | 1 | 0 | 0.667 | 1 | 0.800 |
| **C22** | Hybrid biomedical RAG | Qwen | Lead - 2 (Dibte) | 1 | 1 | 1 | 8 | 0.500 | 0.111 | 0.182 |
| **C23** | Hybrid biomedical RAG | Qwen | Lead - 2 (Dibte) | 2 | 37 | 7 | 66 | 0.841 | 0.359 | 0.503 |
| **C24** | Hybrid biomedical RAG | Qwen | Lead - 2 (Dibte) | 1 | 4 | 1 | 10 | 0.800 | 0.286 | 0.421 |
| **C25** | Hybrid biomedical RAG | Qwen | Lead - 2 (Dibte) | 2 | 17 | 36 | 1 | 0.321 | 0.944 | 0.479 |
| **C26** | Hybrid biomedical RAG | Qwen | Lead - 2 (Dibte) | 2 | 1 | 0 | 7 | 1 | 0.125 | 0.222 |
| **C27** | Hybrid biomedical RAG | Qwen | Lead - 2 (Dibte) | 1 | 14 | 2 | 0 | 0.875 | 1 | 0.933 |
| **C28** | Hybrid biomedical RAG | Qwen | AMPLIFY (Cardiovascula r / VTE) | 1 | 44 | 6 | 102 | 0.880 | 0.301 | 0.449 |
| **C29** | Hybrid | Qwen | AMPLIFY | 1 | 38 | 129 | 120 | 0.228 | 0.241 | 0.234 |

| | | | | | | | | | | |
|---|---|---|---|---|---|---|---|---|---|---|
| | biomedical RAG | | (Cardiovascula r / VTE) | | | | | | | |
| **C30** | Hybrid biomedical RAG | Qwen | AMPLIFY (Cardiovascula r / VTE) | 3 | 107 | 6 | 32 | 0.947 | 0.770 | 0.849 |
| **C31** | Hybrid biomedical RAG | Qwen | AMPLIFY (Cardiovascula r / VTE) | 2 | 4 | 0 | 20 | 1 | 0.167 | 0.286 |
| **C32** | Hybrid biomedical RAG | Qwen | AMPLIFY (Cardiovascula r / VTE) | 1 | 3 | 0 | 2 | 1 | 0.600 | 0.750 |
| **C33** | Hybrid biomedical RAG | Qwen | ARISTOTLE | 2 | 159 | 4 | 258 | 0.975 | 0.381 | 0.548 |
| **C34** | Hybrid biomedical RAG | Qwen | INSPIRE | 2 | 19 | 0 | 3 | 1 | 0.864 | 0.927 |
| **C35** | Hybrid biomedical RAG | Qwen | leader | 1 | 91 | 12 | 1 | 0.883 | 0.989 | 0.933 |
| **C36** | Hybrid biomedical RAG | Qwen | leader | 2 | 8 | 29 | 10 | 0.216 | 0.444 | 0.291 |
| **C37** | Hybrid biomedical RAG | Qwen | leader | 2 | 23 | 0 | 17 | 1 | 0.575 | 0.730 |
| **C38** | Hybrid biomedical RAG | Qwen | RECORD1 | 1 | 32 | 0 | 20 | 1 | 0.615 | 0.762 |
| **C39** | Hybrid biomedical RAG | Qwen | PRONOUNCE_ | 3 | 100 | 68 | 51 | 0.595 | 0.662 | 0.627 |
| **C40** | Hybrid biomedical RAG | Qwen | PRONOUNCE_ | 2 | 3 | 3 | 1 | 0.500 | 0.750 | 0.600 |
| **C01** | Hybrid biomedical RAG | MedGemma | DAPA-CKD(Kidney / cardio-renal) | 1 | 2 | 9 | 0 | 0.182 | 1 | 0.308 |
| **C02** | Hybrid biomedical RAG | MedGemma | DAPA-CKD(Kidney / cardio-renal) | 2 | 7 | 1 | 1 | 0.875 | 0.875 | 0.875 |
| **C03** | Hybrid biomedical RAG | MedGemma | DAPA-CKD(Kidney / cardio-renal) | 1 | 2 | 0 | 107 | 1 | 0.018 | 0.036 |
| **C04** | Hybrid biomedical RAG | MedGemma | DAPA-CKD(Kidney / cardio-renal) | 2 | 4 | 0 | 3 | 1 | 0.571 | 0.727 |
| **C05** | Hybrid biomedical RAG | MedGemma | DAPA-CKD(Kidney / cardio-renal) | 1 | 10 | 0 | 0 | 1 | 1 | 1 |
| **C06** | Hybrid biomedical RAG | MedGemma | DAPA-CKD(Kidney / cardio-renal) | 1 | 1 | 2 | 4 | 0.333 | 0.200 | 0.250 |
| **C07** | Hybrid biomedical RAG | MedGemma | ONTARGET | 1 | 2 | 0 | 2 | 1 | 0.500 | 0.667 |
| **C08** | Hybrid biomedical RAG | MedGemma | ONTARGET | 3 | 0 | 0 | 198 | 0 | 0 | 0 |
| **C09** | Hybrid | MedGemma | ONTARGET | 3 | 5 | 20 | 130 | 0.200 | 0.037 | 0.062 |

| | | | | | | | | | | |
|---|---|---|---|---|---|---|---|---|---|---|
| | biomedical RAG | | | | | | | | | |
| **C10** | Hybrid biomedical RAG | MedGemma | ONTARGET | 1 | 3 | 36 | 124 | 0.077 | 0.024 | 0.036 |
| **C11** | Hybrid biomedical RAG | MedGemma | ONTARGET | 1 | 19 | 3 | 0 | 0.864 | 1 | 0.927 |
| **C12** | Hybrid biomedical RAG | MedGemma | ONTARGET | 1 | 4 | 23 | 0 | 0.148 | 1 | 0.258 |
| **C13** | Hybrid biomedical RAG | MedGemma | ONTARGET | 1 | 3 | 6 | 3 | 0.333 | 0.500 | 0.400 |
| **C14** | Hybrid biomedical RAG | MedGemma | IMPACT (Pulmonary / COPD) | 1 | 7 | 0 | 17 | 1 | 0.292 | 0.452 |
| **C15** | Hybrid biomedical RAG | MedGemma | IMPACT (Pulmonary / COPD) | 1 | 2 | 14 | 0 | 0.125 | 1 | 0.222 |
| **C16** | Hybrid biomedical RAG | MedGemma | IMPACT (Pulmonary / COPD) | 2 | 39 | 23 | 7 | 0.629 | 0.848 | 0.722 |
| **C17** | Hybrid biomedical RAG | MedGemma | IMPACT (Pulmonary / COPD) | 1 | 30 | 22 | 45 | 0.577 | 0.400 | 0.472 |
| **C18** | Hybrid biomedical RAG | MedGemma | IMPACT (Pulmonary / COPD) | 2 | 13 | 23 | 0 | 0.361 | 1 | 0.531 |
| **C19** | Hybrid biomedical RAG | MedGemma | vero(Bone / osteoporosis) | 1 | 4 | 9 | 11 | 0.308 | 0.267 | 0.286 |
| **C20** | Hybrid biomedical RAG | MedGemma | vero(Bone / osteoporosis) | 2 | 6 | 1 | 45 | 0.857 | 0.118 | 0.207 |
| **C21** | Hybrid biomedical RAG | MedGemma | vero(Bone / osteoporosis) | 1 | 2 | 25 | 0 | 0.074 | 1 | 0.138 |
| **C22** | Hybrid biomedical RAG | MedGemma | Lead - 2 (Dibte) | 1 | 2 | 18 | 7 | 0.100 | 0.222 | 0.138 |
| **C23** | Hybrid biomedical RAG | MedGemma | Lead - 2 (Dibte) | 2 | 11 | 8 | 92 | 0.579 | 0.107 | 0.180 |
| **C24** | Hybrid biomedical RAG | MedGemma | Lead - 2 (Dibte) | 1 | 2 | 0 | 12 | 1 | 0.143 | 0.250 |
| **C25** | Hybrid biomedical RAG | MedGemma | Lead - 2 (Dibte) | 2 | 9 | 26 | 9 | 0.257 | 0.500 | 0.340 |
| **C26** | Hybrid biomedical RAG | MedGemma | Lead - 2 (Dibte) | 2 | 0 | 1 | 8 | 0 | 0 | 0 |
| **C27** | Hybrid biomedical RAG | MedGemma | Lead - 2 (Dibte) | 1 | 10 | 0 | 4 | 1 | 0.714 | 0.833 |
| **C28** | Hybrid biomedical RAG | MedGemma | AMPLIFY (Cardiovascular / VTE) | 1 | 111 | 15 | 35 | 0.881 | 0.760 | 0.816 |
| **C29** | Hybrid biomedical RAG | MedGemma | AMPLIFY (Cardiovascular / VTE) | 1 | 146 | 18 | 12 | 0.890 | 0.924 | 0.907 |
| **C30** | Hybrid biomedical RAG | MedGemma | AMPLIFY (Cardiovascula | 3 | 36 | 10 | 103 | 0.783 | 0.259 | 0.389 |

| | | | | | | | | | | |
|---|---|---|---|---|---|---|---|---|---|---|
| | | | r / VTE) | | | | | | | |
| **C31** | Hybrid biomedical RAG | MedGemma | AMPLIFY (Cardiovascular / VTE) | 2 | 1 | 14 | 23 | 0.067 | 0.042 | 0.051 |
| **C32** | Hybrid biomedical RAG | MedGemma | AMPLIFY (Cardiovascular / VTE) | 1 | 2 | 3 | 3 | 0.400 | 0.400 | 0.400 |
| **C33** | Hybrid biomedical RAG | MedGemma | ARISTOTLE | 2 | 20 | 0 | 397 | 1 | 0.048 | 0.092 |
| **C34** | Hybrid biomedical RAG | MedGemma | INSPIRE | 2 | 4 | 0 | 18 | 1 | 0.182 | 0.308 |
| **C35** | Hybrid biomedical RAG | MedGemma | leader | 1 | 1 | 9 | 91 | 0.100 | 0.011 | 0.020 |
| **C36** | Hybrid biomedical RAG | MedGemma | leader | 2 | 4 | 0 | 14 | 1 | 0.222 | 0.364 |
| **C37** | Hybrid biomedical RAG | MedGemma | leader | 2 | 3 | 28 | 37 | 0.097 | 0.075 | 0.085 |
| **C38** | Hybrid biomedical RAG | MedGemma | RECORD1 | 1 | 34 | 0 | 18 | 1 | 0.654 | 0.791 |
| **C39** | Hybrid biomedical RAG | MedGemma | PRONOUNCE _ | 3 | 7 | 13 | 144 | 0.350 | 0.046 | 0.082 |
| **C40** | Hybrid biomedical RAG | MedGemma | PRONOUNCE _ | 2 | 0 | 3 | 4 | 0 | 0 | 0 |
| **C01** | Family expansion | Claude | DAPA-CKD(Kidney / cardio-renal) | 1 | 2 | 0 | 0 | 1 | 1 | 1 |
| **C02** | Family expansion | Claude | DAPA-CKD(Kidney / cardio-renal) | 2 | 7 | 0 | 1 | 1 | 0.875 | 0.933 |
| **C03** | Family expansion | Claude | DAPA-CKD(Kidney / cardio-renal) | 1 | 87 | 12 | 22 | 0.879 | 0.798 | 0.837 |
| **C04** | Family expansion | Claude | DAPA-CKD(Kidney / cardio-renal) | 2 | 6 | 0 | 1 | 1 | 0.857 | 0.923 |
| **C05** | Family expansion | Claude | DAPA-CKD(Kidney / cardio-renal) | 1 | 10 | 1 | 0 | 0.909 | 1 | 0.952 |
| **C06** | Family expansion | Claude | DAPA-CKD(Kidney / cardio-renal) | 1 | 1 | 0 | 4 | 1 | 0.200 | 0.333 |
| **C07** | Family expansion | Claude | ONTARGET | 1 | 4 | 0 | 0 | 1 | 1 | 1 |
| **C08** | Family expansion | Claude | ONTARGET | 3 | 155 | 415 | 43 | 0.272 | 0.783 | 0.404 |
| **C09** | Family expansion | Claude | ONTARGET | 3 | 26 | 100 | 109 | 0.206 | 0.193 | 0.199 |
| **C10** | Family expansion | Claude | ONTARGET | 1 | 45 | 248 | 82 | 0.154 | 0.354 | 0.214 |
| **C11** | Family | Claude | ONTARGET | 1 | 19 | 37 | 0 | 0.339 | 1 | 0.507 |

| | | | | | | | | | | |
|---|---|---|---|---|---|---|---|---|---|---|
| | expansion | | | | | | | | | |
| **C12** | Family expansion | Claude | ONTARGET | 1 | 0 | 1 | 4 | 0 | 0 | 0 |
| **C13** | Family expansion | Claude | ONTARGET | 1 | 1 | 0 | 5 | 1 | 0.167 | 0.286 |
| **C14** | Family expansion | Claude | IMPACT (Pulmonary / COPD) | 1 | 21 | 22 | 3 | 0.488 | 0.875 | 0.627 |
| **C15** | Family expansion | Claude | IMPACT (Pulmonary / COPD) | 1 | 2 | 0 | 0 | 1 | 1 | 1 |
| **C16** | Family expansion | Claude | IMPACT (Pulmonary / COPD) | 2 | 39 | 11 | 7 | 0.780 | 0.848 | 0.812 |
| **C17** | Family expansion | Claude | IMPACT (Pulmonary / COPD) | 1 | 72 | 0 | 3 | 1 | 0.960 | 0.980 |
| **C18** | Family expansion | Claude | IMPACT (Pulmonary / COPD) | 2 | 0 | 0 | 13 | 0 | 0 | 0 |
| **C19** | Family expansion | Claude | vero(Bone / osteoporosis) | 1 | 10 | 1 | 5 | 0.909 | 0.667 | 0.769 |
| **C20** | Family expansion | Claude | vero(Bone / osteoporosis) | 2 | 13 | 7 | 38 | 0.650 | 0.255 | 0.366 |
| **C21** | Family expansion | Claude | vero(Bone / osteoporosis) | 1 | 2 | 3 | 0 | 0.400 | 1 | 0.571 |
| **C22** | Family expansion | Claude | Lead - 2 (Dibte) | 1 | 4 | 2 | 5 | 0.667 | 0.444 | 0.533 |
| **C23** | Family expansion | Claude | Lead - 2 (Dibte) | 2 | 47 | 0 | 56 | 1 | 0.456 | 0.627 |
| **C24** | Family expansion | Claude | Lead - 2 (Dibte) | 1 | 3 | 0 | 11 | 1 | 0.214 | 0.353 |
| **C25** | Family expansion | Claude | Lead - 2 (Dibte) | 2 | 17 | 36 | 1 | 0.321 | 0.944 | 0.479 |
| **C26** | Family expansion | Claude | Lead - 2 (Dibte) | 2 | 2 | 0 | 6 | 1 | 0.250 | 0.400 |
| **C27** | Family expansion | Claude | Lead - 2 (Dibte) | 1 | 14 | 0 | 0 | 1 | 1 | 1 |
| **C28** | Family expansion | Claude | AMPLIFY (Cardiovascular / VTE) | 1 | 141 | 78 | 5 | 0.644 | 0.966 | 0.773 |
| **C29** | Family expansion | Claude | AMPLIFY (Cardiovascular / VTE) | 1 | 38 | 180 | 120 | 0.174 | 0.241 | 0.202 |
| **C30** | Family expansion | Claude | AMPLIFY (Cardiovascular / VTE) | 3 | 128 | 25 | 11 | 0.837 | 0.921 | 0.877 |
| **C31** | Family expansion | Claude | AMPLIFY (Cardiovascular / VTE) | 2 | 13 | 13 | 11 | 0.500 | 0.542 | 0.520 |

| | | | | | | | | | | |
|---|---|---|---|---|---|---|---|---|---|---|
| **C32** | Family expansion | Claude | AMPLIFY (Cardiovascular / VTE) | 1 | 1 | 0 | 4 | 1 | 0.200 | 0.333 |
| **C33** | Family expansion | Claude | ARISTOTLE | 2 | 330 | 173 | 87 | 0.656 | 0.791 | 0.717 |
| **C34** | Family expansion | Claude | INSPIRE | 2 | 15 | 0 | 7 | 1 | 0.682 | 0.811 |
| **C35** | Family expansion | Claude | leader | 1 | 78 | 13 | 14 | 0.857 | 0.848 | 0.852 |
| **C36** | Family expansion | Claude | leader | 2 | 8 | 28 | 10 | 0.222 | 0.444 | 0.296 |
| **C37** | Family expansion | Claude | leader | 2 | 30 | 18 | 10 | 0.625 | 0.750 | 0.682 |
| **C38** | Family expansion | Claude | RECORD1 | 1 | 29 | 0 | 23 | 1 | 0.558 | 0.716 |
| **C39** | Family expansion | Claude | PRONOUNCE_ | 3 | 133 | 29 | 18 | 0.821 | 0.881 | 0.850 |
| **C40** | Family expansion | Claude | PRONOUNCE_ | 2 | 4 | 0 | 0 | 1 | 1 | 1 |
| **C01** | Family expansion | GPT-5.5 | DAPA-CKD(Kidney / cardio-renal) | 1 | 2 | 1 | 0 | 0.667 | 1 | 0.800 |
| **C02** | Family expansion | GPT-5.5 | DAPA-CKD(Kidney / cardio-renal) | 2 | 8 | 9 | 0 | 0.471 | 1 | 0.640 |
| **C03** | Family expansion | GPT-5.5 | DAPA-CKD(Kidney / cardio-renal) | 1 | 107 | 341 | 2 | 0.239 | 0.982 | 0.384 |
| **C04** | Family expansion | GPT-5.5 | DAPA-CKD(Kidney / cardio-renal) | 2 | 7 | 13 | 0 | 0.350 | 1 | 0.519 |
| **C05** | Family expansion | GPT-5.5 | DAPA-CKD(Kidney / cardio-renal) | 1 | 10 | 22 | 0 | 0.312 | 1 | 0.476 |
| **C06** | Family expansion | GPT-5.5 | DAPA-CKD(Kidney / cardio-renal) | 1 | 1 | 0 | 4 | 1 | 0.200 | 0.333 |
| **C07** | Family expansion | GPT-5.5 | ONTARGET | 1 | 4 | 154 | 0 | 0.025 | 1 | 0.049 |
| **C08** | Family expansion | GPT-5.5 | ONTARGET | 3 | 53 | 593 | 145 | 0.082 | 0.268 | 0.126 |
| **C09** | Family expansion | GPT-5.5 | ONTARGET | 3 | 133 | 467 | 2 | 0.222 | 0.985 | 0.362 |
| **C10** | Family expansion | GPT-5.5 | ONTARGET | 1 | 44 | 231 | 83 | 0.160 | 0.346 | 0.219 |
| **C11** | Family expansion | GPT-5.5 | ONTARGET | 1 | 19 | 37 | 0 | 0.339 | 1 | 0.507 |
| **C12** | Family expansion | GPT-5.5 | ONTARGET | 1 | 0 | 1 | 4 | 0 | 0 | 0 |
| **C13** | Family | GPT-5.5 | ONTARGET | 1 | 1 | 0 | 5 | 1 | 0.167 | 0.286 |

| | | | | | | | | | | |
|---|---|---|---|---|---|---|---|---|---|---|
| | expansion | | | | | | | | | |
| **C14** | Family expansion | GPT-5.5 | IMPACT (Pulmonary / COPD) | 1 | 22 | 3 | 2 | 0.880 | 0.917 | 0.898 |
| **C15** | Family expansion | GPT-5.5 | IMPACT (Pulmonary / COPD) | 1 | 2 | 6 | 0 | 0.250 | 1 | 0.400 |
| **C16** | Family expansion | GPT-5.5 | IMPACT (Pulmonary / COPD) | 2 | 39 | 19 | 7 | 0.672 | 0.848 | 0.750 |
| **C17** | Family expansion | GPT-5.5 | IMPACT (Pulmonary / COPD) | 1 | 27 | 204 | 48 | 0.117 | 0.360 | 0.176 |
| **C18** | Family expansion | GPT-5.5 | IMPACT (Pulmonary / COPD) | 2 | 2 | 0 | 11 | 1 | 0.154 | 0.267 |
| **C19** | Family expansion | GPT-5.5 | vero(Bone / osteoporosis) | 1 | 4 | 1 | 11 | 0.800 | 0.267 | 0.400 |
| **C20** | Family expansion | GPT-5.5 | vero(Bone / osteoporosis) | 2 | 12 | 5 | 39 | 0.706 | 0.235 | 0.353 |
| **C21** | Family expansion | GPT-5.5 | vero(Bone / osteoporosis) | 1 | 2 | 8 | 0 | 0.200 | 1 | 0.333 |
| **C22** | Family expansion | GPT-5.5 | Lead - 2 (Dibte) | 1 | 4 | 2 | 5 | 0.667 | 0.444 | 0.533 |
| **C23** | Family expansion | GPT-5.5 | Lead - 2 (Dibte) | 2 | 90 | 12 | 13 | 0.882 | 0.874 | 0.878 |
| **C24** | Family expansion | GPT-5.5 | Lead - 2 (Dibte) | 1 | 4 | 30 | 10 | 0.118 | 0.286 | 0.167 |
| **C25** | Family expansion | GPT-5.5 | Lead - 2 (Dibte) | 2 | 16 | 21 | 2 | 0.432 | 0.889 | 0.582 |
| **C26** | Family expansion | GPT-5.5 | Lead - 2 (Dibte) | 2 | 2 | 0 | 6 | 1 | 0.250 | 0.400 |
| **C27** | Family expansion | GPT-5.5 | Lead - 2 (Dibte) | 1 | 14 | 119 | 0 | 0.105 | 1 | 0.190 |
| **C28** | Family expansion | GPT-5.5 | AMPLIFY (Cardiovascula r / VTE) | 1 | 130 | 42 | 16 | 0.756 | 0.890 | 0.818 |
| **C29** | Family expansion | GPT-5.5 | AMPLIFY (Cardiovascula r / VTE) | 1 | 40 | 356 | 118 | 0.101 | 0.253 | 0.144 |
| **C30** | Family expansion | GPT-5.5 | AMPLIFY (Cardiovascula r / VTE) | 3 | 131 | 119 | 8 | 0.524 | 0.942 | 0.674 |
| **C31** | Family expansion | GPT-5.5 | AMPLIFY (Cardiovascula r / VTE) | 2 | 14 | 100 | 10 | 0.123 | 0.583 | 0.203 |
| **C32** | Family expansion | GPT-5.5 | AMPLIFY (Cardiovascula r / VTE) | 1 | 3 | 0 | 2 | 1 | 0.600 | 0.750 |
| **C33** | Family | GPT-5.5 | ARISTOTLE | 2 | 366 | 5 | 51 | 0.987 | 0.878 | 0.929 |

| | | | | | | | | | | |
|---|---|---|---|---|---|---|---|---|---|---|
| | expansion | | | | | | | | | |
| **C34** | Family expansion | GPT-5.5 | INSPIRE | 2 | 11 | 0 | 11 | 1 | 0.500 | 0.667 |
| **C35** | Family expansion | GPT-5.5 | leader | 1 | 76 | 40 | 16 | 0.655 | 0.826 | 0.731 |
| **C36** | Family expansion | GPT-5.5 | leader | 2 | 6 | 71 | 12 | 0.078 | 0.333 | 0.126 |
| **C37** | Family expansion | GPT-5.5 | leader | 2 | 23 | 92 | 17 | 0.200 | 0.575 | 0.297 |
| **C38** | Family expansion | GPT-5.5 | RECORD1 | 1 | 51 | 28 | 1 | 0.646 | 0.981 | 0.779 |
| **C39** | Family expansion | GPT-5.5 | PRONOUNCE _ | 3 | 150 | 169 | 1 | 0.470 | 0.993 | 0.638 |
| **C40** | Family expansion | GPT-5.5 | PRONOUNCE _ | 2 | 4 | 7 | 0 | 0.364 | 1 | 0.533 |
| **C01** | Family expansion | Qwen | DAPA-CKD(Kidney / cardio-renal) | 1 | 1 | 2 | 1 | 0.333 | 0.500 | 0.400 |
| **C02** | Family expansion | Qwen | DAPA-CKD(Kidney / cardio-renal) | 2 | 8 | 0 | 0 | 1 | 1 | 1 |
| **C03** | Family expansion | Qwen | DAPA-CKD(Kidney / cardio-renal) | 1 | 107 | 0 | 2 | 1 | 0.982 | 0.991 |
| **C04** | Family expansion | Qwen | DAPA-CKD(Kidney / cardio-renal) | 2 | 3 | 0 | 4 | 1 | 0.429 | 0.600 |
| **C05** | Family expansion | Qwen | DAPA-CKD(Kidney / cardio-renal) | 1 | 10 | 0 | 0 | 1 | 1 | 1 |
| **C06** | Family expansion | Qwen | DAPA-CKD(Kidney / cardio-renal) | 1 | 0 | 0 | 5 | 0 | 0 | 0 |
| **C07** | Family expansion | Qwen | ONTARGET | 1 | 4 | 0 | 0 | 1 | 1 | 1 |
| **C08** | Family expansion | Qwen | ONTARGET | 3 | 4 | 0 | 194 | 1 | 0.020 | 0.040 |
| **C09** | Family expansion | Qwen | ONTARGET | 3 | 14 | 9 | 121 | 0.609 | 0.104 | 0.177 |
| **C10** | Family expansion | Qwen | ONTARGET | 1 | 24 | 226 | 103 | 0.096 | 0.189 | 0.127 |
| **C11** | Family expansion | Qwen | ONTARGET | 1 | 5 | 10 | 14 | 0.333 | 0.263 | 0.294 |
| **C12** | Family expansion | Qwen | ONTARGET | 1 | 0 | 0 | 4 | 0 | 0 | 0 |
| **C13** | Family expansion | Qwen | ONTARGET | 1 | 0 | 0 | 6 | 0 | 0 | 0 |
| **C14** | Family expansion | Qwen | IMPACT (Pulmonary / COPD) | 1 | 13 | 0 | 11 | 1 | 0.542 | 0.703 |

| | | | | | | | | | | |
|---|---|---|---|---|---|---|---|---|---|---|
| **C15** | Family expansion | Qwen | IMPACT (Pulmonary / COPD) | 1 | 1 | 0 | 1 | 1 | 0.500 | 0.667 |
| **C16** | Family expansion | Qwen | IMPACT (Pulmonary / COPD) | 2 | 38 | 13 | 8 | 0.745 | 0.826 | 0.784 |
| **C17** | Family expansion | Qwen | IMPACT (Pulmonary / COPD) | 1 | 66 | 545 | 9 | 0.108 | 0.880 | 0.192 |
| **C18** | Family expansion | Qwen | IMPACT (Pulmonary / COPD) | 2 | 0 | 0 | 13 | 0 | 0 | 0 |
| **C19** | Family expansion | Qwen | vero(Bone / osteoporosis) | 1 | 7 | 0 | 8 | 1 | 0.467 | 0.636 |
| **C20** | Family expansion | Qwen | vero(Bone / osteoporosis) | 2 | 11 | 5 | 40 | 0.688 | 0.216 | 0.328 |
| **C21** | Family expansion | Qwen | vero(Bone / osteoporosis) | 1 | 2 | 8 | 0 | 0.200 | 1 | 0.333 |
| **C22** | Family expansion | Qwen | Lead - 2 (Dibte) | 1 | 2 | 1 | 7 | 0.667 | 0.222 | 0.333 |
| **C23** | Family expansion | Qwen | Lead - 2 (Dibte) | 2 | 61 | 294 | 42 | 0.172 | 0.592 | 0.266 |
| **C24** | Family expansion | Qwen | Lead - 2 (Dibte) | 1 | 8 | 0 | 6 | 1 | 0.571 | 0.727 |
| **C25** | Family expansion | Qwen | Lead - 2 (Dibte) | 2 | 0 | 4 | 18 | 0 | 0 | 0 |
| **C26** | Family expansion | Qwen | Lead - 2 (Dibte) | 2 | 1 | 0 | 7 | 1 | 0.125 | 0.222 |
| **C27** | Family expansion | Qwen | Lead - 2 (Dibte) | 1 | 10 | 0 | 4 | 1 | 0.714 | 0.833 |
| **C28** | Family expansion | Qwen | AMPLIFY (Cardiovascula r / VTE) | 1 | 129 | 46 | 17 | 0.737 | 0.884 | 0.804 |
| **C29** | Family expansion | Qwen | AMPLIFY (Cardiovascula r / VTE) | 1 | 29 | 428 | 129 | 0.063 | 0.184 | 0.094 |
| **C30** | Family expansion | Qwen | AMPLIFY (Cardiovascula r / VTE) | 3 | 116 | 7 | 23 | 0.943 | 0.835 | 0.885 |
| **C31** | Family expansion | Qwen | AMPLIFY (Cardiovascula r / VTE) | 2 | 4 | 0 | 20 | 1 | 0.167 | 0.286 |
| **C32** | Family expansion | Qwen | AMPLIFY (Cardiovascula r / VTE) | 1 | 1 | 1 | 4 | 0.500 | 0.200 | 0.286 |
| **C33** | Family expansion | Qwen | ARISTOTLE | 2 | 344 | 5 | 73 | 0.986 | 0.825 | 0.898 |
| **C34** | Family expansion | Qwen | INSPIRE | 2 | 11 | 0 | 11 | 1 | 0.500 | 0.667 |
| **C35** | Family | Qwen | leader | 1 | 42 | 10 | 50 | 0.808 | 0.457 | 0.583 |

| | | | | | | | | | | |
|---|---|---|---|---|---|---|---|---|---|---|
| | expansion | | | | | | | | | |
| **C36** | Family expansion | Qwen | leader | 2 | 0 | 16 | 18 | 0 | 0 | 0 |
| **C37** | Family expansion | Qwen | leader | 2 | 20 | 0 | 20 | 1 | 0.500 | 0.667 |
| **C38** | Family expansion | Qwen | RECORD1 | 1 | 38 | 0 | 14 | 1 | 0.731 | 0.844 |
| **C39** | Family expansion | Qwen | PRONOUNCE_ | 3 | 137 | 31 | 14 | 0.815 | 0.907 | 0.859 |
| **C40** | Family expansion | Qwen | PRONOUNCE_ | 2 | 3 | 0 | 1 | 1 | 0.750 | 0.857 |
| **C01** | Family expansion | MedGemma | DAPA-CKD(Kidney / cardio-renal) | 1 | 1 | 0 | 1 | 1 | 0.500 | 0.667 |
| **C02** | Family expansion | MedGemma | DAPA-CKD(Kidney / cardio-renal) | 2 | 5 | 2 | 3 | 0.714 | 0.625 | 0.667 |
| **C03** | Family expansion | MedGemma | DAPA-CKD(Kidney / cardio-renal) | 1 | 87 | 28 | 22 | 0.757 | 0.798 | 0.777 |
| **C04** | Family expansion | MedGemma | DAPA-CKD(Kidney / cardio-renal) | 2 | 0 | 12 | 7 | 0 | 0 | 0 |
| **C05** | Family expansion | MedGemma | DAPA-CKD(Kidney / cardio-renal) | 1 | 5 | 7 | 5 | 0.417 | 0.500 | 0.455 |
| **C06** | Family expansion | MedGemma | DAPA-CKD(Kidney / cardio-renal) | 1 | 0 | 0 | 5 | 0 | 0 | 0 |
| **C07** | Family expansion | MedGemma | ONTARGET | 1 | 1 | 13 | 3 | 0.071 | 0.250 | 0.111 |
| **C08** | Family expansion | MedGemma | ONTARGET | 3 | 2 | 43 | 196 | 0.044 | 0.010 | 0.016 |
| **C09** | Family expansion | MedGemma | ONTARGET | 3 | 122 | 92 | 13 | 0.570 | 0.904 | 0.699 |
| **C10** | Family expansion | MedGemma | ONTARGET | 1 | 0 | 22 | 127 | 0 | 0 | 0 |
| **C11** | Family expansion | MedGemma | ONTARGET | 1 | 18 | 0 | 1 | 1 | 0.947 | 0.973 |
| **C12** | Family expansion | MedGemma | ONTARGET | 1 | 0 | 0 | 4 | 0 | 0 | 0 |
| **C13** | Family expansion | MedGemma | ONTARGET | 1 | 0 | 0 | 6 | 0 | 0 | 0 |
| **C14** | Family expansion | MedGemma | IMPACT (Pulmonary / COPD) | 1 | 4 | 1 | 20 | 0.800 | 0.167 | 0.276 |
| **C15** | Family expansion | MedGemma | IMPACT (Pulmonary / COPD) | 1 | 1 | 1 | 1 | 0.500 | 0.500 | 0.500 |
| **C16** | Family | MedGemma | IMPACT | 2 | 24 | 10 | 22 | 0.706 | 0.522 | 0.600 |

| | | | | | | | | | | |
|---|---|---|---|---|---|---|---|---|---|---|
| | expansion | | (Pulmonary / COPD) | | | | | | | |
| **C17** | Family expansion | MedGemma | IMPACT (Pulmonary / COPD) | 1 | 48 | 16 | 27 | 0.750 | 0.640 | 0.691 |
| **C18** | Family expansion | MedGemma | IMPACT (Pulmonary / COPD) | 2 | 0 | 0 | 13 | 0 | 0 | 0 |
| **C19** | Family expansion | MedGemma | vero(Bone / osteoporosis) | 1 | 2 | 3 | 13 | 0.400 | 0.133 | 0.200 |
| **C20** | Family expansion | MedGemma | vero(Bone / osteoporosis) | 2 | 6 | 15 | 45 | 0.286 | 0.118 | 0.167 |
| **C21** | Family expansion | MedGemma | vero(Bone / osteoporosis) | 1 | 0 | 0 | 2 | 0 | 0 | 0 |
| **C22** | Family expansion | MedGemma | Lead - 2 (Dibte) | 1 | 1 | 0 | 8 | 1 | 0.111 | 0.200 |
| **C23** | Family expansion | MedGemma | Lead - 2 (Dibte) | 2 | 23 | 0 | 80 | 1 | 0.223 | 0.365 |
| **C24** | Family expansion | MedGemma | Lead - 2 (Dibte) | 1 | 2 | 7 | 12 | 0.222 | 0.143 | 0.174 |
| **C25** | Family expansion | MedGemma | Lead - 2 (Dibte) | 2 | 2 | 14 | 16 | 0.125 | 0.111 | 0.118 |
| **C26** | Family expansion | MedGemma | Lead - 2 (Dibte) | 2 | 0 | 0 | 8 | 0 | 0 | 0 |
| **C27** | Family expansion | MedGemma | Lead - 2 (Dibte) | 1 | 8 | 338 | 6 | 0.023 | 0.571 | 0.044 |
| **C28** | Family expansion | MedGemma | AMPLIFY (Cardiovascula r / VTE) | 1 | 24 | 1 | 122 | 0.960 | 0.164 | 0.281 |
| **C29** | Family expansion | MedGemma | AMPLIFY (Cardiovascula r / VTE) | 1 | 3 | 0 | 155 | 1 | 0.019 | 0.037 |
| **C30** | Family expansion | MedGemma | AMPLIFY (Cardiovascula r / VTE) | 3 | 37 | 1 | 102 | 0.974 | 0.266 | 0.418 |
| **C31** | Family expansion | MedGemma | AMPLIFY (Cardiovascula r / VTE) | 2 | 3 | 6 | 21 | 0.333 | 0.125 | 0.182 |
| **C32** | Family expansion | MedGemma | AMPLIFY (Cardiovascula r / VTE) | 1 | 0 | 0 | 5 | 0 | 0 | 0 |
| **C33** | Family expansion | MedGemma | ARISTOTLE | 2 | 227 | 86 | 190 | 0.725 | 0.544 | 0.622 |
| **C34** | Family expansion | MedGemma | INSPIRE | 2 | 8 | 18 | 14 | 0.308 | 0.364 | 0.333 |
| **C35** | Family expansion | MedGemma | leader | 1 | 14 | 11 | 78 | 0.560 | 0.152 | 0.239 |
| **C36** | Family expansion | MedGemma | leader | 2 | 0 | 0 | 18 | 0 | 0 | 0 |
| **C37** | Family | MedGemma | leader | 2 | 8 | 0 | 32 | 1 | 0.200 | 0.333 |

| | | | | | | | | | | |
|---|---|---|---|---|---|---|---|---|---|---|
| | expansion | | | | | | | | | |
| **C38** | Family expansion | MedGemma | RECORD1 | 1 | 23 | 3 | 29 | 0.885 | 0.442 | 0.590 |
| **C39** | Family expansion | MedGemma | PRONOUNCE_ | 3 | 17 | 15 | 134 | 0.531 | 0.113 | 0.186 |
| **C40** | Family expansion | MedGemma | PRONOUNCE_ | 2 | 0 | 1 | 4 | 0 | 0 | 0 |

#### Supplementary Table S3. Exploratory mixed-effects interaction analysis

Exploratory binomial mixed-effects models tested whether final exact-code performance differed by LLM, pipeline, and criterion-complexity tier. Recall was modeled as cbind(TP, FN), and precision was modeled as cbind(TP, FP), with random intercepts for criteria nested within trial. Models were fit in R 4.6.1 using MASS::glmmPQL. Because Tier 3 included only four criteria, p-values should be interpreted as exploratory rather than confirmatory.

| Outcome | Fixed-effect term | Num df | Den df | F statistic | p value | Interpretation |
|---|---|---|---|---|---|---|
| **Recall** | model | 3 | 407 | 40.53 | <0.001 | Overall differences by LLM. |
| **Recall** | pipeline | 2 | 407 | 10.49 | <0.001 | Overall differences by pipeline. |
| **Recall** | tier | 2 | 27 | 0.54 | 0.587 | Overall differences by complexity tier. |
| **Recall** | model:pipeline | 6 | 407 | 4.19 | <0.001 | Tests whether pipeline differences vary by LLM. |
| **Recall** | model:tier | 6 | 407 | 0.94 | 0.465 | Tests whether LLM differences vary by tier. |
| **Recall** | pipeline:tier | 4 | 407 | 7.04 | <0.001 | Tests whether pipeline differences vary by tier. |
| **Recall** | model:pipeline:tier | 12 | 407 | 2.37 | 0.006 | Three-way interaction: tests whether LLM-specific pipeline effects vary by tier. |
| **Precision** | model | 3 | 381 | 9.72 | <0.001 | Overall differences by LLM. |
| **Precision** | pipeline | 2 | 381 | 46.76 | <0.001 | Overall differences by pipeline. |
| **Precision** | tier | 2 | 27 | 0.09 | 0.913 | Overall differences by complexity tier. |
| **Precision** | model:pipeline | 6 | 381 | 0.93 | 0.475 | Tests whether pipeline differences vary by LLM. |
| **Precision** | model:tier | 6 | 381 | 3.72 | 0.001 | Tests whether LLM differences vary by tier. |
| **Precision** | pipeline:tier | 4 | 381 | 7.30 | <0.001 | Tests whether pipeline differences vary by tier. |
| **Precision** | model:pipeline:tier | 12 | 381 | 1.65 | 0.077 | Three-way interaction: tests whether LLM-specific pipeline effects vary by tier. |

#### Supplementary Table S4. Bootstrap confidence intervals for micro- and macro-averaged exact-code performance by pipeline and model

Each cell reports point estimate [95% CI]. Bootstrap resampling unit is the criterion. Micro metrics aggregate exact code pairs across criteria; macro metrics average criterion-level scores.

| Pipeline | Model | Micro precision, 95% CI | Micro recall, 95% CI | Micro F1, 95% CI | Macro precision, 95% CI | Macro recall, 95% CI | Macro F1, 95% CI |
|---|---|---|---|---|---|---|---|
| **Baseline** | Claude | 0.755 [0.527-0.932] | 0.619 [0.431-0.769] | 0.680 [0.479-0.831] | 0.769 [0.660-0.865] | 0.606 [0.498-0.711] | 0.649 [0.542-0.749] |
| **Baseline** | GPT-5.5 | 0.569 [0.373-0.789] | 0.658 [0.473-0.815] | 0.610 [0.429-0.781] | 0.791 [0.705-0.868] | 0.676 [0.570-0.776] | 0.672 [0.581-0.759] |
| **Baseline** | Qwen | 0.575 [0.409-0.802] | 0.312 [0.200-0.472] | 0.405 [0.288-0.546] | 0.709 [0.591-0.815] | 0.446 [0.335-0.555] | 0.483 [0.374-0.588] |

| | | | | | | | |
|---|---|---|---|---|---|---|---|
| **Baseline** | MedGemma | 0.681 [0.547-0.838] | 0.250 [0.139-0.379] | 0.365 [0.226-0.479] | 0.596 [0.459-0.730] | 0.244 [0.158-0.342] | 0.293 [0.200-0.392] |
| **Hybrid biomedical RAG** | Claude | 0.691 [0.457-0.909] | 0.395 [0.261-0.578] | 0.503 [0.360-0.668] | 0.793 [0.689-0.886] | 0.630 [0.514-0.739] | 0.652 [0.541-0.755] |
| **Hybrid biomedical RAG** | GPT-5.5 | 0.538 [0.377-0.751] | 0.638 [0.461-0.861] | 0.584 [0.434-0.760] | 0.790 [0.703-0.871] | 0.792 [0.707-0.872] | 0.764 [0.683-0.841] |
| **Hybrid biomedical RAG** | Qwen | 0.656 [0.476-0.847] | 0.470 [0.355-0.614] | 0.548 [0.413-0.682] | 0.807 [0.720-0.886] | 0.587 [0.493-0.679] | 0.625 [0.546-0.705] |
| **Hybrid biomedical RAG** | MedGemma | 0.597 [0.406-0.727] | 0.247 [0.121-0.430] | 0.350 [0.190-0.529] | 0.536 [0.420-0.657] | 0.424 [0.311-0.541] | 0.366 [0.272-0.464] |
| **Family expansion** | Claude | 0.517 [0.385-0.689] | 0.678 [0.541-0.785] | 0.587 [0.463-0.715] | 0.708 [0.605-0.807] | 0.649 [0.547-0.748] | 0.618 [0.527-0.709] |
| **Family expansion** | GPT-5.5 | 0.329 [0.214-0.486] | 0.712 [0.559-0.839] | 0.450 [0.313-0.603] | 0.490 [0.386-0.595] | 0.671 [0.567-0.773] | 0.458 [0.379-0.538] |
| **Family expansion** | Qwen | 0.434 [0.236-0.757] | 0.555 [0.360-0.713] | 0.487 [0.298-0.693] | 0.645 [0.512-0.769] | 0.477 [0.370-0.583] | 0.485 [0.376-0.589] |
| **Family expansion** | MedGemma | 0.487 [0.254-0.691] | 0.316 [0.166-0.456] | 0.383 [0.216-0.516] | 0.442 [0.322-0.561] | 0.254 [0.174-0.340] | 0.273 [0.192-0.358] |

#### Supplementary Table S5. Deterministic verifier checks and acceptance rules

The three verifier checks are distinct from the final acceptance rules. Description-consistency flags identify potential mismatches and were not treated as confirmed clinical errors without human adjudication.

| | | | | | |
|---|---|---|---|---|---|
| **Verifier check** | Check A: Code existence and vocabulary membership | Normalize the proposed code, query the claimed local terminology table, and confirm that it exists in that vocabulary. | Exact local-table membership | verified candidate or not_found/wrong vocabulary | core/verify.py; core/tables.py |
| **Verifier check** | Check B: Description consistency | Compare the coder description with the official terminology description using thresholded token overlap. Low-overlap direct codes are held for review rather than treated as confirmed errors. | Direct code threshold = 0.34; expanded child threshold = 0.25 | verified, verified_with_warning, or review_required | core/config.py; core/verify.py |
| **Verifier check** | Check C: Controlled expansion | Expand approved parent categories, stems, ranges, and wildcard patterns deterministically against the local terminology table. | General expansion cap = 500; plain-family automatic cap = 50 | explicit child/member codes or review_required | core/config.py; core/verify.py |
| **Acceptance rule** | Output-label filtering | Exclude codes placed by the LLM under non-primary headings such as related, reminder, see also, or context from the accepted set. | Configured heading-label vocabulary | retained in detailed audit output but excluded from accepted output | core/output.py |
| **Acceptance rule** | Billability/category filtering | Exclude non-billable ICD-10 parent/category/header codes from accepted and comparison outputs when exact extraction codes are required. | ACCEPT_FINAL_BILLABLE_ONLY = True | retained in detailed output but excluded from accepted output | core/config.py; core/output.py |
| **Laboratory handling** | LOINC concept mapping | Map the laboratory test concept to LOINC; preserve thresholds, units, timing windows, and comparison operators as non-code logic. | No numerical threshold encoded as a diagnosis code | LOINC code set plus separately retained criterion logic | core/prompts.py; core/output.py |
| **Medication handling** | RxNorm RxCUI concept matching | Map medication criteria to RxCUIs while preserving drug ingredient, strength, dose form, route, brand/generic, and fixed-dose-combination distinctions. | Exact local RxNorm concept membership plus medication-specific description checks | accepted RxCUI, review_required, or not_found | core/prompts.py; core/verify.py; core/tables.py |
| **Audit/output** | Detailed and accepted exports | Write detailed verification rows, accepted exact-code rows, a comparison-ready row, and the LLM1 expert answer. | Four CSV outputs per successful run | auditable detailed output and query-ready accepted output | core/output.py |

#### Supplementary Table S6. Automated verifier impact by pipeline and model

Counts use distinct criterion-level vocabulary-code pairs after expansion. Description holds are reported as flagged potential mismatches, not confirmed errors. Expansion volumes are reported separately from the impact denominator.

| Pipeline | Model | Proposed pairs | Accepted | Absent | Wrong vocab. | Description holds | Accepted expanded | Parent expressions | Filtered non-primary | Filtered parent |
|---|---|---|---|---|---|---|---|---|---|---|
| **Baseline** | Claude | 2519 | 2314 | 15 | 0 | 59 | 1669 | 118 | 134 | 0 |
| **Baseline** | GPT-5.5 | 3681 | 3551 | 21 | 0 | 105 | 1999 | 100 | 6 | 0 |
| **Baseline** | Qwen | 1731 | 1501 | 118 | 1 | 58 | 912 | 95 | 53 | 0 |
| **Baseline** | MedGemma | 2688 | 1826 | 742 | 0 | 144 | 1729 | 110 | 22 | 0 |
| **Hybrid biomedical RAG** | Claude | 1810 | 1637 | 8 | 0 | 53 | 1371 | 140 | 163 | 0 |
| **Hybrid biomedical RAG** | GPT-5.5 | 3958 | 3635 | 37 | 0 | 151 | 1939 | 172 | 680 | 0 |
| **Hybrid biomedical RAG** | Qwen | 2151 | 1929 | 141 | 0 | 69 | 1330 | 136 | 15 | 0 |
| **Hybrid biomedical RAG** | MedGemma | 3498 | 2018 | 938 | 0 | 290 | 1697 | 182 | 278 | 0 |
| **Family expansion** | Claude | 5070 | 4524 | 11 | 0 | 30 | 4340 | 272 | 521 | 0 |
| **Family expansion** | GPT-5.5 | 8989 | 8837 | 39 | 0 | 43 | 8583 | 575 | 102 | 0 |
| **Family expansion** | Qwen | 4944 | 4640 | 37 | 8 | 34 | 4534 | 351 | 229 | 0 |
| **Family expansion** | MedGemma | 4056 | 3388 | 631 | 0 | 87 | 3360 | 248 | 0 | 0 |

#### Supplementary Table S7. Controlled-expansion volume by pipeline and model

Expansion volume is shown separately from the exact-code impact denominator. Expressions are LLM-proposed parent, family, prefix, range, or wildcard entries; accepted expanded members are the final valid child/member codes retained after deterministic verification.

| Pipeline | Model | Expansion form | Status | Expressions | Accepted expanded members | Notes |
|---|---|---|---|---|---|---|
| **Baseline** | Claude | family / prefix / range / wildcard | accepted expanded members | 118 | 1669 | Aggregated from the active verifier-impact summary after dropping two underspecified criteria. |
| **Baseline** | GPT-5.5 | family / prefix / range / wildcard | accepted expanded members | 100 | 1999 | Aggregated from the active verifier-impact summary after dropping two underspecified criteria. |
| **Baseline** | Qwen | family / prefix / range / wildcard | accepted expanded members | 95 | 912 | Aggregated from the active verifier-impact summary after dropping two underspecified criteria. |
| **Baseline** | MedGemma | family / prefix / range / wildcard | accepted expanded members | 110 | 1729 | Aggregated from the active verifier-impact summary after dropping two underspecified criteria. |
| **Hybrid biomedical RAG** | Claude | family / prefix / range / wildcard | accepted expanded members | 140 | 1371 | Aggregated from the active verifier-impact summary after dropping two underspecified criteria. |

| | | | | | | |
|---|---|---|---|---|---|---|
| **Hybrid biomedical RAG** | GPT-5.5 | family / prefix / range / wildcard | accepted expanded members | 172 | 1939 | Aggregated from the active verifier-impact summary after dropping two underspecified criteria. |
| **Hybrid biomedical RAG** | Qwen | family / prefix / range / wildcard | accepted expanded members | 136 | 1330 | Aggregated from the active verifier-impact summary after dropping two underspecified criteria. |
| **Hybrid biomedical RAG** | MedGemma | family / prefix / range / wildcard | accepted expanded members | 182 | 1697 | Aggregated from the active verifier-impact summary after dropping two underspecified criteria. |
| **Family expansion** | Claude | family / prefix / range / wildcard | accepted expanded members | 272 | 4340 | Aggregated from the active verifier-impact summary after dropping two underspecified criteria. |
| **Family expansion** | GPT-5.5 | family / prefix / range / wildcard | accepted expanded members | 575 | 8583 | Aggregated from the active verifier-impact summary after dropping two underspecified criteria. |
| **Family expansion** | Qwen | family / prefix / range / wildcard | accepted expanded members | 351 | 4534 | Aggregated from the active verifier-impact summary after dropping two underspecified criteria. |
| **Family expansion** | MedGemma | family / prefix / range / wildcard | accepted expanded members | 248 | 3360 | Aggregated from the active verifier-impact summary after dropping two underspecified criteria. |

#### Supplementary Table S8. Vocabulary-stratified performance

| Pipeline | Model | Vocabulary | Criteria | With output | Correct | Wrong | Missed | Precision | Recall | F1 |
|---|---|---|---|---|---|---|---|---|---|---|
| **Baseline** | Claude | ICD9 | 33 | 32 | 436 | 91 | 334 | 0.827 | 0.566 | 0.672 |
| **Baseline** | Claude | ICD10 | 33 | 32 | 859 | 247 | 339 | 0.777 | 0.717 | 0.746 |
| **Baseline** | Claude | LOINC | 1 | 1 | 4 | 2 | 5 | 0.667 | 0.444 | 0.533 |
| **Baseline** | Claude | RxNorm | 6 | 5 | 5 | 3 | 36 | 0.625 | 0.122 | 0.204 |
| **Baseline** | GPT-5.5 | ICD9 | 33 | 32 | 409 | 289 | 361 | 0.586 | 0.531 | 0.557 |
| **Baseline** | GPT-5.5 | ICD10 | 33 | 33 | 921 | 961 | 277 | 0.489 | 0.769 | 0.598 |
| **Baseline** | GPT-5.5 | LOINC | 4 | 4 | 5 | 17 | 4 | 0.227 | 0.556 | 0.323 |
| **Baseline** | GPT-5.5 | RxNorm | 6 | 6 | 8 | 1 | 33 | 0.889 | 0.195 | 0.320 |
| **Baseline** | Qwen | ICD9 | 33 | 31 | 227 | 124 | 543 | 0.647 | 0.295 | 0.405 |
| **Baseline** | Qwen | ICD10 | 33 | 32 | 423 | 432 | 775 | 0.495 | 0.353 | 0.412 |
| **Baseline** | Qwen | LOINC | 4 | 4 | 2 | 9 | 7 | 0.182 | 0.222 | 0.200 |
| **Baseline** | Qwen | RxNorm | 7 | 3 | 2 | 5 | 39 | 0.286 | 0.049 | 0.083 |
| **Baseline** | MedGemma | ICD9 | 33 | 22 | 141 | 96 | 629 | 0.595 | 0.183 | 0.280 |
| **Baseline** | MedGemma | ICD10 | 35 | 27 | 301 | 288 | 897 | 0.511 | 0.251 | 0.337 |
| **Baseline** | MedGemma | LOINC | 1 | 1 | 1 | 0 | 8 | 1 | 0.111 | 0.200 |
| **Baseline** | MedGemma | RxNorm | 6 | 0 | 0 | 0 | 41 | 0 | 0 | 0 |
| **Hybrid biomedical RAG** | Claude | ICD9 | 33 | 24 | 262 | 93 | 508 | 0.738 | 0.340 | 0.466 |
| **Hybrid biomedical RAG** | Claude | ICD10 | 33 | 31 | 591 | 318 | 607 | 0.650 | 0.493 | 0.561 |
| **Hybrid biomedical RAG** | Claude | LOINC | 1 | 1 | 4 | 2 | 5 | 0.667 | 0.444 | 0.533 |
| **Hybrid biomedical RAG** | Claude | RxNorm | 6 | 6 | 24 | 6 | 17 | 0.800 | 0.585 | 0.676 |
| **Hybrid biomedical RAG** | GPT-5.5 | ICD9 | 33 | 33 | 496 | 350 | 274 | 0.586 | 0.644 | 0.614 |
| **Hybrid biomedical RAG** | GPT-5.5 | ICD10 | 33 | 33 | 704 | 1003 | 494 | 0.412 | 0.588 | 0.485 |
| **Hybrid biomedical RAG** | GPT-5.5 | LOINC | 5 | 5 | 4 | 17 | 5 | 0.190 | 0.444 | 0.267 |
| **Hybrid biomedical RAG** | GPT-5.5 | RxNorm | 6 | 6 | 35 | 1 | 6 | 0.972 | 0.854 | 0.909 |
| **Hybrid biomedical RAG** | Qwen | ICD9 | 33 | 33 | 296 | 73 | 474 | 0.802 | 0.384 | 0.520 |
| **Hybrid biomedical RAG** | Qwen | ICD10 | 33 | 32 | 625 | 604 | 573 | 0.509 | 0.522 | 0.515 |
| **Hybrid** | Qwen | LOINC | 2 | 2 | 1 | 4 | 8 | 0.200 | 0.111 | 0.143 |

| | | | | | | | | | | |
|---|---|---|---|---|---|---|---|---|---|---|
| **biomedical RAG** | | | | | | | | | | |
| **Hybrid biomedical RAG** | Qwen | RxNorm | 6 | 6 | 17 | 0 | 24 | 1 | 0.415 | 0.586 |
| **Hybrid biomedical RAG** | MedGemma | ICD9 | 33 | 24 | 116 | 100 | 654 | 0.537 | 0.151 | 0.235 |
| **Hybrid biomedical RAG** | MedGemma | ICD10 | 36 | 31 | 399 | 235 | 799 | 0.629 | 0.333 | 0.436 |
| **Hybrid biomedical RAG** | MedGemma | LOINC | 4 | 4 | 2 | 18 | 7 | 0.100 | 0.222 | 0.138 |
| **Hybrid biomedical RAG** | MedGemma | RxNorm | 6 | 5 | 23 | 53 | 18 | 0.303 | 0.561 | 0.393 |
| **Family expansion** | Claude | ICD9 | 33 | 32 | 396 | 186 | 374 | 0.680 | 0.514 | 0.586 |
| **Family expansion** | Claude | ICD10 | 33 | 33 | 958 | 860 | 240 | 0.527 | 0.800 | 0.635 |
| **Family expansion** | Claude | LOINC | 4 | 4 | 4 | 10 | 5 | 0.286 | 0.444 | 0.348 |
| **Family expansion** | Claude | RxNorm | 6 | 5 | 5 | 1 | 36 | 0.833 | 0.122 | 0.213 |
| **Family expansion** | GPT-5.5 | ICD9 | 33 | 33 | 488 | 663 | 282 | 0.424 | 0.634 | 0.508 |
| **Family expansion** | GPT-5.5 | ICD10 | 33 | 33 | 932 | 2131 | 266 | 0.304 | 0.778 | 0.437 |
| **Family expansion** | GPT-5.5 | LOINC | 4 | 4 | 4 | 17 | 5 | 0.190 | 0.444 | 0.267 |
| **Family expansion** | GPT-5.5 | RxNorm | 6 | 6 | 9 | 1 | 32 | 0.900 | 0.220 | 0.353 |
| **Family expansion** | Qwen | ICD9 | 33 | 30 | 371 | 587 | 399 | 0.387 | 0.482 | 0.429 |
| **Family expansion** | Qwen | ICD10 | 33 | 31 | 879 | 802 | 319 | 0.523 | 0.734 | 0.611 |
| **Family expansion** | Qwen | LOINC | 5 | 5 | 2 | 9 | 7 | 0.182 | 0.222 | 0.200 |
| **Family expansion** | Qwen | RxNorm | 6 | 2 | 2 | 1 | 39 | 0.667 | 0.049 | 0.091 |
| **Family expansion** | MedGemma | ICD9 | 33 | 15 | 162 | 117 | 608 | 0.581 | 0.210 | 0.309 |
| **Family expansion** | MedGemma | ICD10 | 33 | 29 | 426 | 785 | 772 | 0.352 | 0.356 | 0.354 |
| **Family expansion** | MedGemma | LOINC | 2 | 2 | 1 | 1 | 8 | 0.500 | 0.111 | 0.182 |
| **Family expansion** | MedGemma | RxNorm | 6 | 0 | 0 | 0 | 41 | 0 | 0 | 0 |

#### Supplementary Table S9. Verifier input form counts

Counts summarize the active automated verifier-impact dataset after removing two underspecified criteria. The dataset covered 40 criteria across 11 trial groups, with many more verifier rows because family, member, wildcard, and range expressions can expand into multiple exact codes. The refreshed dataset contained 51,898 verifier rows, including 11,183 single-code rows and 40,715 family, member, wildcard, or range-derived rows.

| Form | Verifier rows |
|---|---|
| **Single-code verifier rows** | 11,183 |
| **Family/member/wildcard/range-derived verifier rows** | 40,715 |
| **All active verifier rows** | 51,898 |

#### Supplementary Table S10. Staged verifier ablation by pipeline and model

Micro-averaged metrics aggregate exact code pairs within each pipeline-model-stage combination. Macro-averaged metrics average criterion-level performance and therefore give each criterion equal weight.

| Pipeline | Model | Stage | Criteria | Correct | Wrong | Missed | Micro precision | Micro recall | Micro F1 | Micro Delta F1 | Macro precision | Macro recall | Macro F1 | Mean criterion Delta F1 | Median criterion Delta F1 |
|---|---|---|---|---|---|---|---|---|---|---|---|---|---|---|---|
| **Baseline** | Claude | Before verifier | 40 | 546 | 273 | 1750 | 0.667 | 0.238 | 0.351 | 0.000 | 0.624 | 0.497 | 0.510 | 0.000 | 0.000 |
| **Baseline** | Claude | Existence/vocabulary check | 40 | 546 | 248 | 1750 | 0.688 | 0.238 | 0.353 | 0.003 | 0.641 | 0.497 | 0.518 | 0.009 | 0.000 |
| **Baseline** | Claude | Acceptance filtering | 40 | 507 | 212 | 1789 | 0.705 | 0.221 | 0.336 | -0.014 | 0.705 | 0.473 | 0.515 | 0.005 | 0.000 |
| **Baseline** | Claude | Final verifier + expansion | 40 | 1421 | 460 | 875 | 0.755 | 0.619 | 0.680 | 0.330 | 0.769 | 0.606 | 0.649 | 0.139 | 0.042 |
| **Baseline** | GPT-5.5 | Before verifier | 40 | 1216 | 505 | 1080 | 0.707 | 0.530 | 0.605 | 0.000 | 0.755 | 0.669 | 0.670 | 0.000 | 0.000 |
| **Baseline** | GPT-5.5 | Existence/vocabulary check | 40 | 1215 | 485 | 1081 | 0.715 | 0.529 | 0.608 | 0.003 | 0.773 | 0.669 | 0.672 | 0.003 | 0.000 |
| **Baseline** | GPT-5.5 | Acceptance filtering | 40 | 1138 | 454 | 1158 | 0.715 | 0.496 | 0.585 | -0.020 | 0.806 | 0.648 | 0.662 | -0.008 | 0.000 |
| **Baseline** | GPT-5.5 | Final verifier + expansion | 40 | 1510 | 1145 | 786 | 0.569 | 0.658 | 0.610 | 0.005 | 0.791 | 0.676 | 0.672 | 0.003 | 0.000 |
| **Baseline** | Qwen | Before verifier | 40 | 427 | 359 | 1869 | 0.543 | 0.186 | 0.277 | 0.000 | 0.540 | 0.387 | 0.405 | 0.000 | 0.000 |
| **Baseline** | Qwen | Existence/vocabulary check | 40 | 426 | 251 | 1870 | 0.629 | 0.186 | 0.287 | 0.009 | 0.601 | 0.387 | 0.432 | 0.027 | 0.000 |
| **Baseline** | Qwen | Acceptance filtering | 40 | 405 | 214 | 1891 | 0.654 | 0.176 | 0.278 | 0.001 | 0.658 | 0.367 | 0.426 | 0.020 | 0.000 |

| | | | | | | | | | | | | | | | |
|---|---|---|---|---|---|---|---|---|---|---|---|---|---|---|---|
| **Baseline** | Qwen | Final verifier + expansion | 40 | 717 | 529 | 1579 | 0.575 | 0.312 | 0.405 | 0.128 | 0.709 | 0.446 | 0.483 | 0.077 | 0.003 |
| **Baseline** | MedGemma | Before verifier | 40 | 67 | 814 | 2229 | 0.076 | 0.029 | 0.042 | 0.000 | 0.194 | 0.091 | 0.097 | 0.000 | 0.000 |
| **Baseline** | MedGemma | Existence/vocabulary check | 40 | 66 | 220 | 2230 | 0.231 | 0.029 | 0.051 | 0.009 | 0.258 | 0.089 | 0.116 | 0.019 | 0.000 |
| **Baseline** | MedGemma | Acceptance filtering | 40 | 50 | 116 | 2246 | 0.301 | 0.022 | 0.041 | -0.002 | 0.292 | 0.078 | 0.104 | 0.007 | 0.000 |
| **Baseline** | MedGemma | Final verifier + expansion | 40 | 573 | 268 | 1723 | 0.681 | 0.250 | 0.365 | 0.323 | 0.596 | 0.244 | 0.293 | 0.196 | 0.006 |
| **Hybrid biomedical RAG** | Claude | Before verifier | 40 | 253 | 274 | 2043 | 0.480 | 0.110 | 0.179 | 0.000 | 0.478 | 0.462 | 0.409 | 0.000 | 0.000 |
| **Hybrid biomedical RAG** | Claude | Existence/vocabulary check | 40 | 253 | 265 | 2043 | 0.488 | 0.110 | 0.180 | 0.001 | 0.483 | 0.462 | 0.413 | 0.004 | 0.000 |
| **Hybrid biomedical RAG** | Claude | Acceptance filtering | 40 | 208 | 184 | 2088 | 0.531 | 0.091 | 0.155 | -0.024 | 0.517 | 0.425 | 0.422 | 0.012 | 0.000 |
| **Hybrid biomedical RAG** | Claude | Final verifier + expansion | 40 | 907 | 405 | 1389 | 0.691 | 0.395 | 0.503 | 0.324 | 0.793 | 0.630 | 0.652 | 0.243 | 0.158 |
| **Hybrid biomedical RAG** | GPT-5.5 | Before verifier | 40 | 1342 | 664 | 954 | 0.669 | 0.584 | 0.624 | 0.000 | 0.725 | 0.790 | 0.732 | 0.000 | 0.000 |
| **Hybrid biomedical RAG** | GPT-5.5 | Existence/vocabulary check | 40 | 1341 | 623 | 955 | 0.683 | 0.584 | 0.630 | 0.006 | 0.741 | 0.790 | 0.738 | 0.006 | 0.000 |
| **Hybrid biomedical RAG** | GPT-5.5 | Acceptance filtering | 40 | 1272 | 510 | 1024 | 0.714 | 0.554 | 0.624 | -0.000 | 0.801 | 0.765 | 0.754 | 0.022 | 0.009 |
| **Hybrid biomedical RAG** | GPT-5.5 | Final verifier + expansion | 40 | 1465 | 1256 | 831 | 0.538 | 0.638 | 0.584 | -0.040 | 0.790 | 0.792 | 0.764 | 0.032 | 0.023 |
| **Hybrid biomedical RAG** | Qwen | Before verifier | 40 | 496 | 424 | 1800 | 0.539 | 0.216 | 0.308 | 0.000 | 0.575 | 0.469 | 0.465 | 0.000 | 0.000 |
| **Hybrid biomedical RAG** | Qwen | Existence/vocabulary check | 40 | 496 | 305 | 1800 | 0.619 | 0.216 | 0.320 | 0.012 | 0.659 | 0.469 | 0.485 | 0.020 | 0.006 |
| **Hybrid biomedical RAG** | Qwen | Acceptance filtering | 40 | 459 | 268 | 1837 | 0.631 | 0.200 | 0.304 | -0.005 | 0.728 | 0.446 | 0.489 | 0.024 | 0.005 |
| **Hybrid biomedical RAG** | Qwen | Final verifier + expansion | 40 | 1080 | 567 | 1216 | 0.656 | 0.470 | 0.548 | 0.239 | 0.807 | 0.587 | 0.625 | 0.160 | 0.080 |
| **Hybrid biomedical RAG** | MedGemma | Before verifier | 40 | 228 | 1501 | 2068 | 0.132 | 0.099 | 0.113 | 0.000 | 0.214 | 0.309 | 0.194 | 0.000 | 0.000 |

| | | | | | | | | | | | | | | | |
|---|---|---|---|---|---|---|---|---|---|---|---|---|---|---|---|
| **Hybrid biomedical RAG** | MedGemma | Existence/vocabulary check | 40 | 227 | 656 | 2069 | 0.257 | 0.099 | 0.143 | 0.030 | 0.257 | 0.308 | 0.223 | 0.028 | 0.000 |
| **Hybrid biomedical RAG** | MedGemma | Acceptance filtering | 40 | 186 | 336 | 2110 | 0.356 | 0.081 | 0.132 | 0.019 | 0.323 | 0.285 | 0.242 | 0.048 | 0.000 |
| **Hybrid biomedical RAG** | MedGemma | Final verifier + expansion | 40 | 568 | 383 | 1728 | 0.597 | 0.247 | 0.350 | 0.237 | 0.536 | 0.424 | 0.366 | 0.171 | 0.048 |
| **Family expansion** | Claude | Before verifier | 40 | 134 | 330 | 2162 | 0.289 | 0.058 | 0.097 | 0.000 | 0.389 | 0.252 | 0.267 | 0.000 | 0.000 |
| **Family expansion** | Claude | Existence/vocabulary check | 40 | 134 | 316 | 2162 | 0.298 | 0.058 | 0.098 | 0.000 | 0.404 | 0.252 | 0.276 | 0.009 | 0.000 |
| **Family expansion** | Claude | Acceptance filtering | 40 | 122 | 273 | 2174 | 0.309 | 0.053 | 0.091 | -0.006 | 0.447 | 0.238 | 0.275 | 0.008 | 0.000 |
| **Family expansion** | Claude | Final verifier + expansion | 40 | 1557 | 1453 | 739 | 0.517 | 0.678 | 0.587 | 0.490 | 0.708 | 0.649 | 0.618 | 0.351 | 0.333 |
| **Family expansion** | GPT-5.5 | Before verifier | 40 | 152 | 718 | 2144 | 0.175 | 0.066 | 0.096 | 0.000 | 0.283 | 0.251 | 0.208 | 0.000 | 0.000 |
| **Family expansion** | GPT-5.5 | Existence/vocabulary check | 40 | 152 | 597 | 2144 | 0.203 | 0.066 | 0.100 | 0.004 | 0.299 | 0.251 | 0.212 | 0.005 | 0.000 |
| **Family expansion** | GPT-5.5 | Acceptance filtering | 40 | 147 | 542 | 2149 | 0.213 | 0.064 | 0.098 | 0.002 | 0.320 | 0.245 | 0.213 | 0.006 | 0.000 |
| **Family expansion** | GPT-5.5 | Final verifier + expansion | 40 | 1634 | 3328 | 662 | 0.329 | 0.712 | 0.450 | 0.354 | 0.490 | 0.671 | 0.458 | 0.250 | 0.181 |
| **Family expansion** | Qwen | Before verifier | 40 | 91 | 351 | 2205 | 0.206 | 0.040 | 0.066 | 0.000 | 0.251 | 0.170 | 0.169 | 0.000 | 0.000 |
| **Family expansion** | Qwen | Existence/vocabulary check | 40 | 91 | 243 | 2205 | 0.272 | 0.040 | 0.069 | 0.003 | 0.312 | 0.170 | 0.181 | 0.012 | 0.000 |
| **Family expansion** | Qwen | Acceptance filtering | 40 | 85 | 221 | 2211 | 0.278 | 0.037 | 0.065 | -0.001 | 0.334 | 0.163 | 0.180 | 0.011 | 0.000 |
| **Family expansion** | Qwen | Final verifier + expansion | 40 | 1274 | 1661 | 1022 | 0.434 | 0.555 | 0.487 | 0.421 | 0.645 | 0.477 | 0.485 | 0.316 | 0.166 |
| **Family expansion** | MedGemma | Before verifier | 40 | 84 | 864 | 2212 | 0.089 | 0.037 | 0.052 | 0.000 | 0.126 | 0.078 | 0.079 | 0.000 | 0.000 |
| **Family expansio** | MedGemma | Existence/vocabulary check | 40 | 74 | 274 | 2222 | 0.213 | 0.032 | 0.056 | 0.004 | 0.148 | 0.076 | 0.086 | 0.006 | 0.000 |

| | | | | | | | | | | | | | | | |
|---|---|---|---|---|---|---|---|---|---|---|---|---|---|---|---|
| **n** | | | | | | | | | | | | | | | |
| **Family expansion** | MedGemma | Acceptance filtering | 40 | 59 | 201 | 2237 | 0.227 | 0.026 | 0.046 | -0.006 | 0.142 | 0.060 | 0.073 | -0.006 | 0.000 |
| **Family expansion** | MedGemma | Final verifier + expansion | 40 | 726 | 766 | 1570 | 0.487 | 0.316 | 0.383 | 0.332 | 0.442 | 0.254 | 0.273 | 0.194 | 0.138 |

#### Supplementary Table S11. Stage-transition removal and addition counts

Transition counts compare adjacent staged outputs. Removed true-reference codes are codes present in the reference standard but lost at that transition; removed false positives are non-reference codes removed at that transition. Added true-reference and added false-positive codes summarize codes introduced by deterministic expansion.

| Pipeline | Model | Transition | Criteria | Removed total | Removed true-reference | Removed false-positive | Added total | Added true-reference | Added false-positive |
|---|---|---|---|---|---|---|---|---|---|
| **Baseline** | Claude | existence_vocab_only -> filtered_no_expansion | 40 | 75 | 39 | 36 | 0 | 0 | 0 |
| **Baseline** | Claude | filtered_no_expansion -> final_verifier | 40 | 172 | 46 | 126 | 1334 | 960 | 374 |
| **Baseline** | Claude | pre_verification -> existence_vocab_only | 40 | 25 | 0 | 25 | 0 | 0 | 0 |
| **Baseline** | GPT-5.5 | existence_vocab_only -> filtered_no_expansion | 40 | 108 | 77 | 31 | 0 | 0 | 0 |
| **Baseline** | GPT-5.5 | filtered_no_expansion -> final_verifier | 40 | 98 | 0 | 98 | 1161 | 372 | 789 |
| **Baseline** | GPT-5.5 | pre_verification -> existence_vocab_only | 40 | 21 | 1 | 20 | 0 | 0 | 0 |
| **Baseline** | Qwen | existence_vocab_only -> filtered_no_expansion | 40 | 58 | 21 | 37 | 0 | 0 | 0 |
| **Baseline** | Qwen | filtered_no_expansion -> final_verifier | 40 | 87 | 1 | 86 | 714 | 313 | 401 |
| **Baseline** | Qwen | pre_verification -> existence_vocab_only | 40 | 109 | 1 | 108 | 0 | 0 | 0 |
| **Baseline** | MedGemma | existence_vocab_only -> filtered_no_expansion | 40 | 120 | 16 | 104 | 0 | 0 | 0 |
| **Baseline** | MedGemma | filtered_no_expansion -> final_verifier | 40 | 79 | 0 | 79 | 754 | 523 | 231 |
| **Baseline** | MedGemma | pre_verification -> existence_vocab_only | 40 | 595 | 1 | 594 | 0 | 0 | 0 |
| **Hybrid biomedical RAG** | Claude | existence_vocab_only -> filtered_no_expansion | 40 | 126 | 45 | 81 | 0 | 0 | 0 |
| **Hybrid** | Claude | filtered_no_expansion | 40 | 117 | 0 | 117 | 1037 | 699 | 338 |

| | | | | | | | | | |
|---|---|---|---|---|---|---|---|---|---|
| **biomedical RAG** | | -> final_verifier | | | | | | | |
| **Hybrid biomedical RAG** | Claude | pre_verification -> existence_vocab_only | 40 | 9 | 0 | 9 | 0 | 0 | 0 |
| **Hybrid biomedical RAG** | GPT-5.5 | existence_vocab_only -> filtered_no_expansion | 40 | 182 | 69 | 113 | 0 | 0 | 0 |
| **Hybrid biomedical RAG** | GPT-5.5 | filtered_no_expansion -> final_verifier | 40 | 119 | 1 | 118 | 1058 | 194 | 864 |
| **Hybrid biomedical RAG** | GPT-5.5 | pre_verification -> existence_vocab_only | 40 | 42 | 1 | 41 | 0 | 0 | 0 |
| **Hybrid biomedical RAG** | Qwen | existence_vocab_only -> filtered_no_expansion | 40 | 74 | 37 | 37 | 0 | 0 | 0 |
| **Hybrid biomedical RAG** | Qwen | filtered_no_expansion -> final_verifier | 40 | 120 | 1 | 119 | 1040 | 622 | 418 |
| **Hybrid biomedical RAG** | Qwen | pre_verification -> existence_vocab_only | 40 | 119 | 0 | 119 | 0 | 0 | 0 |
| **Hybrid biomedical RAG** | MedGemma | existence_vocab_only -> filtered_no_expansion | 40 | 361 | 41 | 320 | 0 | 0 | 0 |
| **Hybrid biomedical RAG** | MedGemma | filtered_no_expansion -> final_verifier | 40 | 132 | 0 | 132 | 561 | 382 | 179 |
| **Hybrid biomedical RAG** | MedGemma | pre_verification -> existence_vocab_only | 40 | 846 | 1 | 845 | 0 | 0 | 0 |
| **Family expansion** | Claude | existence_vocab_only -> filtered_no_expansion | 40 | 55 | 12 | 43 | 0 | 0 | 0 |
| **Family expansion** | Claude | filtered_no_expansion -> final_verifier | 40 | 213 | 0 | 213 | 2828 | 1435 | 1393 |
| **Family expansion** | Claude | pre_verification -> existence_vocab_only | 40 | 14 | 0 | 14 | 0 | 0 | 0 |
| **Family expansion** | GPT-5.5 | existence_vocab_only -> filtered_no_expansion | 40 | 60 | 5 | 55 | 0 | 0 | 0 |
| **Family expansion** | GPT-5.5 | filtered_no_expansion -> final_verifier | 40 | 401 | 0 | 401 | 4674 | 1487 | 3187 |
| **Family expansion** | GPT-5.5 | pre_verification -> existence_vocab_only | 40 | 121 | 0 | 121 | 0 | 0 | 0 |
| **Family expansion** | Qwen | existence_vocab_only -> filtered_no_expansion | 40 | 28 | 6 | 22 | 0 | 0 | 0 |
| **Family** | Qwen | filtered_no_expansion | 40 | 194 | 0 | 194 | 2823 | 1189 | 1634 |

| | | | | | | | | | |
|---|---|---|---|---|---|---|---|---|---|
| **expansion** | | -> final_verifier | | | | | | | |
| **Family expansion** | Qwen | pre_verification -> existence_vocab_only | 40 | 108 | 0 | 108 | 0 | 0 | 0 |
| **Family expansion** | MedGemma | existence_vocab_only -> filtered_no_expansion | 40 | 88 | 15 | 73 | 0 | 0 | 0 |
| **Family expansion** | MedGemma | filtered_no_expansion -> final_verifier | 40 | 160 | 0 | 160 | 1392 | 667 | 725 |
| **Family expansion** | MedGemma | pre_verification -> existence_vocab_only | 40 | 600 | 10 | 590 | 0 | 0 | 0 |
